# Equity in protection: bridging global data gaps for an EBV vaccine – a systematic review and meta-analysis

**DOI:** 10.1101/2025.05.28.25328417

**Authors:** Marisa D. Muckian, Ting Shi, Vesa Qarkaxhija, Simran Kapoor, Tomos Morgan, Helen R. Stagg

## Abstract

**Introduction:** Epstein-Barr virus (EBV) is linked to multiple malignancies and autoimmune conditions, with different disease burdens globally. Pharmaceutical companies and researchers are placing substantial investment in the development of EBV vaccines. To ensure optimal vaccine roll-out, particularly in resource-limited settings, it is essential to have data on the age at acquisition of EBV. We aimed to systematically review and meta-analyse seroprevalence by age by country, World Health Organization (WHO) region, and country income level; to identify knowledge gaps; and determine an approach to bridge these gaps.

**Methods:** MEDLINE, Embase, and Web of Science databases were searched on March 22, 2022, for studies that measured EBV seroprevalence by age. An updated search was conducted on October 22, 2022. There were no language restrictions. Papers were assessed for quality using an adapted version of the Downs and Black checklist. Seroprevalence by age was estimated using a fixed-effect (country) or random-effects (WHO region, income) meta-analysis. This review has been registered on PROSPERO (CRD42022349900).

**Results:** Only one country (United States) had enough data for a country meta-analysis. WHO Regional analyses revealed the Western Pacific region to have a higher seroprevalence at younger age groups than other WHO regions. Country income level better explained seroprevalence trends per age. Middle-income countries displayed a quicker rise to equilibrium seroprevalence than high-income countries, with a 30% absolute increase in 0- to 4-year-olds in middle-income than high-income countries (59% [95% CI 28–91%, I^2^ =99%] versus 29% [95% CI 16–41%, I^2^ =99%], respectively).

**Conclusion:** This first meta-analysis producing estimates of EBV seroprevalence by age, provides crucial information to guide governments when choosing if to implement a vaccine for EBV. However, data variability and limited consistency of methodologies and EBV seroprevalence measurements hindered comprehensive meta-analyses across all WHO regions and countries. We provide an interim framework for the extrapolation of seroprevalence using country-specific income levels to aid vaccine rollout decisions.

**Key Messages:** *What is already known on this topic:* This study built on a previous systematic review evaluating the global literature to identify risk factors associated with EBV acquisition. The review searched three databases, (MEDLINE, Embase, and Web of Science), for studies investigating EBV risk factors (including age), on March 16 2017. No meta-analysis was performed, limiting the conclusions that could be drawn, but when data were grouped by country and WHO region it was revealed that EBV seroconversion tends to occur at younger ages in Asia versus Europe and North America. A substantial data gap was identified for countries in Africa and South America.

*What this study adds:* Our study documents more than double the number of publications on EBV seroprevalence by age globally and provides the first meta-analysis on the topic, which will be vital for governments seeking to deploy an EBV vaccine in their country. In the face of a dearth of data for many countries (particularly low- and middle-income countries, such as China, where EBV-associated nasopharyngeal carcinoma is endemic), we demonstrate how country income can be used to group studies and thus extrapolate to fill data gaps.

*How this study might affect research, practice or policy:* The deployment strategy and cost-effectiveness case for an EBV vaccine will vary substantially among countries due to geographic variability in a) the burden of EBV-associated diseases and b) when infection is acquired. To inform deployment, it is critical to either have country-by-country data on the age at acquisition or a means to extrapolate from another appropriate setting. Our study informs policymakers on both points, by providing both meta-analysis estimates where possible and an interim framework for extrapolation of data among countries before new seroprevalence studies arise. This is particularly critical for low- and middle-income countries where data are sparse.

## Introduction

Epstein-Barr virus (EBV) is known to be associated with the development of several malignancies, including nasopharyngeal carcinoma (NPC) and Hodgkin’s lymphoma (HL), Burkitt’s lymphoma (BL), as well as autoimmune conditions including multiple sclerosis (MS).[1,2] The incidence of these illnesses varies from country-to-country. NPC, a cancer of the neck and throat, shows a distinct geographic distribution and is endemic to areas in Southeast Asia, such as southern China, Taiwan and Hong Kong.[3] BL also shows a geographic distribution, with high incidence in Equatorial Africa.[4] Endemic BL (eBL) is the most common childhood EBV-associated cancer in countries where malaria is also endemic.[4] The burden of MS is much greater in high-income countries, particularly in the Northern hemisphere, such as Sweden.[5] These illnesses carry a high economic burden, not least by costs associated with diagnosis and treatment where health care resources may be limited. Because these diseases usually affect individuals in their productive years, there is a subsequent negative impact on the economic output of the country. Moreover, the impact of these diseases extends beyond their economic burden. These illnesses contribute to a decreased quality of life and fatality. Therefore, the prevention of EBV is imperative and of great interest to global health.

Efforts to develop an EBV vaccine have spanned decades, focusing initially on the gp350 antigen, which showed promise in animal models by eliciting neutralising antibodies but failed to prevent infection in humans.[6] Subsequent attempts included subunit vaccines targeting multiple viral glycoproteins, yet these too were only modestly effective. The complexity of EBV’s life cycle, involving distinct entry mechanisms into B cells and epithelial cells, has posed significant challenges.[6] Recent advances, including virus-like particles (VLPs) and mRNA-based vaccines, offer renewed hope. Three vaccine candidates aiming to prevent the incidence of infectious mononucleosis are currently in phase I clinical trials at the time of writing, offering new hope to the quest to produce an implementable candidate. [7–9]

Due to the high number of EBV-associated illnesses and their profound impact on countries across the globe, the implementation of preventative measures through vaccination has the potential to mitigate their impact on health. If indeed a prophylactic vaccine candidate for EBV was developed and rolled out globally, it is important to understand how best to deploy the vaccine to improve public health, particularly where resources are limited.

Understanding the seroprevalence of EBV across different age groups per country plays a critical role in informing policymakers which age groups to target with a future vaccine. In 2017, Winter et al. published a systematic review of the global literature on risk factors for the acquisition of EBV, within which age was one of the risk factors of interest. [10] EBV is known to have an equilibrium seroprevalence of 95% worldwide, although the age of acquisition varies among populations. Although this review did not include a meta-analysis, EBV seroconversion was found to occur in younger age groups in Asia and older age groups in Europe and North America. A large data gap was uncovered for countries in Africa and South America. The data that were available for these regions were reported only from specific populations, such as HIV-exposed infants, [11] and thus had limited generalisability and were for very specific ages.

As the possibility of a prophylactic EBV vaccine becomes greater,[12–14] it is imperative for recommending bodies to understand EBV seroprevalence by age geographically, including the evidence gaps that need to be filled prior to vaccine licensing and implementation. Together with the country-specific burden of EBV-associated disease data, this will inform decision-making for vaccine deployment. In this study, we systematically reviewed the globally available data and, building on the work by Winter et al, [10] present seroprevalence estimates by age, and then country, and WHO region. Further, to better explain how countries group by their seroprevalence as a function of age, we additionally present estimates by country income level.

## Methods

This systematic review was reported in accordance with the PRISMA guidelines.

### Search strategy and study selection

The search strategy for this review was adapted from Winter et al.[10] The search terms included EBV, infectious mononucleosis, glandular fever, seroprevalence, and study design terms such as ‘case control’, ‘cohort’, ‘intervention study’, ‘cross sectional’, and ‘clinical trial’. Additional terms related to public health surveillance and monitoring were also included (Supplementary table 1). Unlike the previous review, age was our sole risk factor of interest. Studies were included if they reported seroprevalence for specific age groups and excluded if they reported seroprevalence estimates for broad age ranges or only contained people with EBV-associated disease. Studies measuring all EBV antibody-antigen combinations were included. The full inclusion and exclusion criteria are presented in Supplementary table 2.

The databases MEDLINE, Embase, and Web of Science were searched on March 22, 2022, for studies from March 7, 2017. Those published prior to that point were taken from Winter et al. [10] An updated search was conducted on October 22, 2022, with a 2-week overlap with the March 2022 search.

### Screening

The studies returned from the search were uploaded to the systematic review management software, Covidence, where title, abstract, and full-text screening was carried out in duplicate by MM and either VQ or SK. Conflicts were flagged by Covidence and resolved by consensus. Cohen’s kappa statistic for measuring agreement between reviewers was calculated.[15] The full texts of studies from the Winter et al. [10] review were also screened for suitability. Foreign language papers were screened by fluent speakers of the language together with MM.

### Data extraction, synthesis, and quality assessment

Data was extracted to a predesigned Microsoft Excel spreadsheet and was performed by MM and TM, recording relevant information such as study design, study population, years of study, country, EBV test type, antibody used, antigen measured, overall seroprevalence, and age-specific seroprevalence. Disagreements were resolved through consensus, and foreign language papers were extracted by fluent speakers of the respective language along with MM.

Age categories were created for the analysis due to the impracticality of measuring and meta-analysing seroprevalence per year of age. The predetermined age groups were 0–4, 5–9, 10–14, 15–19, 20–29, 30–39, and ≥40 years, as it was not expected studies would report single age years. Where the age groups described in a study did not precisely match these categories it was placed into a category if it included an age range of 2 years on either side of the upper and lower age group studied. Studies were grouped by WHO region and country income level according to the World Bank designation. [16,17]

Quality assessment was conducted using the Downs and Black[18] checklist, which was adapted as per the guidance from Deeks et al[19] (Supplementary table 3). This included questions surrounding information bias, misclassification, and measurement of the exposure and the outcome. The assessment included whether the paper’s primary aim was to assess seroprevalence per age and whether the age groups analysed were drawn from the same populations. Detecting seroprevalence using manufacturers’ kits were deemed less susceptible to information bias for the outcome compared to in-house tests.

All data was descriptively analysed, including by generating seroprevalence plots by country, WHO region, and country income level. Only studies measuring anti−viral capsid antigen immunoglobulin G (VCA IgG), as this antibody-antigen combination is most indicative of previous infection, were included from this point. Where possible, meta-analyses of proportions were carried out using metaprop in STATA version 17. The metaprop command automatically applies a logit transformation to the seroprevalence proportions prior to pooling, which stabilises variances and accounts for proportions near zero or one. The pooled estimates are then back transformed to the original proportion scale for interpretation and presentation. Metaprop also provides I^2^ estimates to estimate heterogeneity between studies. Meta-analyses were conducted globally per age group and then, WHO region, country, and country income level to determine potential sources of heterogeneity. A minimum of three studies was required for each age group for meta-analysis. Fixed-effect meta-analysis was deemed appropriate for analyses per country due to the presence of a single underlying effect estimate and random effects for the WHO region and income analyses because we did not anticipate this expectation to be fulfilled.

Where multiple papers/data points from the same study and age group were present, a sensitivity analysis was performed that restricted the analysis to one data point per study to ensure that no individual study was influencing the overall seroprevalence estimate. We also examined the impact of the calendar years in which the study was conducted on seroprevalence estimates, using the midpoint of the years in which people were recruited to the study.

Meta-regression analyses were performed using Stata version 17 to percentage of between-study heterogeneity explained by the predictors income level (high vs. middle) and region (Europe vs. Americas) on EBV seroprevalence. Random effects meta-regressions were conducted for each age group where there was sufficient data to do so; the dependent variable was the logit-transformed proportion of EBV seroprevalence.

### Publication bias

Whilst publication bias was unlikely to have been an issue for the studies included in this paper due to their descriptive nature (across different age groups), for the global meta-analyses we plotted funnel plots for each age group. For subgroups with ≥10 studies, Egger’s regression test was applied to assess funnel plot asymmetry. Analyses were conducted in Stata using the metafunnel and metabias commands.

### Registration

This review has been registered on PROSPERO (CRD42022349900).

## Results

### Search Results

After deduplication, the literature search returned 5914 papers. Thirty-nine studies[20–58] met our inclusion criteria at the full-text stage (figure 1, Supplementary table 4). An additional 32 studies[11,31,59–88] were included from the previous review (figure 1, Supplementary table 4), bringing the total to 71. Cohen’s kappa statistic between MM and SK and MM and VQ were 0·76 and 0·61, respectively, indicating substantial agreement among reviewers.

Papers were present from all WHO regions and across 25 different countries. After full-text extraction, it was found that these 71 papers represented 68 studies. Three studies appeared in multiple papers.[33,34,36,37,44,45,71] In this instance, seroprevalence data were usually taken from the most recent publication. Most of these studies measured seroprevalence using VCA IgG, with the next most common antigen-antibody combination being EBNA1 IgG. A small number of studies measured VCA or EBNA1 IgM/IgA (Supplementary table 4).

### Overall Quality Assessment

Studies were assessed against 12 domains of quality (Supplementary table 5). Of the 71 papers, 30 (42·3%)[11,23,24,26–28,30,31,37,38,43–48,50,51,53,55,57,62,64,65,74,75,77,81,84,86] were aiming to examine seroprevalence by age and were assessed accordingly. All studies drew all age groups from the same populations and time periods. Only eleven studies (11/71, 15·4%)[33,35–37,39,41,54,60,68,71,78] used tests for seroprevalence that may have introduced information bias, per investigator assessment. Of these 11, only one set out to measure seroprevalence by age.

### Meta-analysis

Firstly, we performed a meta-analysis of proportions across all the studies where three studies were available for each age group. This analysis revealed substantial heterogeneity (Supplementary figure 1), particularly within the younger age groups. To explore potential sources of this heterogeneity, we performed subgroup analyses and meta-regression, stratifying by WHO region and World Bank income level.

We also undertook an analysis of potential publication bias. Funnel plots for all age groups are presented in Supplementary Figures 2A-G and Egger’s test results in Supplementary table 6. The only age group with potential publication bias was 20–29 year-olds, but it is unlikely that such a phenomenon could have played out for this age group independently of the others.

### WHO region analyses

Next, we present descriptive analyses and meta-analyses of the studies included in our review. We initially grouped studies by country and WHO region as per Winter et al.[10]

#### African Region

Five papers [11,22,49,74,82] were included from the African region (AFR), including studies from (Ghana, Kenya, Malawi, and Zambia Supplementary figure 3). Four of these studies focussed on young children, with ages ranging between 0-11 years old. Only one study was carried out in adults (18-65 years old). No meta-analyses were possible for this WHO region.

#### European Region

There was a total of 19 studies[21,23,24,35,37–40,45,52,54,55,57–59,70,75,79,85]from the European Region (EUR) in populations spanning aged 0–85 years old (Supplementary figure 4). Overall, seroprevalence was estimated to rapidly increase until the age of 15 years, then more slowly thereafter. Barring a single study in the UK that showed a decreased seroprevalence after the age of 70 that may have suffered from survival bias[79], data were relatively consistent.

Meta-analyses were performed for the European region as one per age category, except for age group 15-19 (0–4, 5–9, 10–14, 20–29, 30–39, ≥40), using data from 14 separate studies[16,17,28,30–32,38,45,47,48,50–52,63,72] (figure 2, Supplementary figure 5). Seroprevalence per age category increased rapidly until age 7, before gradually increasing to 96% at age 35. I^2^ values varied between 79% and 99% across the meta-analyses, indicating that the percentage of total variability due to between-study heterogeneity was high.

Removing instances where there were duplicate data points from a single study within a sensitivity analysis did not impact seroprevalence estimates (Supplementary figure 6).

#### Region of the Americas

There was a total of nine studies from the Americas region (AMR) spanning ages from 0-85 years old (Supplementary figure 5).[27,30,53,56,60–62,65,66] Studies were mostly from the USA (n=6/9, 66·7%),with three from Brazil (33·3%). Patients’ ages ranged from 0 to 85 years. The two nations provided very distinct estimates of seroprevalence (Supplementary Figure 7). Country specific trends for the USA and Brazil are discussed in the Country Analyses section of the results.

Four studies were included in the meta-analysis for the AMR (Supplementary figure 8); three studies or greater were available for age categories 0–4, 5–9, and 16–19 only. Seroprevalence was higher than in the European region for these age groups, due to the higher seroprevalence in Brazil influencing this estimate. I^2^ values varied between 90% and 99% across the meta-analyses, indicating that the percentage of total variability due to between-study heterogeneity was high.

Removing instances where there were duplicate data points from a single study within a sensitivity analysis did not impact seroprevalence estimates (Supplementary figure 9).

#### Southeast Asia Region

One paper from the Southeast Asia region (SEA) was included, from Thailand[84] (Supplementary figure 10). This study covered many age groups, from 0 to 6 months old until >40 years old. Seroprevalence increased to 100% at ages 6–8 years old and stayed consistently over 96% into the older age categories (≥40). No meta-analyses were possible.

#### Eastern Mediterranean Region

There was only one study from the Eastern Mediterranean region (EMR), which was from Iran[47] (Supplementary figure 11). The total age range of these studies was 0 to ≥85 (maximum age not stated). Seroprevalence in Iran increased gradually with age from 50·5% at ages 1–3 years old and reached 92% by age 20–29 years old. No meta-analyses were possible.

#### Western Pacific Region

Five of the included studies were from the Western Pacific region (WPR), China/Taiwan (n=4),[28,48,64,87] and Singapore (n=1)[46] (Supplementary figure 12). The age range across these papers was from 0–85 years old. Seroprevalence was around 66% between ages 0 and 5 years old and rapidly increased to 99%–100% by age 30. Likely due to the prevalence of NPC in this region (particularly in specific geographic regions of China), contrary to previous regions, most studies measured either IgA or IgM antibodies. Data were consistent within China. No meta-analyses were possible.

#### WHO region meta-regression

Region explained a portion of variation in seroprevalence for two younger (0-4 year olds, and 5*-*9 year olds; R^2^=3.1% and 25.1%, respectively), nand two older age categories (20-29, >40s; 25.3% and 28.5%, respectively), (Supplementary table 7).

### Country income-level analyses

As grouping studies by WHO region did not result in consistent seroprevalence curves across the countries, we then grouped the included papers into different country income levels based on where they were conducted. The total number of papers found for each income level (high, medium, and low) at the time of the study, were 47/71 (63·4%),[23–27,31–46,52,54–59,61,62,65,66,68–73,75,77–79,81,83,85,86]21/71 (28·2%),[20,21,28–30,47,48,50,51,53,60,63,64,67,76,80,84,87,88] and 4/71 (6·6%),[11,22,74,82]respectively. One study (from Kenya)[49] did not report the time period of their study and, therefore, income level was not determined.

#### High-Income

Across the 47 high-income studies, data were available from 0 to 85 years of age. Broadly, seroprevalence increased gradually between ages 0 and 25, peaking at 90–100% by around age 25 (Supplementary 13A). One study showed a decrease in seroprevalence after age 70; this may be due to survivor bias.[79]

In the meta-analysis, seroprevalence steadily increased to the age of 30 years (figure 3, Supplementary figure 14) I^2^ values varied between 79% and 99% across the meta-analyses, indicating that the percentage of total variability due to between-study heterogeneity was very high (with the lowest levels in people over the age of 40 years).

Removing instances where there were duplicate data points from a single study within a sensitivity analysis did not impact seroprevalence estimates (Supplementary 15).

#### Middle-Income

Across the 21 middle-income papers, seroprevalence data were available for ages ranging between 0 and 85 years. Two main groupings of studies appeared, one with a more rapid approach to the equilibrium of seroprevalence and one with a shallower approach (Supplementary 13B), although both had steeper trajectories than the high-income studies.

Six of the papers were included in the meta-analysis,[28,30,47,53,60,84] across five of the seven age categories (figure 2, Supplementary figure 16). Meta-analysis was not available for 10- to 14- and 15- to 19-year-olds. Data showed a steep increase in seroprevalence up to the middling teenage years, followed by a slower increase. I^2^ values varied between 46% and 99% across the meta-analyses, indicating that the percentage of total variability due to between-study heterogeneity was high. Again, data were less variable for individuals over age 40.

Removing instances where there were duplicate data points from a single study within a sensitivity analysis did not impact seroprevalence estimates (Supplementary figure 17).

Compared to the high-income countries, seroprevalence in the middle-income countries had a much sharper increase between the ages of 0 and 10 years, with almost 90% seroprevalence being reached by 10 years old (figure 3).

#### Low-Income

Finally, four papers included populations from low-income countries at the time of the study (Supplementary figure 13C). [11,22,74,82] The age range across these papers was 0 to 65 years old. The seroprevalence in the low-income countries was much higher at younger age groups in comparison to both high- and middle-income countries, with one study reaching ∼90% seroprevalence by age 5. [22] A further study from Ghana was an outlier.[11]

It was not possible to meta-analyse the low-income country papers.

#### Country income-level Income Level Meta-regression

Meta-regression was possible for all age groups apart from 15-19 years (Supplementary table 9). For 0-4 year olds, 5-9 year olds (in particular) and 10-14 year olds, more variation was explained by income level than WHO region, which is in line with the greater degree of difference in seroprevalence between settings for these age groups.

### Country Analyses

The papers included in this review covered 25 countries, with 17 reporting seroprevalence data using VCA IgG (Supplementary Table 6). SEA and EMR only had one study each, and thus their studies are described above.

In AFR, both Kenya and Malawi demonstrated sharp increases in seropositivity at very young ages (Supplementary Figure 3C, D). Only Ghana (Supplementary Figure 3B) had data for adults, with substantially lower seroprevalences, even for the younger adult age groups than the other African studies.

It was possible to examine data from eight EUR nations: UK (n=6), [35,38,45,57,75,79]Sweden (n=3),[24,39,70]the Netherlands (n=3),[40,54,59,85]Croatia (n=2),[23,55] Germany, Finland, Turkey, and Greece (n=1 each),[21,37,52,58]but not meta-analyse any of them (Supplementary 4b-i). Only Croatia, Finland, Turkey, and the UK had data across the lifespan. Within nations, data were reasonably consistent.

There were a total of three studies from Brazil (Supplementary figure 7A).[30,53,60] The seroprevalence across age groups appeared to be 80-90% by 10 years old. In contrast the six studies from the USA,[27,56,61,62,65,66] showed a much slower increase in seroprevalence (Supplementary figure 7B). It was possible to meta-analyse for the US individually (Supplementary figure 18). Three studies[62,65,66] were included in the meta-analysis, for age categories 5–9, 10–14 and 15–19 only. Seroprevalence was similar across each group.

Removing instances where there were duplicate data points from a single study within a sensitivity analysis did not impact seroprevalence estimates (Supplementary figure 19) At the country level, we further sought to examine whether there was any impact of the calendar years in which the study was conducted on seroprevalence estimates. The only country where this was possible was the USA (Supplementary figure 20). For the 10-14 and 15-19 year olds no obvious trend was observed. Estimates for 5-9 year olds showed some indication of lower seroprevalence in later studies, albeit across limited datapoints, with overlapping CIs, and with two different estimates from Delaney *et al*.[66].

## Discussion

This systematic review and meta-analysis provide critical insights into EBV seroprevalence across different countries, income levels, and age groups. One of the key findings from this study is the clear distinction in EBV seroprevalence trends between high- and middle-income countries. Middle-income countries demonstrate a faster rise to equilibrium seroprevalence, particularly among children aged 0-4 years, compared to high-income countries where infection tends to occur later. Specifically, a 30% higher seroprevalence was noted in middle-income countries (59% vs. 29% in high-income countries) for this age group.

Our review is the first to synthesise and meta-analyse global EBV seroprevalence by age, including presenting data grouped by country, WHO region, and by country income level. We provide recommending bodies with critical information that will be informative when governments are considering implementing targeted vaccine strategies. We double the number of eligible papers from the original Winter et al review.[10] Despite this, we did identify substantial gaps in the literature for low-and-middle income countries, however this review does provide evidence of a sharper rise in EBV seroprevalence among younger children in middle-and-low-income countries. This highlights the role of a complex array of social and economic factors, including childcare practices and living conditions, in facilitating early transmission.[89] The meta-regression results in this study suggest that income level explains more of the variation in EBV seroprevalence, particularly in the 5-9 year old age group. Income also explained variation in seroprevalence among 10–14-year-olds, where regional analysis showed no explanatory power. This suggests that economic factors may play a larger role in shaping early-life transmission patterns.

The lack of data in the African, Eastern Mediterranean, and Southeast Asia regions makes it difficult for policymakers to get a reliable estimate of EBV seroprevalence per age group. Studies from South America fall into the WHO Americas region (AMR) and while this review included two studies from South America, both from Brazil; extrapolating the AMR findings to other South American countries should be done cautiously. It is crucial for future research efforts to include a broader range of countries in South America. However, we do provide a way to extrapolate data to countries with missing data, based on their income level.

While this systematic review synthesised a large body of global data on EBV seroprevalence, the limitations need to be acknowledged. The meta-analyses displayed substantial heterogeneity across studies with I² values (up to 99%). This review was also limited by data variability, due to different methodologies and the fact that not all studies used VCA IgG antibodies to determine seroprevalence. VCA IgG is the only antigen-antibody combination that has persistent titres detected in both acute and past infection.[90] These issues limited our ability to meta-analyse for most WHO regions and countries. Underlying studies were often restricted to specific populations; for example, in the African region, studies were limited to women and infants, with no data available for men or middle-aged categories. Limited numbers of data points for the meta-analyses also meant that we were unable to undertake sensitivity analyses to examine the impact of study quality on our estimates.

To address the identified limitations, future research should focus on filling the substantial data gaps in low- and middle-income countries. These regions currently lack sufficient age-stratified seroprevalence data, which is essential for guiding vaccine deployment strategies. It is possible that EBV seroprevalence estimates, particularly for younger age groups in high income settings, are changing over time, but we lacked the data to explore this fully. Ideally, such analyses would be taken from a single data source over a series of calendar years. Future studies should use gold-standard serological assays measuring VCA IgG to ensure the accuracy and reliability of the measured seroprevalence in order to avoid non-differential misclassification. Studies also need to be conducted to maximise their generalisability within a setting (i.e. with a representative sample to the general population and with a large enough sample size per age group).

The findings of this review highlight that age is a critical factor to consider when deciding how to deploy a vaccine to prevent EBV infection. Preventing EBV and its associated illnesses is crucial, and the age of acquisition of infection, duration of protection from a vaccine, and the burden of EBV-associated disease in a country will affect the roll-out of a vaccine. [91] Approximately half of China’s population is EBV seropositive by the age of 5, and thus it would be critical to vaccinate children under this age. A vaccine could potentially be delivered at the same time as other infant vaccinations. Given that China has NPC endemic areas, cost-effectiveness is more likely than in a country with a lower burden of EBV-associated disease. As the onset of NPC is commonly many decades after EBV infection, [92] a lengthy duration of protection is likely to be required from a vaccine, although booster doses could also be used. In Uganda, where BL is highly prevalent, the age of vaccination will be similar to that of China. As BL predominantly affects children in that country, the duration of protection is not as critical. [93] In contrast, in high-income countries such as Sweden (where HL and MS, which are both associated with IM, are of concern), [94,95] infection is often acquired in the teenage years and thus deployment could wait until later in life than in China and Uganda. A vaccine with a lower duration of protection may be less problematic in such countries if it is assumed that delayed infection will take individuals outside of the highest risk period for IM and thus disease.

## Conclusion

In conclusion, this study provides the most current global analysis and meta-analyses of EBV seroprevalence by age across the globe. Knowledge of EBV acquisition per age is useful for future EBV vaccine campaigns, to ensure they are as beneficial and cost-effective as possible. We note an unfortunate data gap which means that seroprevalence as a function of age cannot be parameterised for LMIC; however, we provide an interim framework for extrapolation which would allow for informed decision-making about vaccine roll-out in these countries before additional studies are performed.

## Author Contributions

MDM acted as a guarantor. MDM and HRS were responsible conceptualisation and study design and interpretation of the results. MDM was also responsible for conducting the literature search, screening papers, analyses, and preparation of the manuscript. TM, SK and VQ were each responsible for screening the papers for inclusion in the systematic review. TM also conducted quality assessment of the papers along with MDM. TS assisted with data analysis, providing methodological expertise and assistance with the interpretation the of results. All authors contributed to editing the manuscript.

## Competing interests

We declare no competing interests.

## Funding

This study was funded by Moderna, Inc. (103661ED). The funders had no role in the study design, analysis, and interpretation of data; in the writing of the paper; nor in the decision to submit the paper for publication.

## Data Sharing

All data is included in the published article and its appendices.

## Ethics Statements

No ethical approval needed

## Patient and Public Involvement

Patients and the public were not involved in the design, conduct, reporting, or dissemination plans of our research.

## Data Availability

All data is included in the published article and its appendices.

## Acknowledgments

The authors would like to acknowledge the funders of this project, Moderna, Inc. We give special thanks to Dr Aysenur Kilic and Alex Dieiev for assisting with the translation of papers not in English. We would like to thank MEDiSTRAVA for their support in compiling this manuscript.

---

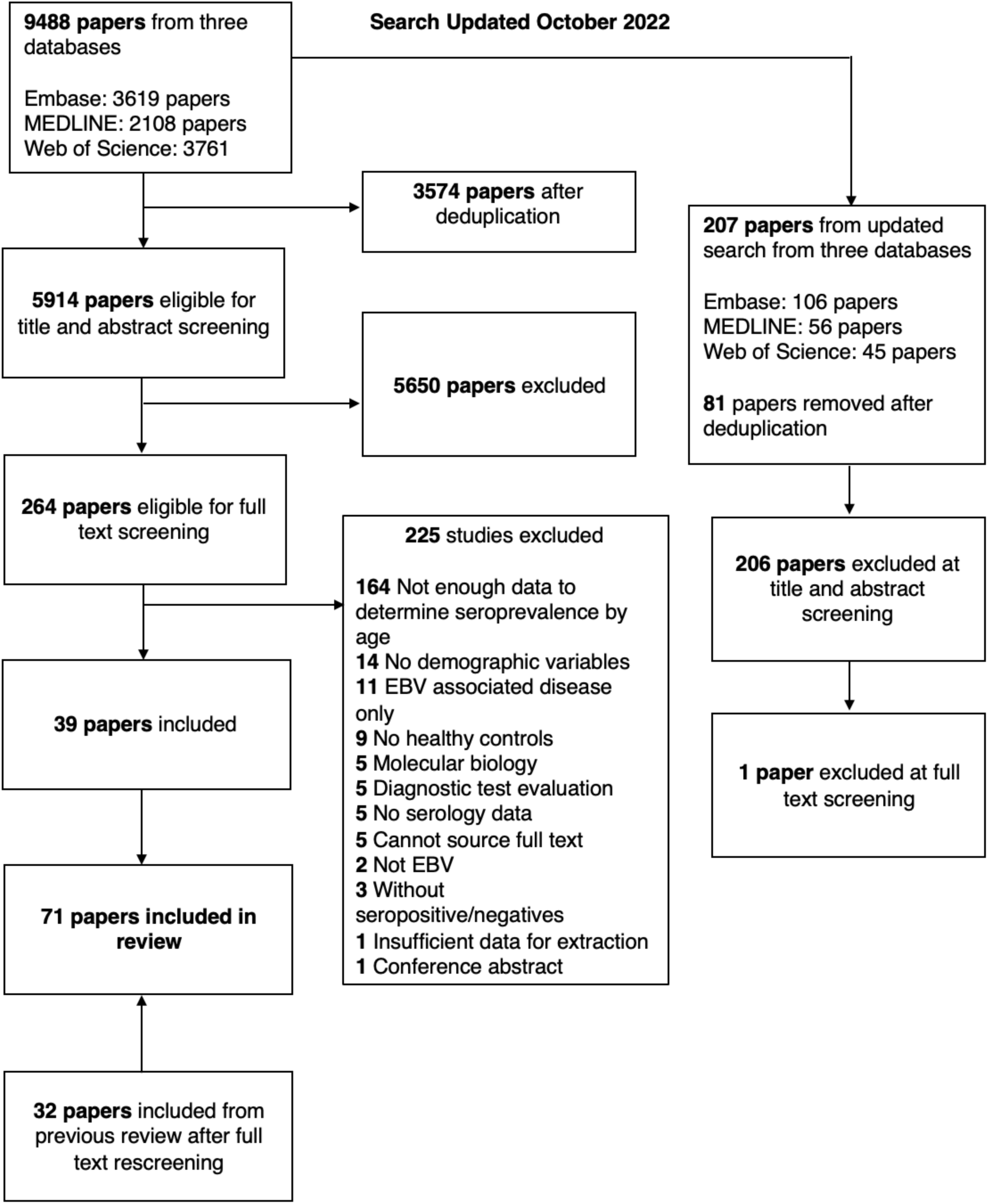

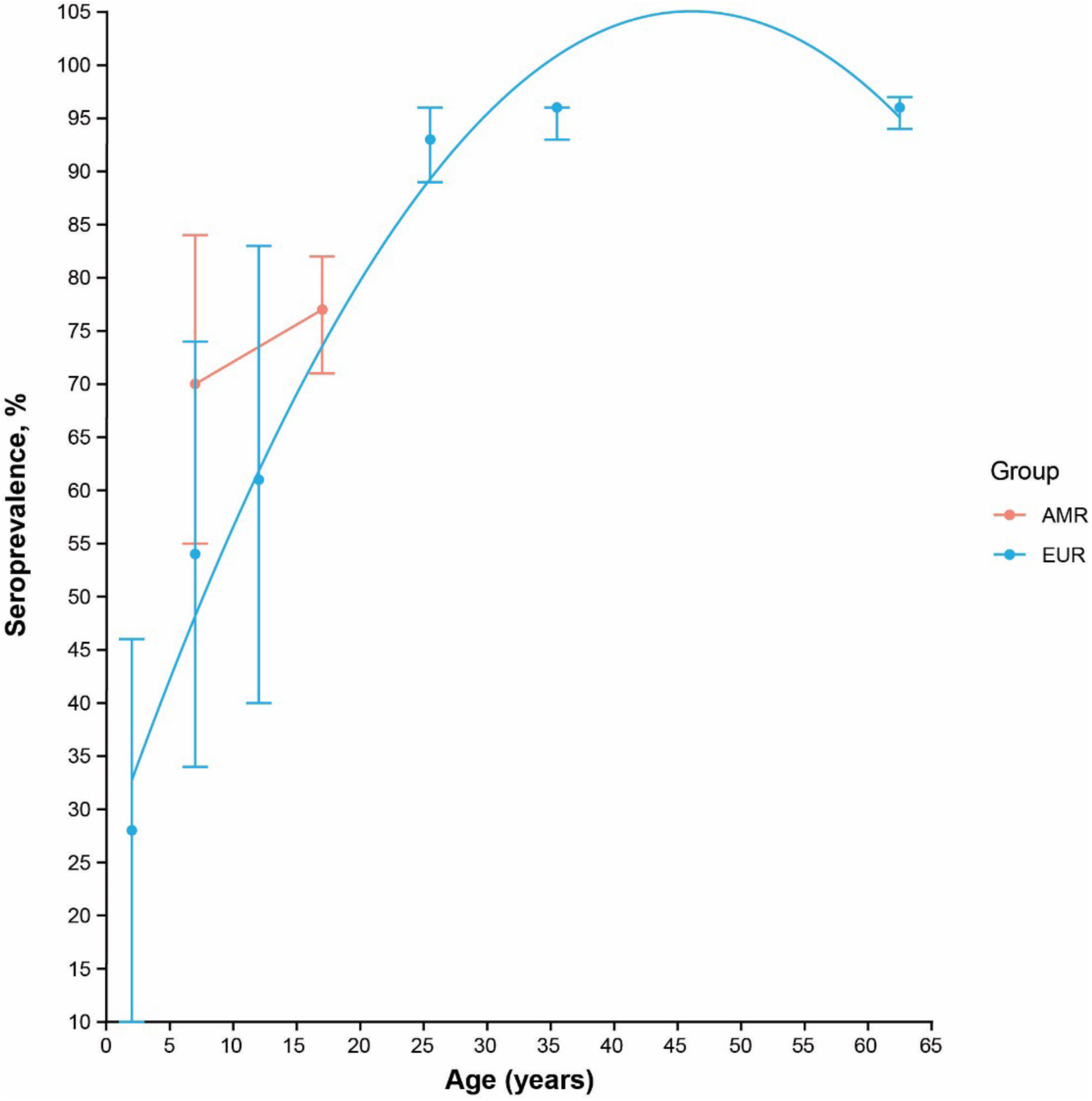

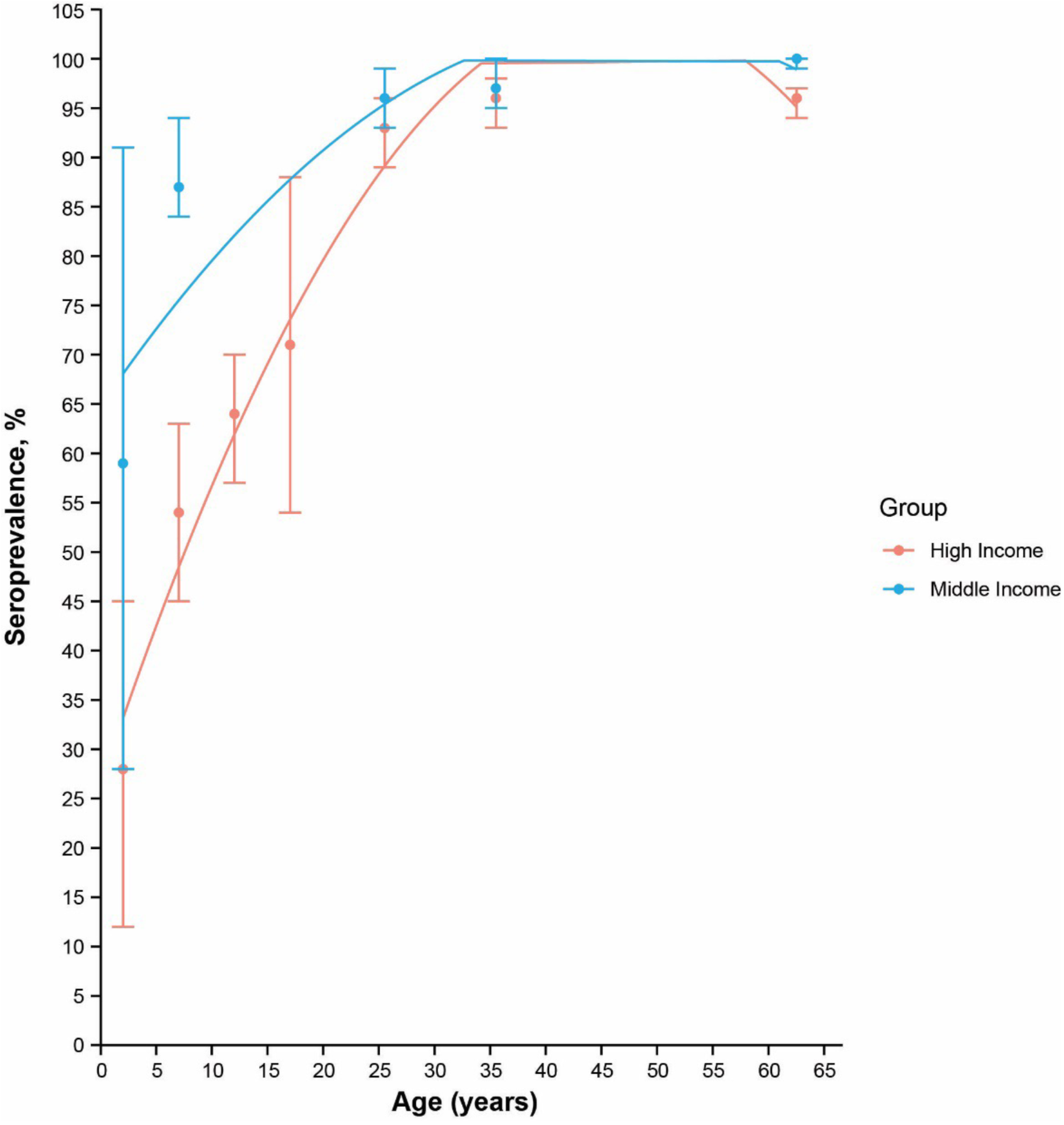

## Notes

### Competing Interest Statement

The authors have declared no competing interest.

