## Supplementary Materials for "Equity in protection: bridging global data gaps for an EBV vaccine – a systematic review and meta-analysis"

**Table of Contents**


Supplementary Table 1. Search terms 2

Supplementary Table 2 - Inclusion and Exclusion Criteria 7

Supplementary Table 3. Quality assessment questions 8

Supplementary Table 4. Included studies 9

Supplementary Table 5. Quality assessment 19

Supplementary Figure 1. Seroprevalence by age (forest plots) Globally. 25

Supplementary Figure 2 Funnel plots to assess publication bias 32

Supplementary Table 6. Egger's regression test for funnel plot asymmetry 36

Supplementary Figure 3. Seroprevalence by age plotted for the World Health Organization African Region 37

Supplementary Figure 4. Seroprevalence by age plotted for the World Health Organization European Region 38

Supplementary Figure 5. Seroprevalence by age (forest plots) for the European Region 40

Supplementary Figure 6. Seroprevalence by age (forest plots) for the European Region (Sensitivity analysis) 42

Supplementary Figure 7. Seroprevalence by age plotted for the World Health Organization Region of the Americas 43

Supplementary Figure 8. Seroprevalence by age (forest plots) for the Region of the Americas 44

Supplementary Figure 9. Seroprevalence by age (forest plots) for the Region of the Americas (sensitivity analyses) 45

Supplementary Figure 10. Seroprevalence by age plotted for the World Health Organization South-East Asia Region 46

Supplementary Figure 11. Seroprevalence by age plotted for the World Health Organization Eastern Mediterranean Region 47

Supplementary Figure 12. Seroprevalence by age plotted for the World Health Organization Western Pacific Region 48

Supplementary Table 7. Meta-regression results for region (EUR at baseline vs AMR) 49

Supplementary Figure 13. Seroprevalence by age plotted by country income level according to the World Bank50

Supplementary Figure 14. Seroprevalence by age (forest plots) in high-income countries 51

Supplementary Figure 15. Seroprevalence by age (forest plots) for high-income countries (sensitivity analysis) 54

Supplementary Figure 16. Seroprevalence by age (forest plots) for middle-income countries 56

Supplementary Figure 17. Seroprevalence by age (forest plots) for middle-income countries (sensitivity analysis) 58

Supplementary Table 8. Meta-regression results for income-level (High-income at baseline vs middle-income) 59

Supplementary Table 9. Number of studies per country 60

Supplementary Figure 18. Seroprevalence by age forest plots for the USA 61

Supplementary Figure 19. Seroprevalence by age forest plots for the USA (sensitivity analyses) 62

Supplementary Figure 20. Cumulative seroprevalence by age forest plots for the USA. 63

**Supplementary Table 1. Search terms**

### MEDLINE

| # | Query | Results |
|---|---|---|
| 1 | herpesvirus 4, human.mp. or exp Herpesvirus 4, Human/ | 25 466 |
| 2 | limit 1 to yr="2017 -Current" | 3339 |
| 3 | epstein barr virus.mp. | 36 982 |
| 4 | limit 3 to yr="2017 -Current" | 7151 |
| 5 | epstein-barr virus.mp. | 36 982 |
| 6 | limit 5 to yr="2017 -Current" | 7151 |
| 7 | EBV.mp. | 28 688 |
| 8 | limit 7 to yr="2017 -Current" | 6168 |
| 9 | EB virus.mp. | 950 |
| 10 | limit 9 to yr="2017 -Current" | 106 |
| 11 | HHV-4.mp. | 48 |
| 12 | limit 11 to yr="2017 -Current" | 19 |
| 13 | HHV4.mp. | 23 |
| 14 | limit 13 to yr="2017 -Current" | 7 |
| 15 | HHV 4.mp. | 48 |
| 16 | limit 15 to yr="2017 -Current" | 19 |
| 17 | exp Infectious Mononucleosis/ or infectious mononucleosis.mp. | 8852 |
| 18 | limit 17 to yr="2017 -Current" | 572 |
| 19 | glandular fever.mp. | 273 |
| 20 | limit 19 to yr="2017 -Current" | 12 |
| 21 | 2 or 4 or 6 or 8 or 10 or 12 or 14 or 16 or 18 or 20 | 8762 |
| 22 | seropositiv*.mp. [mp=title, abstract, original title, name of substance word, subject heading word, floating sub-heading word, keyword heading word, organism supplementary concept word, protocol supplementary concept word, rare disease supplementary concept word, unique identifier, synonyms] | 61 037 |
| 23 | limit 22 to yr="2017 -Current" | 10 442 |
| 24 | seronegativ*.mp. [mp=title, abstract, original title, name of substance word, subject heading word, floating sub-heading word, keyword heading word, organism supplementary concept word, protocol supplementary concept word, rare disease supplementary concept word, unique identifier, synonyms] | 23 043 |
| 25 | limit 24 to yr="2017 -Current" | 4 109 |
| 26 | exp Seroepidemiologic Studies/ or seroepidemiol*.mp. | 29 079 |
| 27 | limit 26 to yr="2017 -Current" | 6370 |
| 28 | seroprevalence.mp. | 23 366 |
| 29 | limit 28 to yr="2017 -Current" | 7138 |

| | | |
|---|---|---|
| 30 | 23 or 25 or 27 or 29 | 18 253 |
| 31 | epidemiol*.mp. | 2 191 707 |
| 32 | limit 30 to yr="2017 -Current" | 18 253 |
| 33 | exp Epidemiology/ | 28 044 |
| 34 | limit 33 to yr="2017 -Current" | 3912 |
| 35 | risk factor.mp. or exp Risk Factors/ | 1 052 298 |
| 36 | limit 35 to yr="2017 -Current" | 284 534 |
| 37 | exp Cross-Sectional Studies/ or cross-sectional.mp. | 578 755 |
| 38 | limit 37 to yr="2017 -Current" | 245 690 |
| 39 | exp Cohort Studies/ or cohort.mp. | 2 588 477 |
| 40 | limit 39 to yr="2017 -Current" | 807 745 |
| 41 | exp Case-Control Studies/ or case control.mp. | 1 343 196 |
| 42 | limit 41 to yr="2017 -Current" | 454 323 |
| 43 | case-control.mp. | 368 685 |
| 44 | limit 43 to yr="2017 -Current" | 103 512 |
| 45 | exp Clinical Trial/ | 932 991 |
| 46 | limit 45 to yr="2017 -Current" | 160 555 |
| 47 | intervention study.mp. | 11 314 |
| 48 | limit 47 to yr="2017 -Current" | 4083 |
| 49 | exp Public Health Surveillance/ | 5130 |
| 50 | limit 49 to yr="2017 -Current" | 3459 |
| 51 | exp Health Surveys/ | 610 977 |
| 52 | limit 51 to yr="2017 -Current" | 127 539 |
| 53 | (Monitor* or Surveillance or Survey*).mp. [mp=title, abstract, original title, name of substance word, subject heading word, floating sub-heading word, keyword heading word, organism supplementary concept word, protocol supplementary concept word, rare disease supplementary concept word, unique identifier, synonyms] | 2 386 860 |
| 54 | limit 53 to yr="2017 -Current" | 724 619 |
| 55 | 32 or 34 or 36 or 38 or 40 or 42 or 44 or 46 or 48 or 50 or 52 or 54 | 1 845 268 |
| 56 | Humans/ | 20 277 141 |
| 57 | limit 56 to yr="2017 -Current" | 3 633 551 |
| 58 | 30 or 55 | 1 845 268 |
| 59 | 21 and 56 and 58 | 2108 |

**EMBASE**

| # | Query | Results |
|---|---|---|
| 1 | herpesvirus 4, human.mp. or exp Herpesvirus 4, Human/ | 43 411 |
| 2 | limit 1 to yr="2017 -Current" | 8322 |
| 3 | epstein barr virus.mp. | 58 972 |
| 4 | limit 3 to yr="2017 -Current" | 13 713 |
| 5 | epstein-barr virus.mp. | 58 972 |
| 6 | limit 5 to yr="2017 -Current" | 13 713 |
| 7 | EBV.mp. | 41 556 |
| 8 | limit 7 to yr="2017 -Current" | 11 141 |
| 9 | EB virus.mp. | 1165 |
| 10 | limit 9 to yr="2017 -Current" | 171 |
| 11 | HHV-4.mp. | 79 |
| 12 | limit 11 to yr="2017 -Current" | 30 |
| 13 | HHV4.mp. | 35 |
| 14 | limit 13 to yr="2017 -Current" | 9 |
| 15 | HHV 4.mp. | 79 |
| 16 | limit 15 to yr="2017 -Current" | 30 |
| 17 | exp Infectious Mononucleosis/ or infectious mononucleosis.mp. | 8649 |
| 18 | limit 17 to yr="2017 -Current" | 1243 |
| 19 | glandular fever.mp. | 172 |
| 20 | limit 19 to yr="2017 -Current" | 24 |
| 21 | 2 or 4 or 6 or 8 or 10 or 12 or 14 or 16 or 18 or 20 | 17 895 |
| 22 | seropositiv*.mp. | 53 650 |
| 23 | limit 22 to yr="2017 -Current" | 13 750 |
| 24 | seronegativ*.mp. | 26 929 |
| 25 | limit 24 to yr="2017 -Current" | 6392 |
| 26 | exp seroprevalence/ or exp seroepidemiology/ or seroepidemiol*.mp. | 33 291 |
| 27 | limit 26 to yr="2017 -Current" | 12 032 |
| 28 | seroprevalence.mp. | 35 040 |
| 29 | limit 28 to yr="2017 -Current" | 12 158 |
| 30 | 23 or 25 or 27 or 29 | 26 182 |
| 31 | epidemiol*.mp. | 1 706 195 |
| 32 | limit 31 to yr="2017 -Current" | 373 353 |
| 33 | exp Epidemiology/ | 3 978 241 |
| 34 | limit 33 to yr="2017 -Current" | 1 399 760 |

| | | |
|---|---|---|
| 35 | risk factor.mp. or exp Risk Factors/ | 1 345 774 |
| 36 | limit 35 to yr="2017 -Current" | 463 324 |
| 37 | exp Cross-Sectional Studies/ or cross-sectional.mp. | 689 536 |
| 38 | limit 37 to yr="2017 -Current" | 326 366 |
| 39 | exp Cohort Studies/ or cohort.mp. | 1 331 631 |
| 40 | limit 39 to yr="2017 -Current" | 689 711 |
| 41 | exp Case-Control Studies/ or case control.mp. | 266 947 |
| 42 | limit 41 to yr="2017 -Current" | 91 341 |
| 43 | case-control.mp. | 266 947 |
| 44 | limit 43 to yr="2017 -Current" | 91 341 |
| 45 | exp Clinical Trial/ | 1 681 599 |
| 46 | limit 45 to yr="2017 -Current" | 511 210 |
| 47 | intervention study.mp. | 63 409 |
| 48 | limit 47 to yr="2017 -Current" | 25 925 |
| 49 | exp Public Health Surveillance/ | 318 |
| 50 | limit 49 to yr="2017 -Current" | 312 |
| 51 | exp Health Surveys/ | 247 569 |
| 52 | limit 51 to yr="2017 -Current" | 63 056 |
| 53 | (Monitor* or Surveillance or Survey*).mp. [mp=title, abstract, heading word, drug trade name, original title, device manufacturer, drug manufacturer, device trade name, keyword heading word, floating subheading word, candidate term word] | 3 320 125 |
| 54 | limit 53 to yr="2017 -Current" | 919 107 |
| 55 | 32 or 34 or 36 or 38 or 40 or 42 or 44 or 46 or 48 or 50 or 52 or 54 | 3 054 621 |
| 56 | Humans/ | 16 243 254 |
| 57 | limit 56 to yr="2017 -Current" | 5 648 894 |
| 58 | 30 or 55 | 3 059 919 |
| 59 | 21 and 57 and 58 | 6119 |
| 60 | limit 59 to conference abstract status | 2500 |
| 61 | 59 not 60 | 3619 |

### Web of Science

| Search Terms | Number of Hits |
|---|---|
| TS="human herpesvirus 4" OR TI="human herpesvirus 4 | 3758 |
| TS="epstein barr virus" OR TI="epstein barr virus" | 15 005 |
| TS="epstein-barr virus" OR TI="epstein-barr virus" | 15 005 |
| TS=EBV OR TI=EBV | 12 367 |
| TS="eb virus" OR TI="eb virus" | 235 |
| TS=hhv-4 OR TI=hhv-4 | 43 |

| | |
|---|---|
| TS=hhv4 OR TI=hhv4 | 15 |
| TS="hhv 4" OR TI="hhv 4" | 43 |
| TS="glandular fever" OR TI="glandular fever" | 1129 |
| #1 OR #2 OR #3 OR #4 OR #5 OR #6 OR #7 OR #8 OR #9 OR #10 | 190 |
| TS=seropositiv* OR TI=seropositiv* | 18 992 |
| TS=seronegativ* OR TI=seronegativ* | 15 427 |
| TS=seroepidemiol* OR TI=seroepidemiol* | 6034 |
| TS=seroprevalence OR TI=seroprevalence | 9235 |
| #15 OR #14 OR #13 OR #12 | 15 105 |
| TS=epidemiol* OR TI=epidemiol* | 30 553 |
| TS="risk factor" OR TI="risk factor" | 840 624 |
| TS=cohort OR TI=cohort | 137 056 |
| TS="case control" OR TI="case control" | 550 762 |
| TS=case-control OR TI=case-control | 132 769 |
| TS="intervention study" OR TI="intervention study" | 6244 |
| TS="cross-sectional" OR TI="cross-sectional" | 671 787 |
| TS="clinical trial" OR TI="clinical trial" | 137 308 |
| TS=monitor* OR TI=monitor* | 1 552 904 |
| TS=surveillance OR TI=surveillance | 180 267 |
| TS=survey* OR TI=survey* | 1 000 155 |
| TS="Health Surveys" OR TI="Health Surveys" | 16 675 |
| TS="Public Health Surveillance" OR TI="Public Health Surveillance" | 5205 |
| #29 OR #28 OR #27 OR #26 OR #25 OR #24 OR #23 OR #22 OR #21 OR #20 OR #19 OR #18 OR #17 | 4 327 119 |
| #30 OR #16 | 4 336 631 |
| TS=human OR TI=human | 6 606 772 |
| #32 AND #31 AND #11 | 3761 |

**Supplementary Table 2 - Inclusion and Exclusion Criteria**

| | **Inclusion** | **Exclusion** |
|---|---|---|
| **Language** | Studies in any language | |
| **Time** | Studies from any time point after 2008—those published prior to 7 March 2017 taken from the previous review | Studies published prior to 2008, to ensure relevance for the modern day |
| **Study type** | Original research articles<br><br>Cross-sectional<br><br>Cohort<br><br>Case-control<br><br>Clinical trials (unless trial could have interfered with EBV acquisition) | Case reports<br><br>Retracted studies<br><br>Molecular biology studies |
| **Study population** | Human studies | Animal studies<br><br>Population those with EBV-associated disease only |
| **EBV serostatus** | Studies reported EBV serostatus by age | No comparison group (all participants were either seronegative or seropositive) |

EBV=Epstein-Barr virus.

### Supplementary Table 3. Quality assessment questions

| # | Question |
|---|---|
| 1 | Author |
| 2 | Was the study aiming to investigate seroprevalence by age? |
| 3 | Are the main outcomes to be measured clearly described in the introduction or methods section? |
| 4 | Are the main findings of the study clearly described? |
| 5 | Does the study provide estimates of the random variability in the data for the main outcomes? |
| 6 | Have the characteristics of patients lost to follow-up been described? |
| 7 | Was there potential for information bias in the ascertainment of the exposure? |
| 8 | Was there potential for differential or non-differential misclassification of the exposure? |
| 9 | Was there potential for information bias in ascertainment of the outcome? |
| 10 | Was there potential for differential or non-differential misclassification of the outcome? |
| 11 | Were the main outcome measures used accurate (valid and reliable)? |
| 12 | Were the patients in different age groups recruited from the same population? |
| 13 | Were study subjects in different age groups recruited over the same period of time? |
| 14 | Are the study results appropriately interpreted (e.g., in terms of the strength of their findings)? |

**Supplementary Table 4. Included studies**

| Author | Study population | Population summary | Number of participants | Study design | Country | WHO region | World Bank income level | Years in which study conducted | Age group | Female (%) | Diagnostic test | EBV antigen | Antibody type | Overall seroprevalence (%) |
|---|---|---|---|---|---|---|---|---|---|---|---|---|---|---|
| Abbas, H.[1] | Beta-thalassemia patients | 2- to 14-year-old patients | 212 | Case control | Pakistan | EMR | Mid | 2017–2018 | 2–14 y | 43·0 | Euroimmune, Germany ELISA kit | VCA | IgM | 5·0 |
| Adjei, A.[2] | HIV-seronegative blood donors and AIDS patients | Blood donors from 7 blood banks in Ghana | 3275 | Cross-sectional | Ghana | AFR | Low | 2001–2002 | 18–65 y | 21·4 | ELISA (Advanced Biotechnologies, Columbia, MD, USA) | Not stated | IgG | 77·6 |
| Alcantara-Neves, N.M.[3] | Children | Brazilian children's health cohorts | 1445 | Nested cross-sectional | Brazil | AMR | Mid | 2005 | 4–11 y | 46·4 | Not stated | Not stated | IgG | 88·5 |
| Altinas, J.[4] | Turkish adult population | Hospital outpatients and blood bank attendees | 500 | Retrospective cohort | Turkey | EUR | Mid | 2012 | 15–87 y | 42·6 | NovoTec NovaLisa | VCA | IgG | 96·4 |
| Balfour, H.H., Jr (a)[6] | US population | General population | 7516 | Continuous cross-sectional | USA | AMR | High | 2003–2010 | 6–19 y | 49·2 | EIAs (Diamedix, Miami, FL, USA) | VCA | IgG | Overall seroprevalence not given |
| Balfour, H.H, Jr (b)[5] | Students | Students from 3 freshman residence halls | 546 | Longitudinal | USA | AMR | High | 2006–2007 | 18–22 y | 59·7 | EIAs | VCA | IgG | 63·0 |
| Baroncelli, S.[7] | HIV-exposed uninfected infants | Infants born to HIV-infected mothers | 149 | Longitudinal | Malawi | AFR | Low | 2008–2011 | 6–24 m | 52·0 | ELISA MyBioSource Inc, San Diego, CA, USA | VCA | IgG | 88·7 |
| Beader, N.[8] | Croatian population | Different groups including healthy adults and children | 2022 | Prospective cohort | Croatia | EUR | High | 2015–2016 | 1–84 y | 91·4 | EBV VCA ELFA; VIDAS, Bimerieux, Marcy, l'Etoile, France) | VCA | IgM/IgG | 91·4 |

| Author | Study population | Population summary | Number of participants | Study design | Country | WHO region | World Bank income level | Years in which study conducted | Age group | Female (%) | Diagnostic test | EBV antigen | Antibody type | Overall seroprevalence (%) |
|---|---|---|---|---|---|---|---|---|---|---|---|---|---|---|
| Bistrom, M.[9] | Swedish MS patients and matched controls | Data combined from 6 Swedish biobanks | 670 cases; 670 controls | Nested case control | Sweden | EUR | High | 2012 | Not stated | 84·0 | Bead-based multiplex assay | EBNA1 trunc EBNA1 pep VCA p18 | IgG | 93·1 |
| Bulka, C.M.[10] | NHANES health survey of children and adults | Survey of health across the US | 8778 | Longitudinal | USA | AMR | High | 1999–2016[a] | 12–19 y | 47·3 | Commercial ELISA (Diamedix, Miami, FL, USA) | Not stated | IgG | 72·6 |
| Caputo, M.[11] | LISA study | Study of newborns with samples taken age 2 in Germany | 3094 | Longitudinal | Germany | EUR | High | 1997–1999 | 2 y | 47·5 | ELISAs | Not stated | IgG | 15·8 |
| Chang, C.M.[12] | Adults | Family members of those with NPC | 2393 | Cross-sectional | Taiwan | WPR | Mid | Not stated | >18y | 53·0 | Immunofluorescence assay, ELISA, enzyme neutralisation assay | Not stated | IgA | 49·3 |
| Chen, C.[13] | Survey | | 1411 | Cross-sectional | Taiwan | WPR | Mid | 2007 | All ages | 58·8 | ELISA (Euroimmun, Germany) | Not stated | IgG | 88·5 |
| Choi, A.[14] | University students | First-year university students | 198 | Longitudinal | USA | AMR | High | 2018 | 18–19 y | 63·1 | Commercially available EIA kits | EBNA1 VCA | IgG | 56·1 |
| Condon, L.M.[15] | Adults | Those with venous blood samples taken at an outpatient clinic | 705 | Cross-sectional | USA | AMR | High | 2011–2012 | 1–19y | 56·0 | EIA, (Diamedix, Miami, FL, USA) | VCA | IgG | Overall seroprevalence not given |
| Cui, J.[16] | Chinese population | Inpatients' and outpatients' samples for routine medical tests | 11 122 | Retrospective | China | WPR | Mid | 2013–2017 | 8 d–101 y | 48·6 | Commercially available ELISA kit (Euroimmun Medical Diagnostics, Lübek, Germany) | VCA EBNA1 EA-D | IgG, IgA, IgM | 94·5 |

| Author | Study population | Population summary | Number of participants | Study design | Country | WHO region | World Bank income level | Years in which study conducted | Age group | Female (%) | Diagnostic test | EBV antigen | Antibody type | Overall seroprevalence (%) |
|---|---|---|---|---|---|---|---|---|---|---|---|---|---|---|
| Cui, X.[17] | Chinese children | Chinese children admitted to West China Second University Hospital | 622 | Cross-sectional | China | WPR | Mid | 2011–2013 | 6 m–12 y | Not stated | Not stated | EBV-IgM | EBV-IgM | 42·0 |
| de Castro Alves, C.E[18] | General population | Outpatient attendees to a clinic in Presidente Figueiredo | 443 | Cross-sectional | Brazil | AMR | Mid | 2015–2016 | 1–80 y | 63·9 | Serion ELISA classic, SerionGmbH, Germany | VCA | IgG | 95·9 |
| Delaney, A.S.[19] | US population | | 2857 | Cross-sectional | USA | AMR | High | 2003–2004 | 6–19 y | 49·5 | Semi-quantitative EIA (Daimedix, Miami, FL, USA) | VCA | IgG | 70·0 |
| Dowd, J.B.[20] | NHANES | | 8417 | Cross-sectional | USA | AMR | High | 2003–2010 | 6–19 y | 48·0 | Semi-quantitative EIA (Daimedix, Miami, FL, USA) | Not stated | IgG | 66·5 |
| Dowd, J.B[21] | Whitehall II study of adults | Whitehall II epidemiological cohort | 400 | Longitudinal | UK | EUR | High | 2006–2008 | 53–76 y | 53·3 | Solid-phase enzyme immunoassay | EBNA1 | IgG | 63·3 |
| Du, J.L.[22] | Population from NPC endemic area | | 18 286 | Longitudinal | China | WPR | Middle | 1987–1992 | 30–59 y | 61·7 | Immunoenzymatic assay | Not stated | IgA | 7·0 |
| Durovic, B.[23] | Healthy blood donors | | 56 | Cross-sectional | Switzerland | EUR | High | | >60 y | 14·3 | Multiplex microscopy | Not stated | Not stated | 69·6 |
| Friborg, J.T.[24] | Sisimiut community cohort study | | 247 | Prospective cohort | Greenland (Denmark) | EUR | High | 1997–1998 | 0–4.5 y | 53·8 | ELISA (Novitec) | Not stated | IgG/IgM | 83·4 |
| Grut, V.[25] | Swedish MS registry and controls | Swedish microbiological biobank, and MS registry, cross linked | 670 cases; 670 controls | Nested case control | Sweden | EUR | High | Not stated | <20–39 y | 83·9 | Bead-based multiplex assay | EBNA1 trunc<br>EBNA1 pep<br>VCA p18 | Not stated | 93·1 |

| Author | Study population | Population summary | Number of participants | Study design | Country | WHO region | World Bank income level | Years in which study conducted | Age group | Female (%) | Diagnostic test | EBV antigen | Antibody type | Overall seroprevalence (%) |
|---|---|---|---|---|---|---|---|---|---|---|---|---|---|---|
| Hesla, H.[26] | Children from a birth cohort | Children | 157 | Prospective cohort | Sweden | EUR | High | 2004–2007 | 0–2 y | 49·0 | Immunofluorescence | VCA | IgG | Overall seroprevalence not given |
| Jansen, M.A.E.[27] | Generation R cohort | Children | 4464 | Prospective cohort | The Netherlands | EUR | High | 2002–2012 | 0–6 y | 48·3 | Enzyme immunoassay (Euroimmun) | Not stated | IgG | 51·1 |
| Jonker, I.[28] | TRAIL study of adolescents | Dutch adolescents | 2230 | Prospective cohort | The Netherlands | EUR | High | 2001 | 16 y | 53·4 | Immunoassay | Not stated | Not stated | 25·0 |
| Jonker, I.[29] | TRAIL study of adolescents | Dutch adolescents | 2230* | Prospective cohort | The Netherlands | EUR | High | 2001 | 16 y | 53·4 | Immunoassay | Not stated | Not stated | 24·1 |
| Kachuri, L.[30] | UKBiobank | Prospective cohort of UK adults aged 40–69 | 7948 | Prospective cohort | UK | EUR | High | 2006–2010 | 40–69 y | Not stated | ELISAs | VCA EBNA1 EA-D ZEBRA | IgG | 94·6 |
| Karachaliou, M.[31] | Rhea birth cohort | Cohort of children born to pregnant women | Serology available for 690 children | Longitudinal | Greece | EUR | High | 2007–2008* | 4 y | 47·6 | Multiplex serology | EBNA1 VCA ZEBRA EA-D | IgG | 53·0 |
| Karachaliou, M.[32] | Rhea birth cohort | Cohort of children born to pregnant women | Serology available for 690 children | Longitudinal | Greece | EUR | High | 2007–2008* | 4 y | 47·6 | Multiplex serology | EBNA1 VCA ZEBRA EA-D | IgG | 52·5 |
| Karachaliou, M.[33] | 81 out of 690 with serology | Children | 81/690 | Cohort | Greece | EUR | High | 2007–2008 | 0–4 y | Not stated | In-house fluorescence bead-based multiplex serology | Not stated | IgG | Overall seroprevalence not given |
| Khandaker, G.M.[34] | Children | N/A | 530 | Cross-sectional | UK | EUR | High | 1995–1996 | 4 y | 57·9 | Indirect immunofluorescence | Not stated | IgG | 25·3 |

| Author | Study population | Population summary | Number of participants | Study design | Country | WHO region | World Bank income level | Years in which study conducted | Age group | Female (%) | Diagnostic test | EBV antigen | Antibody type | Overall seroprevalence (%) |
|---|---|---|---|---|---|---|---|---|---|---|---|---|---|---|
| Kuri, A.[35] | PHA SEU samples | Samples across geographical areas in England from all age groups | 2366 | Cross-sectional | UK | EUR | High | 2016 | 1–25 y | 51·4 | ELISA (Abcam, cat no. ab108730) | VCA | IgG | 85·3 |
| Lasaviciute, G.[36] | Swedish birth cohort | Birth cohort with samples taken at 2, 5, and 10yo | 86 | Prospective birth cohort | Sweden | EUR | High | Not stated | 2–10 y | 57·0 | Previously described methods[b] | VCA | IgG | Overall seroprevalence not given |
| Levine, H.[37] | Israeli army recruits | All male | 1249 | Cross-sectional | Israel | EUR | High | 1994–2004 | 18–20 y | 0 | Enzyme-linked immunosorbent assay (novate, Germany) | Not stated | IgG | Overall seroprevalence not given |
| Looman, K.I.M.[38] | Generation R | Population based prospective cohort in the Netherlands | 3189 | Prospective cohort | The Netherlands | EUR | High | Not stated | 6 y | 51·6 | ELISA | VCA | IgG | 50·4 |
| Lu, Y.[39] | Adults in community dwellings | Subsample of the Singapore longitudinal ageing studies | 844 | Prospective cohort | Singapore | WPR | High | Not stated | 55+ y | 59·2 | ELISA (Virion) | EBNA | IgG | 97·3 |
| McDonald, J.A.[40] | Legacy girls' study | Girls aged 6–13 | 1068 | Prospective cohort | USA | AMR | High | 2011–2013 | 6–13 y | 100 | ELISA (Sera Quest International Inc., Miami, FL, USA) | Not stated | IgG, IgM | 60·8 |
| Minhas, V.[41] | Zambian infants | N/A | 677 | Cohort | Zambia | AFR | Low | 1998–2004 | 1 y | 48·7 | Not stated | Not stated | IgG | 25·2 |
| Ng, T.P.[42] | Community-dwelling older adults | Elderly adults | 2804 | Longitudinal | Singapore | WPR | High | 2003–2004 | 55 y | 39·5 | ELISA | VCA | not stated | 90·8 |

| Author | Study population | Population summary | Number of participants | Study design | Country | WHO region | World Bank income level | Years in which study conducted | Age group | Female (%) | Diagnostic test | EBV antigen | Antibody type | Overall seroprevalence (%) |
|---|---|---|---|---|---|---|---|---|---|---|---|---|---|---|
| Pembrey, L.[43] | Children | Born in Bradford infection and allergy study | 391 | Prospective cohort | UK | EUR | High | 2008–2012 | 0–2 y | 47·0 | Indirect chemiluminescence immunoassay | VCA | IgG | Overall seroprevalence not given |
| Pembrey, L.[44] | Children | Born in Bradford infection and allergy study | 391 | Prospective cohort | UK | EUR | High | 2008–2012 | 0–2 y | 47·0 | Indirect chemiluminescence immunoassay | VCA | IgG | 41·0 |
| Pembrey, L.[45] | Pregnant women | | 949 | Cross-sectional | UK | EUR | High | 2008–2009 | >20 y | 100 | Indirect chemiluminescence immunoassay | VCA | IgG | Overall seroprevalence not given |
| Qin, H.D.[46] | Healthy family members of NPC patients | | 3395 | Cross-sectional | China | WPR | Mid | 1999–2005 | All ages | 48·6 | Guangdon Zhonshan company | VCA | IgA | 27·0 |
| Rubicz, R.[47] | Mexican American families participating in San Antonio Family Heart Study | | 1227 | Cohort/Cross-sectional | USA | AMR | High | 1991–1995 | 15–94 y | 60·7 | Commerical ELISA | EBNA1 | IgG | 49·0 |
| Saghafian-Hedengren, S. [48] | 2-year-olds from previous maternity cohort; parent(s) have a history of allergy | | 51 | Cohort | Sweden | EUR | High | 1999–2002 | 2 y | 49·0 | In-house | Not stated | IgG | 49·0 |
| Savva, G.M.[49] | Older individuals without severe cognitive or physical impairments included in ESCR Healthy Aging Study | | 489 | Cohort | UK | EUR | High | Unknown | 65–94 y | 51·0 | Not stated | VCA | IgG | 88·8 |

| Author | Study population | Population summary | Number of participants | Study design | Country | WHO region | World Bank income level | Years in which study conducted | Age group | Female (%) | Diagnostic test | EBV antigen | Antibody type | Overall seroprevalence (%) |
|---|---|---|---|---|---|---|---|---|---|---|---|---|---|---|
| Setoh, J.[50] | Children | Stored samples from a women and children's hospital | 896 | Prospective cohort | Singapore | WPR | High | 2014–2015 | 1–19 y | 53·0 | Abbot Architect Assays | VCA EBNA | IgG, IgM | 68·3 |
| Sharifipour, S.[51] | Children and adults | Randomly selected samples from families living in districts of Tehran | 1220 | Cross-sectional | Iran | EMR | Mid | 2015–2019 | <1 m to ≥40 y | Not stated | ELISA (Behring's kit, Marburg, Germany) | EBV anti-VCA | IgG | 81·4 |
| Shen, G.P.[52] | Healthy individuals | | 755 | Cross-sectional | China | WPR | Mid | 2005–2007 | Not stated | 26·4 | Not stated | Not stated | IgA | 83·0 |
| Shi, T.[53] | Children | Children admitted to hospital with suspected EBV-associated illness | 3567 | Retrospective cohort | China | WPR | Mid | 2018–2020 | 0–16 y | 42·4 | Euroimmune, Germany ELISA kit | VCA EA EBNA1 | IgG, IgM | 74·6 |
| Simon, K.C.[54] | 10-year-olds from previous maternity cohort; parent(s) have a history of allergy | | 154 | Cohort | Sweden | EUR | High | 2002–2010 | 10 y | Not stated | Immunofluorescence | Not stated | IgG | 45·5 |
| Slyker, J.A.[55] | Extracted data for HIV-negative patients only, HIV-negative infants only followed for 12 months | Infants born to HIV-infected women | 125 | Prospective cohort | Kenya | AFR | Low | 1999–2003 (when the mothers were enrolled) | 0–2 y | 44·0 | PCR and ELISA (Wampole) | Not stated | IgG, IgM | 40·0 |
| Smith, N.A.[56] | Infants | Samples were taken from malaria holoendemic region in Western Kenya | 36 | Prospective cohort | Kenya | AFR | Not stated | Not stated | 0–11 y | 41·0 | Luminex assay (Immune-tech, New York, NY, USA) | VCA EBNA1 VCA Zta | IgG | 68·9 |

| **Author** | **Study population** | **Population summary** | **Number of participants** | **Study design** | **Country** | **WHO region** | **World Bank income level** | **Years in which study conducted** | **Age group** | **Female (%)** | **Diagnostic test** | **EBV antigen** | **Antibody type** | **Overall seroprevalence (%)** |
|---|---|---|---|---|---|---|---|---|---|---|---|---|---|---|
| Stowe, R.P.[57] | Hispanic families living close to petrochemical plants | | 1457 | Cohort (but for our purposes, cross-sectional) | USA | AMR | High | Not stated | Not stated | 54·8 | Not stated | Not stated | IgG | |
| Suntornlohanakul,R. [58] | Participants in study of universal HBV vaccination | 583 | Cross-sectional | Thailand | 2014 | SEAR | Mid | Not stated | 0–57 y | 50·4 | Not stated | VCA | IgG<br>IgM | 87·9 |
| Tang, Z.G.[59] | Adults | Not clear how they were recruited | 972 | Prospective cohort | China | WPR | Mid | 2017 | 19–91 | 36·5 | Shenzen New Industries | VCA<br>EA<br>EBNA1 | Not stated | Overall seroprevalence not given |
| Tiguman, G.M.B.[60] | Adults | Adults that had previously partaken in a population-based survey | 136 | Cross-sectional | Brazil | AMR | Mid | 2016 | 18<br>≥50y | 68·8 | Serion ELISA classic, SerionGmbH, Germany | Not stated | Not stated | 97·8 |
| Torniane-Holm, M.[61] | Adults | Health 2000 study of adults recruited between 2000–2001 | 6250 | Prospective cohort | Finland | EUR | High | 2000–2011 | ≥30y | 52·5 | Solid-phase immunoassay (IBL America) | VCA | IgG | 98·0 |
| Tuon, F.F.[62] | Tissue donors | Tissue donors with available serum samples after donation | 115 | Retrospective cohort | Brazil | AMR | Middle | 2016–2017 | 16–70 y | 38·9 | ELISA | VCA | IgG | 98·3 |
| van den Heuvel, D.[63] | Study of childhood, starting pre-birth | | 1079 | Cohort (but essentially cross-sectional for our purposes) | The Netherlands | EUR | High | Not stated | 5–7.9 y | Not stated | ELISA | VCA | IgG | 47·1 |

| Author | Study population | Population summary | Number of participants | Study design | Country | WHO region | World Bank income level | Years in which study conducted | Age group | Female (%) | Diagnostic test | EBV antigen | Antibody type | Overall seroprevalence (%) |
|---|---|---|---|---|---|---|---|---|---|---|---|---|---|---|
| Meel, E.R.[64] | Children | Generation R study | 3546 | Prospective cohort | The Netherlands | EUR | High | 2002–2006 | 5.6–7.2 y | 48·8 | ELISA (EUROIMMUN) | VCA | IgG | 50·4 |
| Vilibic-Cavlek, T.[65] | Haeomdialysis patients and healthy controls | Control group sent for routine testing | 150 | Case control | Croatia | EUR | High | 2013–2015 | 19–87 y | 42·7 | ELISA (EUROIMMUN) | VCA | IgG | Overall seroprevalence not given |
| Wang, G.C.[66] | Women | Women with difficulties in physical function dwelling in the community | 633 | Cohort (but cross-sectional for our purposes) | USA | AMR | High | 1992 onwards | 70–79 y | 100 | Gangway Biotech | Not stated | IgG | 72·7 |
| Wang, J.[67] | NHANES | Subset of neurodivergent children from NHANES | 2849 | Retrospective cohort | USA | AMR | High | 2003–2004 | 6–19 y | 48·7 | Enzyme immunoassay (Diamedix, Miami, FL, USA) | VCA | IgG | 69·6 |
| Winter, J.R.[68] | HSE data | Cross-sectional study from households in England | 732 | Cross-sectional | UK | EUR | High | 2002–2003 | 11–24 y | 50·3 | ELISA (EUROIMMUN) | VCA | IgG | 74·6 |
| Xiong, G.[69] | Separated by region (Guangzhou and Beijing - I combined seroprevalence) | Individuals who had undergone health and nutrition examinations | 1778 | Cross-sectional | China | WPR | Mid | 2012–2013 | 0–10 y | 48·9 | ELISA (EUROIMMUN) | Not stated | IgG, IgM | 80·1 |
| Xu, F.H.[70] | High-risk area for NPC | Healthy men in Guangdong | 3228 | Cross-sectional | China | WPR | Mid | 2005–2007 | 18–67 y | 0 | Immunoturbidimetry method (Hitachi) | Not stated | IgA | 17·5 |
| Zeeb, M.[71] | MEMO study | Elderly adults in Germany | 385 | Retrospective cohort | Germany | EUR | High | 1997–1998 | 65–83 | 46·0 | ELISA(Abbott-Diagnostics) | VCA | IgG | 99·0 |

AFR=African Region. AIDS=acquired immunodeficiency syndrome. AMR=Region of the Americas. d=day. EA-D=Early-D. EBNA=Epstein-Barr nuclear antigen. EBV=Epstein-Barr virus. EIAs=enzyme immunoassay. ELISA=enzyme-linked immunosorbent assay. EMR=Eastern Mediterranean Region. EUR, European Region. Mid=middle. HBV=Hepatitis B virus. HIV=human immunodeficiency virus. HSE=Health Survey England. LISA=Longitudinal and International Study of Adults. m=months. MS=multiple sclerosis. N/A=not applicable. NHANES=National

Health and Nutrition Examination Survey. NPC=nasopharyngeal carcinoma. PCR=polymerase chain reaction. PHA SEU=Public Health Seroepidemiology Unit. SEAR=South-East Asia Region. TRAIL=Tracking Adolescents' Individual Lives Survey. Trunc=truncated. UK=United Kingdom. US/USA= United States/United States of America. VCA=viral capsid antigen. WPR=Western Pacific Region. ZEBRA=Z EBV replication activator. neg=negative; y=years.

Footnotes: [a]This study excluded data from 2001-2002, [b] This study cited methods used in two other sources[72,73]

**Supplementary Table 5. Quality assessment of included studies using Downs and Black**

| Author | Aim seroprevalence by age? | Main outcomes clear? | Main findings clear? | Random variability? | Characteristics losses to follow-up? | Information bias-exposure? | Differential or non-differential misclassification-exposure? | Information bias-outcome? | Differential or non-differential misclassification-outcome? | Outcome measures accurate? | Different ages from same population? | Different ages, same time period? | Results correctly interpreted? |
|---|---|---|---|---|---|---|---|---|---|---|---|---|---|
| Abbas, H.[1] | No | Yes | Yes | No | N/A | No | No | No | No | Yes | Same population | Same population | Yes |
| Adjei, A.[2] | Yes | Yes | Yes | No | N/A | No | No | No | No | Yes | Same population | Same population | Yes |
| Alcantara-Neves, N.M.[3] | No | No | Yes | Yes | N/A | No | No | No | No | No | Same population | Same population | N/A |
| Altinas, J.[4] | No | Yes | Yes | No | N/A | No | No | No | No | Yes | Same population | Same population | Yes |
| Balfour, H.H., Jr (a)[6] | Yes | Yes | Yes | No | No | No | No | No | No | Yes | Same population | Same population | Yes |
| Balfour, H.H.,Jr (b)[5] | No | Yes | Yes | Yes | N/A | No | No | No | No | Yes | Same population | Same population | Yes |
| Baroncelli, S.[7] | No | Yes | Yes | Yes | No | No | No | No | No | Yes | Same population | Same population | Yes |
| Beader, N.[8] | Yes | Yes | Yes | No | N/A | No | No | No | No | Yes | Same population | Same population | Yes |
| Bistrom, M.[9] | Yes | Yes | Yes | Yes | N/A | No | No | No | No | Yes | Same population | Same population | Yes |
| Bulka, C.M.[10] | No | Yes | Yes | No | N/A | No | No | No | No | Yes | Same population | Same population | Yes |
| Caputo, M.[11] | Yes | Yes | Yes | No | Yes | No | No | No | No | Yes | Same population | Same population | Yes |
| Chang, C.M.[12] | No | No | Yes | Yes | N/A | No | No | No | No | Yes | Same population | Same population | Yes |
| Chen, C.Y.[13] | Yes | Yes | Yes | Yes | N/A | No | No | No | No | Yes | Same population | Same population | Yes |
| Choi, A.[14] | Yes | Yes | Yes | No | N/A | No | No | No | No | Yes | Same population | Same population | Yes |

| Author | Aim seroprevalence by age? | Main outcomes clear? | Main findings clear? | Random variability? | Characteristics losses to follow-up? | Information bias-exposure? | Differential or non-differential misclassification-exposure? | Information bias-outcome? | Differential or non-differential misclassification-outcome? | Outcome measures accurate? | Different ages from same population? | Different ages, same time period? | Results correctly interpreted? |
|---|---|---|---|---|---|---|---|---|---|---|---|---|---|
| Condon, L.M.[15] | Yes | Yes | Yes | Yes | N/A | No | No | No | No | Yes | Same population | Same population | Yes |
| Cui, J.[16] | No | Yes | Yes | Yes | N/A | No | No | No | No | Yes | Same population | Same population | Yes |
| Cui, Y.[74] | Yes | Yes | Yes | No | N/A | No | No | Yes | No | Yes | Same population | Same population | Yes |
| de Castro Alves, C.E.[18] | Yes | Yes | Yes | No | N/A | No | No | No | No | Yes | Same population | Same population | Yes |
| Delaney, A.S.[19] | No | Yes | Yes | Yes | N/A | No | No | No | No | Yes | Same population | Same population | Yes |
| Dowd, J.B.[21] | No | Yes | Yes | No | No | No | No | No | No | Yes | Same population | Same population | Yes |
| Dowd, J.B.[20] | Yes | Yes | Yes | Yes | N/A | No | No | No | No | Yes | Same population | Same population | Yes |
| Du, J.[75] | No | Yes | Yes | Yes | N/A | No | No | No | No | Yes | Same population | Same population | N/A |
| Durovic, B.[23] | No | No | No | No | N/A | No | No | No | No | No | Same population | Same population | N/A |
| Friborg, J.T.[24] | No | Yes | Yes | No | No | No | No | No | No | Yes | Same population | Same population | Yes |
| Grut, V.[25] | No | Yes | Yes | No | N/A | No | No | No | No | Yes | Same population | Same population | Yes |
| Hesla, H.M.[26] | No | No | Yes | No | No | No | No | No | No | Yes | Same population | Same population | N/A |
| Jansen, M.A.E.[27] | No | Yes | Yes | No | No | No | No | No | No | Yes | Same population | Same population | Yes |
| Jonker, I.[28] | No | Yes | Yes | No | No | No | No | Yes | No | Yes | Same population | Same population | Yes |

| Author | Aim seroprevalence by age? | Main outcomes clear? | Main findings clear? | Random variability? | Characteristics losses to follow-up? | Information bias-exposure? | Differential or non-differential misclassification-exposure? | Information bias-outcome? | Differential or non-differential misclassification-outcome? | Outcome measures accurate? | Different ages from same population? | Different ages, same time period? | Results correctly interpreted? |
|---|---|---|---|---|---|---|---|---|---|---|---|---|---|
| Jonker, I.[29] | No | Yes | Yes | No | No | No | No | Yes | No | No | Same population | Same population | Yes |
| Kachuri, L.[30] | No | Yes | Yes | No | No | No | No | Yes | No | No | Same population | Same population | Yes |
| Karachaliou, M.[31] | No | Yes | Yes | No | No | No | No | Yes | No | No | Same population | Same population | Yes |
| Karachaliou, M.[32] | No | Yes | Yes | No | No | No | No | Yes | No | No | Same population | Same population | Yes |
| Karachaliou, M.[33] | Yes | Yes | Yes | Yes | No | No | No | No | No | No | Same population | Same population | Yes |
| Khandaker, G.M.[34] | No | No | Yes | No | N/A | No | No | No | No | Yes | Same population | Same population | N/A |
| Kuri, A.[35] | Yes | Yes | Yes | No | N/A | No | No | No | No | Yes | Same population | Same population | Yes |
| Lasaviciute, G.[36] | No | Yes | Yes | No | N/A | No | No | Yes | No | No | Same population | Same population | Yes |
| Levine, H.[37] | No | Yes | Yes | Yes | N/A | No | No | No | No | Yes | Same population | Same population | Yes |
| Looman, I.M.[38] | No | Yes | Yes | No | N/A | No | No | No | No | Yes | Same population | Same population | Yes |
| Lu, Y.[76] | No | Yes | Yes | No | N/A | No | No | Yes | No | No | Same population | Same population | Yes |
| McDonald, J.A [40] | No | Yes | Yes | No | N/A | No | No | No | No | Yes | Same population | Same population | Yes |
| Minhas, V.[41] | Yes | No | Yes | No | No | No | No | No | No | Yes | Same population | Same population | Yes |
| Ng, T.P. [42] | Yes | Yes | Yes | No | N/A | No | No | No | No | Yes | Same population | Same population | Yes |

| Author | Aim seroprevalence by age? | Main outcomes clear? | Main findings clear? | Random variability? | Characteristics losses to follow-up? | Information bias-exposure? | Differential or non-differential misclassification-exposure? | Information bias-outcome? | Differential or non-differential misclassification-outcome? | Outcome measures accurate? | Different ages from same population? | Different ages, same time period? | Results correctly interpreted? |
|---|---|---|---|---|---|---|---|---|---|---|---|---|---|
| Pembrey, L.[44] | Yes | Yes | Yes | No | N/A | No | No | No | No | Yes | Same population | Same population | Yes |
| Pembrey, L.[43] | Yes | Yes | Yes | No | N/A | No | No | No | No | Yes | Same population | Same population | Yes |
| Pembrey, L.[45] | Yes | No | Yes | No | N/A | No | No | No | No | Yes | Same population | Same population | Yes |
| Qin, H.D.[46] | No | No | Yes | No | N/A | No | No | No | No | Yes | Same population | Same population | Yes |
| Rubicz, R.[47] | Yes | Yes | Yes | No | N/A | No | No | No | No | Yes | Same population | Same population | Yes |
| Saghafian-Hedengren, S.[48] | No | Yes | Yes | No | N/A | No | No | No | No | No | Same population | Same population | N/A |
| Savva, G.M.[49] | No | Yes | Yes | No | N/A | No | No | No | No | Yes | Same population | Same population | N/A |
| Setoh, J.W.S.[50] | Yes | Yes | No | No | N/A | No | No | No | No | Yes | Same population | Same population | Yes |
| Sharifipour, S.[51] | Yes | Yes | No | No | N/A | No | No | No | No | Yes | Same population | Same population | Yes |
| Shen, G.P.[52] | No | Yes | Yes | No | N/A | No | No | No | No | Yes | Same population | Same population | Yes |
| Shi, T.[53] | Yes | Yes | Yes | No | N/A | No | No | No | No | Yes | Same population | Same population | Yes |
| Simon, K.C.[54] | Yes | No | Yes | No | No | No | No | No | No | Yes | Same population | Same population | Yes |
| Slyker, J.A.[55] | No | No | Yes | Yes | No | No | No | No | No | Yes | Same population | Same population | Yes |
| Smith, N. A[56] | No | No | Yes | No | N/A | No | No | No | No | Yes | Same population | Same population | Yes |

| Author | Aim seroprevalence by age? | Main outcomes clear? | Main findings clear? | Random variability? | Characteristics losses to follow-up? | Information bias-exposure? | Differential or non-differential misclassification-exposure? | Information bias-outcome? | Differential or non-differential misclassification-outcome? | Outcome measures accurate? | Different ages from same population? | Different ages, same time period? | Results correctly interpreted? |
|---|---|---|---|---|---|---|---|---|---|---|---|---|---|
| Stowe, R.P.[57] | No | No | Yes | Yes | N/A | No | No | No | No | Yes | Same population | Same population | Yes |
| Suntornlohanakul, R.[58] | Yes | No | Yes | Yes | N/A | No | No | No | No | Yes | Same population | Same population | Yes |
| Tang, Z.G.[59] | Yes | Yes | Yes | No | N/A | No | No | No | No | Yes | Same population | Same population | Yes |
| Tiguman, G. M.B.[60] | Yes | Yes | Yes | Yes | N/A | No | No | No | No | Yes | Same population | Same population | Yes |
| Torniainen-Holm, M.[61] | No | Yes | Yes | No | No | No | No | No | No | Yes | Same population | Same population | Yes |
| Tuon, F.F.[62] | Yes | Yes | Yes | No | N/A | No | No | No | No | Yes | Same population | Same population | Yes |
| van den Heuvel, D.[63] | No | Yes | Yes | No | N/A | No | No | No | No | Yes | Same population | Same population | N/A |
| van Meel, E. R[64] | No | Yes | Yes | No | No | No | No | Yes | No | No | Same population | Same population | Yes |
| Vilibic-Cavlek, T.[65] | Yes | Yes | Yes | No | N/A | No | No | No | No | Yes | Same population | Same population | Yes |
| Wang, J. [67] | Yes | Yes | Yes | No | N/A | No | No | No | No | Yes | Same population | Same population | N/A |
| Wang, G.C.[66] | No | Yes | Yes | No | N/A | No | No | No | No | Yes | Same population | Same population | Yes |
| Winter, J.R.[68] | Yes | Yes | Yes | Yes | N/A | No | No | No | No | Yes | Same population | Same population | Yes |
| Xiong, G.[69] | No | Yes | Yes | No | N/A | No | No | No | No | Yes | Same population | Same population | Yes |
| Xu, F.H.[70] | No | No | Yes | Yes | N/A | No | No | No | No | Yes | Same population | Same population | Yes |

| Author | Aim seroprevalence by age? | Main outcomes clear? | Main findings clear? | Random variability? | Characteristics losses to follow-up? | Information bias-exposure? | Differential or non-differential misclassification-exposure? | Information bias-outcome? | Differential or non-differential misclassification-outcome? | Outcome measures accurate? | Different ages from same population? | Different ages, same time period? | Results correctly interpreted? |
|---|---|---|---|---|---|---|---|---|---|---|---|---|---|
| Zeeb, M.[71] | No | Yes | Yes | No | N/A | No | No | No | No | Yes | Same population | Same population | Yes |

**Supplementary Figure 1.** Study-specific seroprevalence (proportion) plotted for all included studies[8,9,26,32,35,36,43,45,49,61,64,65,68,71] Meta-analysed across the following age groups: A) 0–4 years, B) 5–9 years, C) 10–14 years, D) 20–29 years, E) 30–39 years, and F) ≥40 years. Multiple data points were used from the same study if the study had a more than one seroprevalence measurement within the meta-analysis age-range. CI=confidence interval. ES=effect size.

**A)**

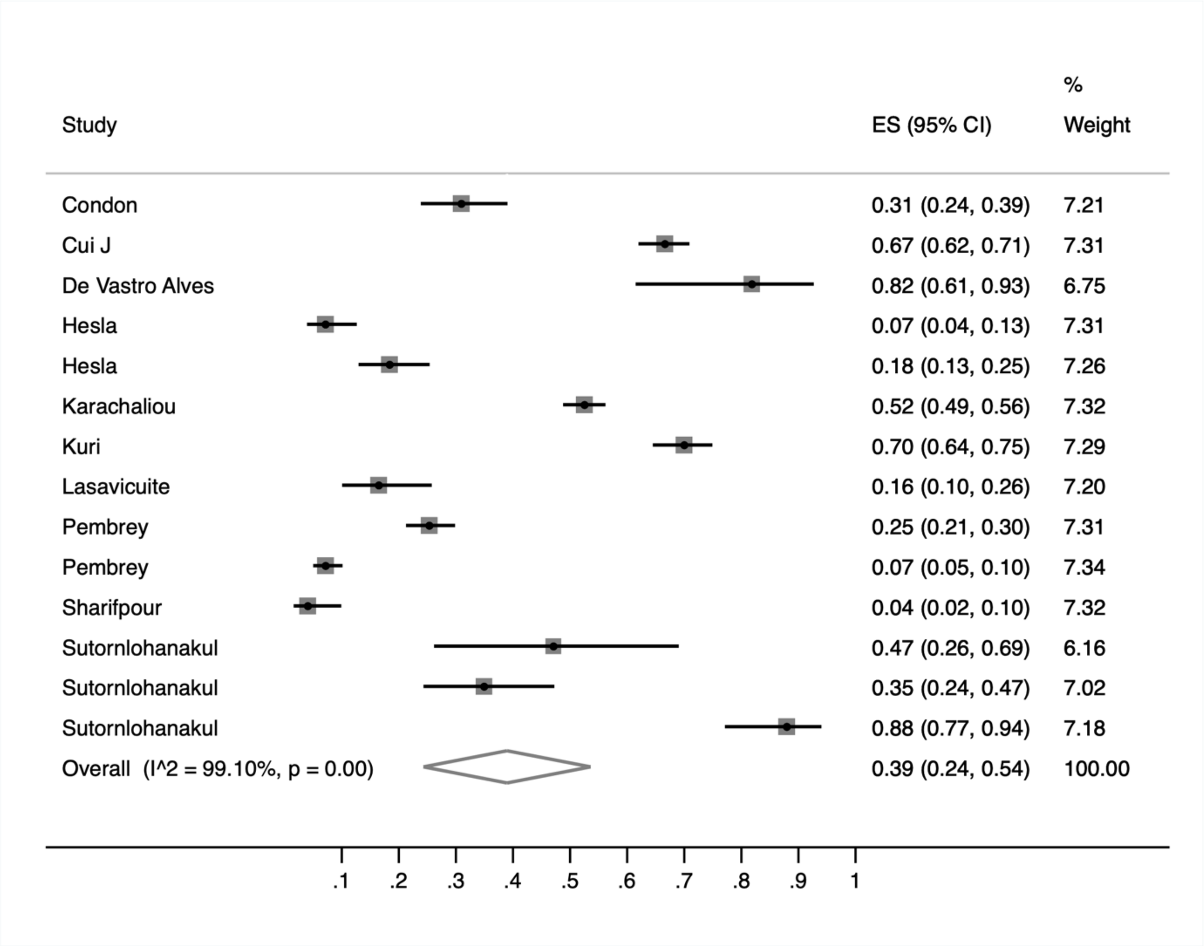

B)

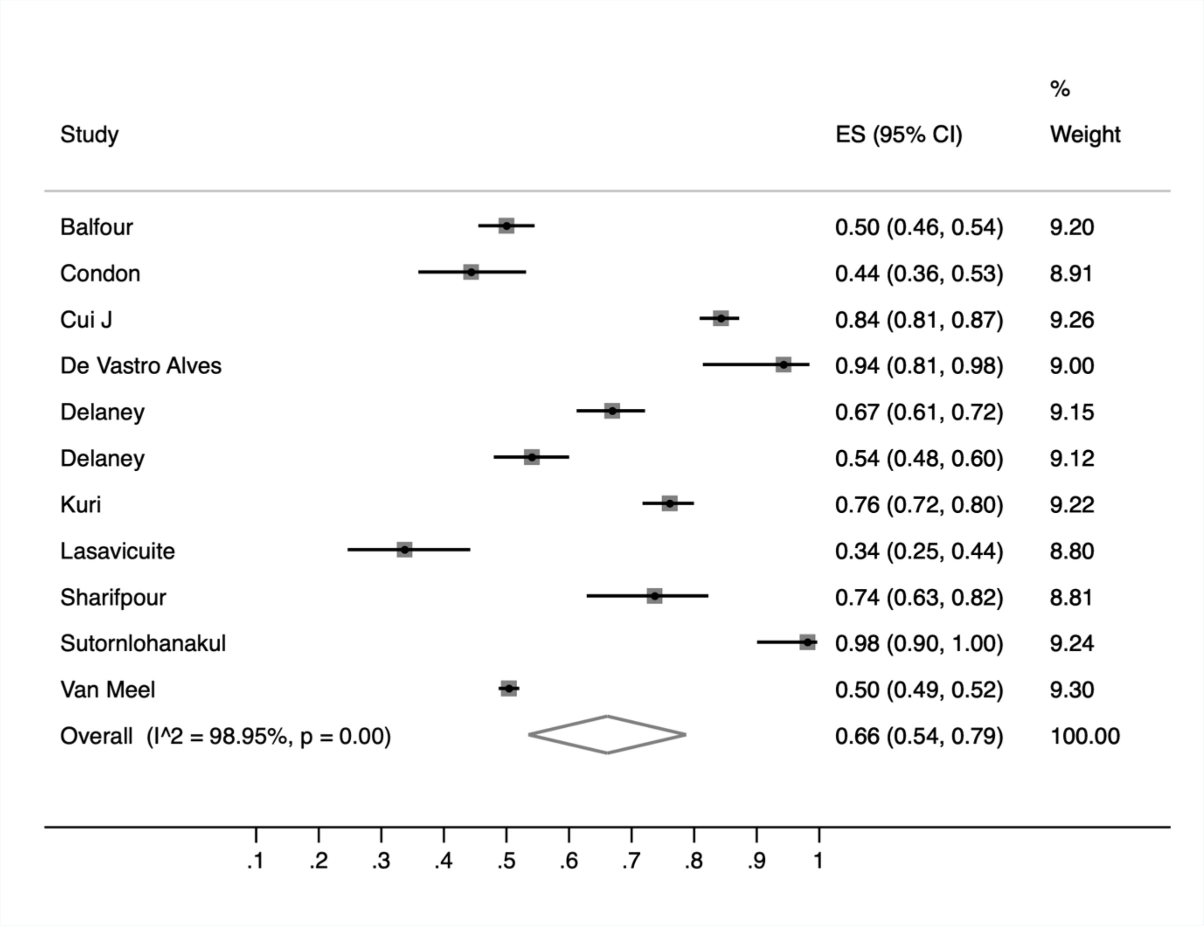
Study
ES (95% CI)
%
Weight
Balfour
0.50 (0.46, 0.54)
9.20
Condon
0.44 (0.36, 0.53)
8.91
Cui J
0.84 (0.81, 0.87)
9.26
De Vastro Alves
0.94 (0.81, 0.98)
9.00
Delaney
0.67 (0.61, 0.72)
9.15
Delaney
0.54 (0.48, 0.60)
9.12
Kuri
0.76 (0.72, 0.80)
9.22
Lasavicuite
0.34 (0.25, 0.44)
8.80
Sharifpour
0.74 (0.63, 0.82)
8.81
Sutornlohanakul
0.98 (0.90, 1.00)
9.24
Van Meel
0.50 (0.49, 0.52)
9.30
Overall (I^2 = 98.95%, p = 0.00)
0.66 (0.54, 0.79)
100.00
.1
.2
.3
.4
.5
.6
.7
.8
.9
1

**C)**

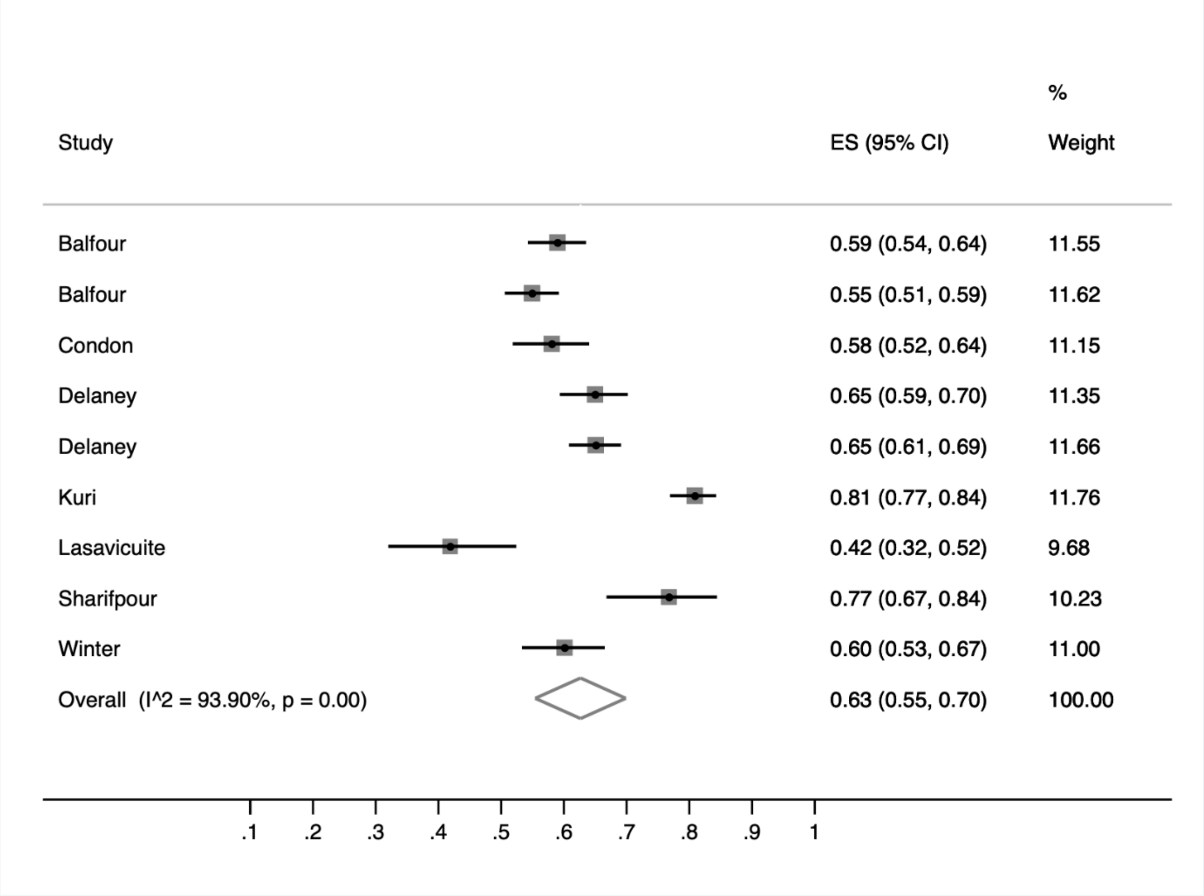
%
Study
ES (95% CI)
Weight
Balfour
0.59 (0.54, 0.64)
11.55
Balfour
0.55 (0.51, 0.59)
11.62
Condon
0.58 (0.52, 0.64)
11.15
Delaney
0.65 (0.59, 0.70)
11.35
Delaney
0.65 (0.61, 0.69)
11.66
Kuri
0.81 (0.77, 0.84)
11.76
Lasavicuite
0.42 (0.32, 0.52)
9.68
Sharifpour
0.77 (0.67, 0.84)
10.23
Winter
0.60 (0.53, 0.67)
11.00
Overall (I^2 = 93.90%, p = 0.00)
0.63 (0.55, 0.70)
100.00
.1
.2
.3
.4
.5
.6
.7
.8
.9
1

D

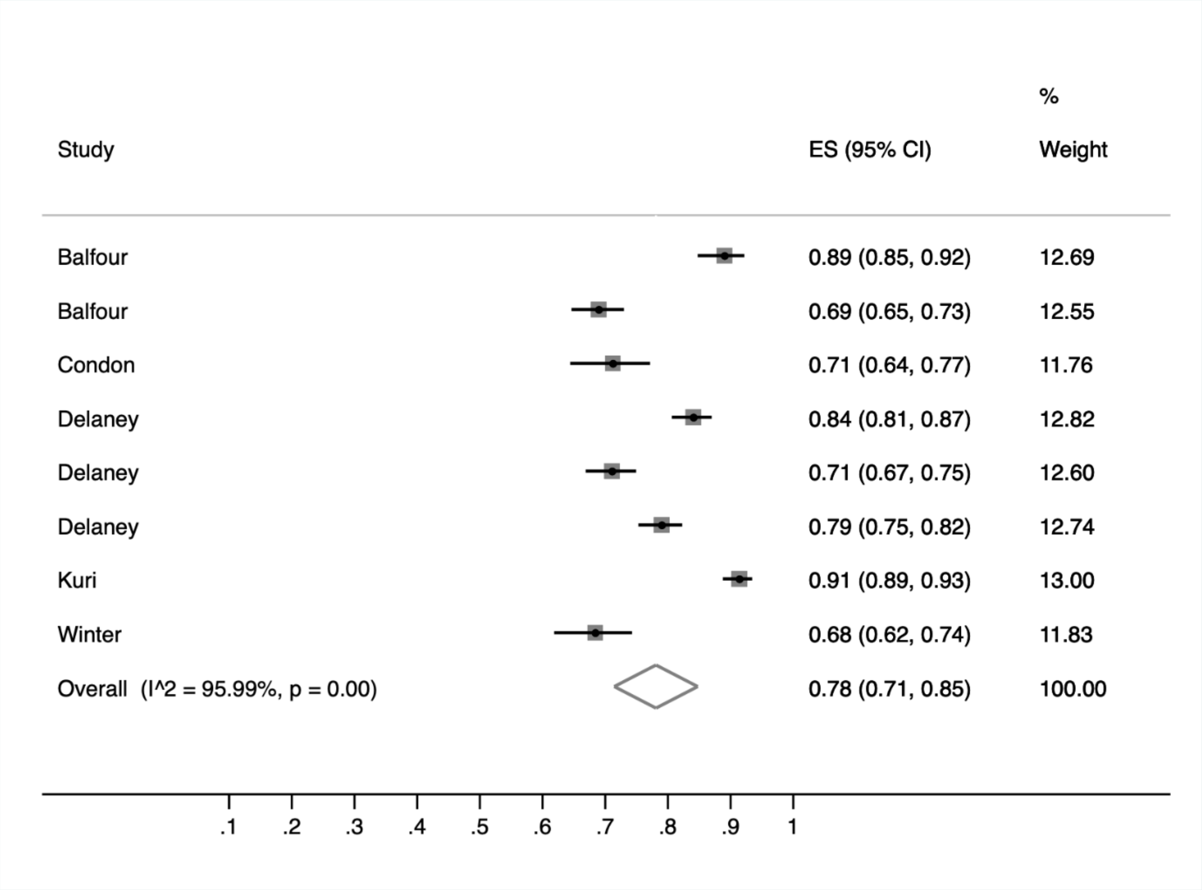

Study
ES (95% CI)
%
Weight
Balfour
0.89 (0.85, 0.92)
12.69
Balfour
0.69 (0.65, 0.73)
12.55
Condon
0.71 (0.64, 0.77)
11.76
Delaney
0.84 (0.81, 0.87)
12.82
Delaney
0.71 (0.67, 0.75)
12.60
Delaney
0.79 (0.75, 0.82)
12.74
Kuri
0.91 (0.89, 0.93)
13.00
Winter
0.68 (0.62, 0.74)
11.83
Overall (I^2 = 95.99%, p = 0.00)
0.78 (0.71, 0.85)
100.00
.1
.2
.3
.4
.5
.6
.7
.8
.9
1

E)

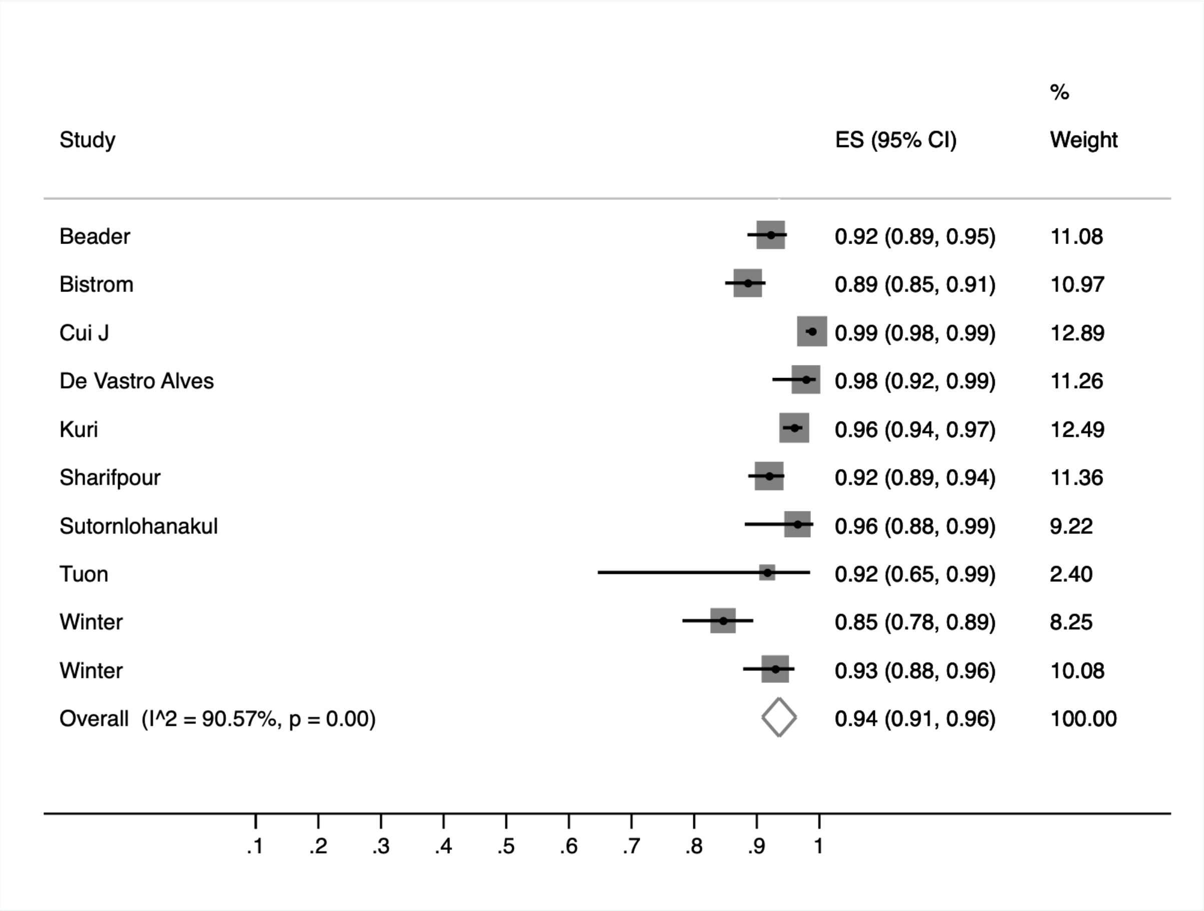
Study
ES (95% CI)
%
Weight
Beader
0.92 (0.89, 0.95)
11.08
Bistrom
0.89 (0.85, 0.91)
10.97
Cui J
0.99 (0.98, 0.99)
12.89
De Vastro Alves
0.98 (0.92, 0.99)
11.26
Kuri
0.96 (0.94, 0.97)
12.49
Sharifpour
0.92 (0.89, 0.94)
11.36
Sutornlohanakul
0.96 (0.88, 0.99)
9.22
Tuon
0.92 (0.65, 0.99)
2.40
Winter
0.85 (0.78, 0.89)
8.25
Winter
0.93 (0.88, 0.96)
10.08
Overall (I^2 = 90.57%, p = 0.00)
0.94 (0.91, 0.96)
100.00
.1
.2
.3
.4
.5
.6
.7
.8
.9
1

**F)**

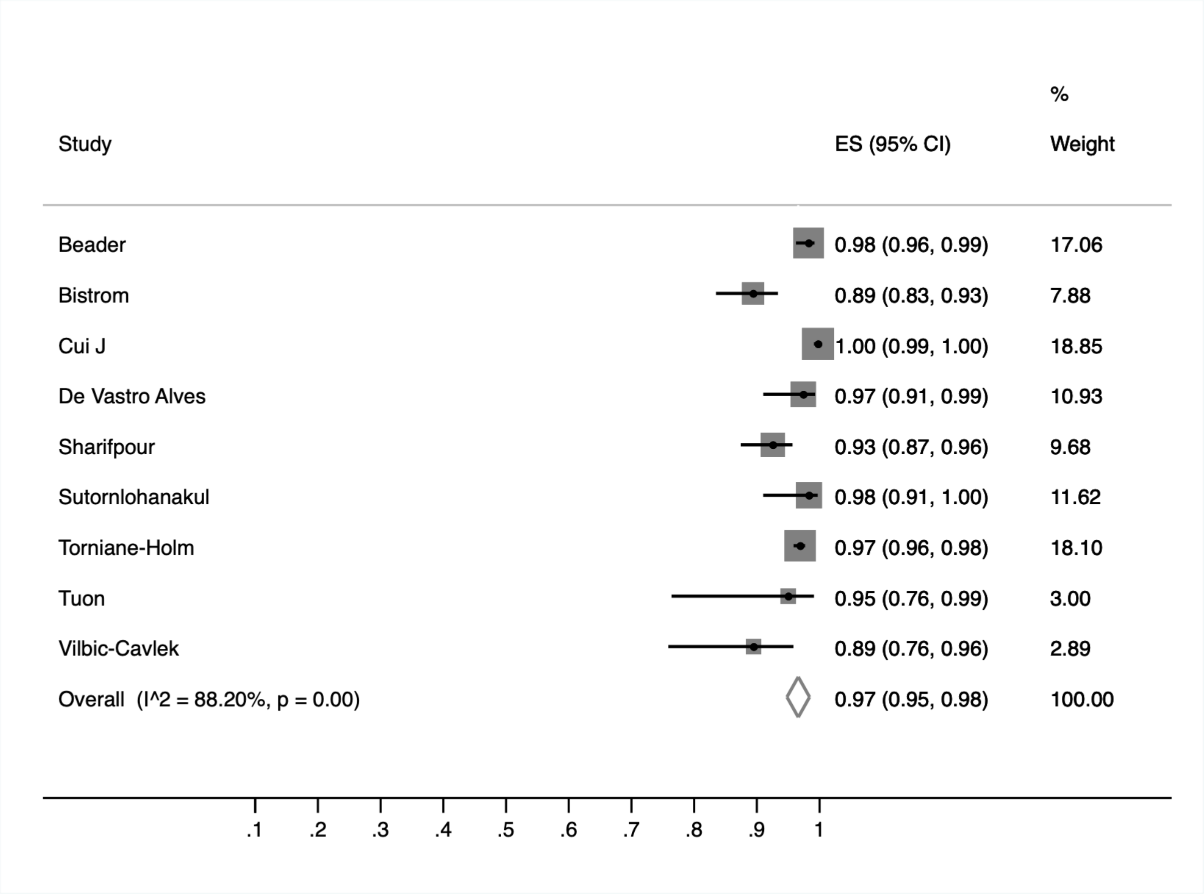
Study
ES (95% CI)
%
Weight
Beader
0.98 (0.96, 0.99)
17.06
Bistrom
0.89 (0.83, 0.93)
7.88
Cui J
1.00 (0.99, 1.00)
18.85
De Vastro Alves
0.97 (0.91, 0.99)
10.93
Sharifpour
0.93 (0.87, 0.96)
9.68
Sutornlohanakul
0.98 (0.91, 1.00)
11.62
Torniane-Holm
0.97 (0.96, 0.98)
18.10
Tuon
0.95 (0.76, 0.99)
3.00
Vilbic-Cavlek
0.89 (0.76, 0.96)
2.89
Overall (I^2 = 88.20%, p = 0.00)
0.97 (0.95, 0.98)
100.00
.1
.2
.3
.4
.5
.6
.7
.8
.9
1

G)

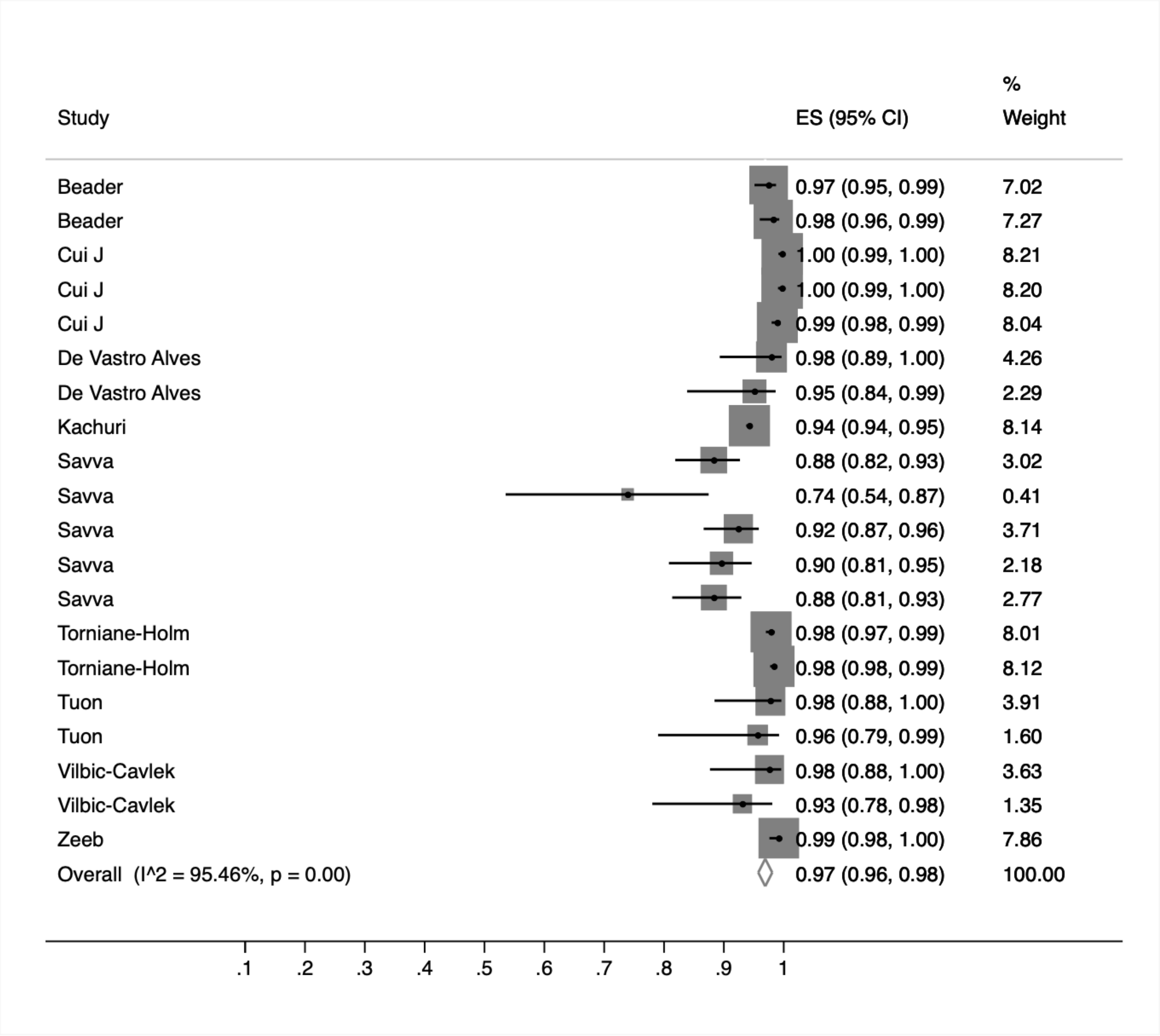
Study
ES (95% CI)
% Weight
Beader 0.97 (0.95, 0.99) 7.02
Beader 0.98 (0.96, 0.99) 7.27
Cui J 1.00 (0.99, 1.00) 8.21
Cui J 1.00 (0.99, 1.00) 8.20
Cui J 0.99 (0.98, 0.99) 8.04
De Vastro Alves 0.98 (0.89, 1.00) 4.26
De Vastro Alves 0.95 (0.84, 0.99) 2.29
Kachuri 0.94 (0.94, 0.95) 8.14
Savva 0.88 (0.82, 0.93) 3.02
Savva 0.74 (0.54, 0.87) 0.41
Savva 0.92 (0.87, 0.96) 3.71
Savva 0.90 (0.81, 0.95) 2.18
Savva 0.88 (0.81, 0.93) 2.77
Torniane-Holm 0.98 (0.97, 0.99) 8.01
Torniane-Holm 0.98 (0.98, 0.99) 8.12
Tuon 0.98 (0.88, 1.00) 3.91
Tuon 0.96 (0.79, 0.99) 1.60
Vilbic-Cavlek 0.98 (0.88, 1.00) 3.63
Vilbic-Cavlek 0.93 (0.78, 0.98) 1.35
Zeeb 0.99 (0.98, 1.00) 7.86
Overall (I^2 = 95.46%, p = 0.00) 0.97 (0.96, 0.98) 100.00
.1 .2 .3 .4 .5 .6 .7 .8 .9 1

**Supplementary Figure 2. Funnel plots to assess publication bias.** Each plot displays the standard error of the effect size (SE[ES]) on the vertical axis against the effect size (ES) on the horizontal axis. The dashed lines represent pseudo 95% confidence limits. The solid black line represents the summary effect size from the meta-analysis.Symmetry suggests absence of publication bias, while asymmetry may indicate small-study effects. Funnel plots are shown for the following age groups: A) 0–4 years, B) 5–9 years, C) 10–14 years, D) 15–19 years, E) 20–29 years, F) 30–39 years, G) ≥40 years

**A)**

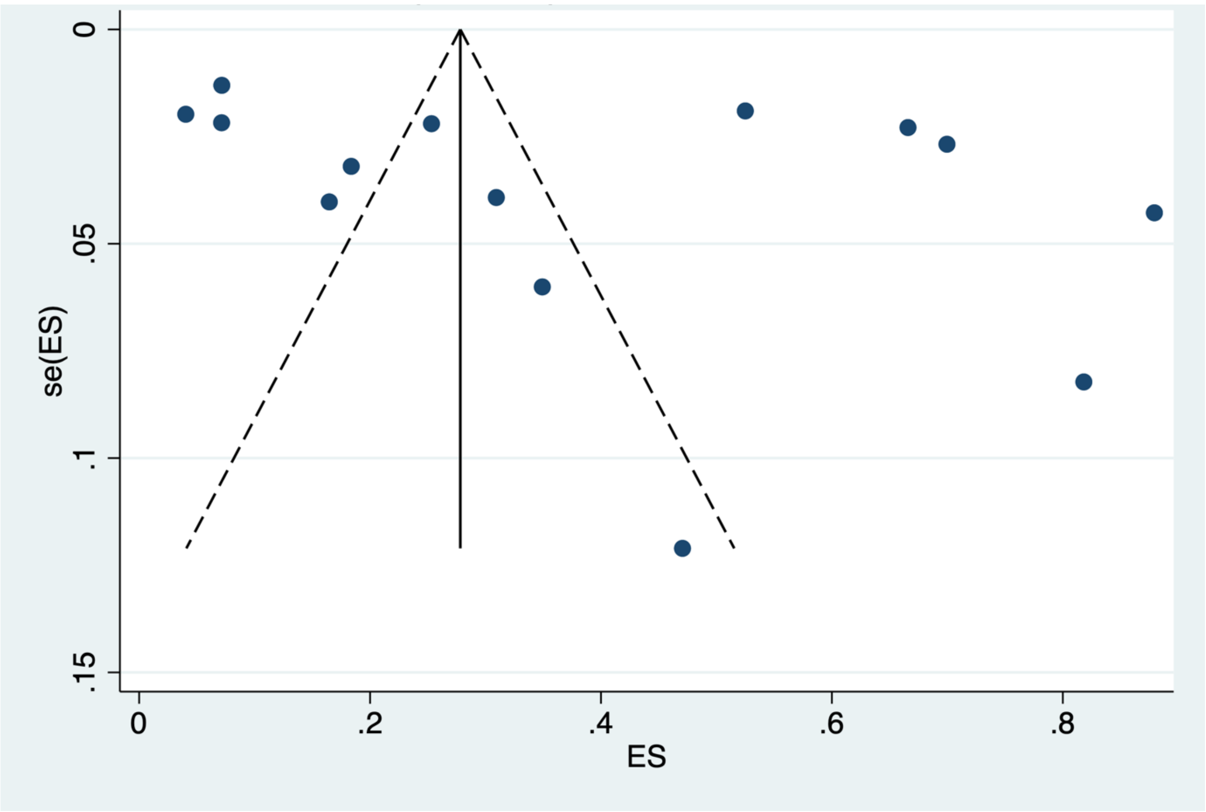

B)

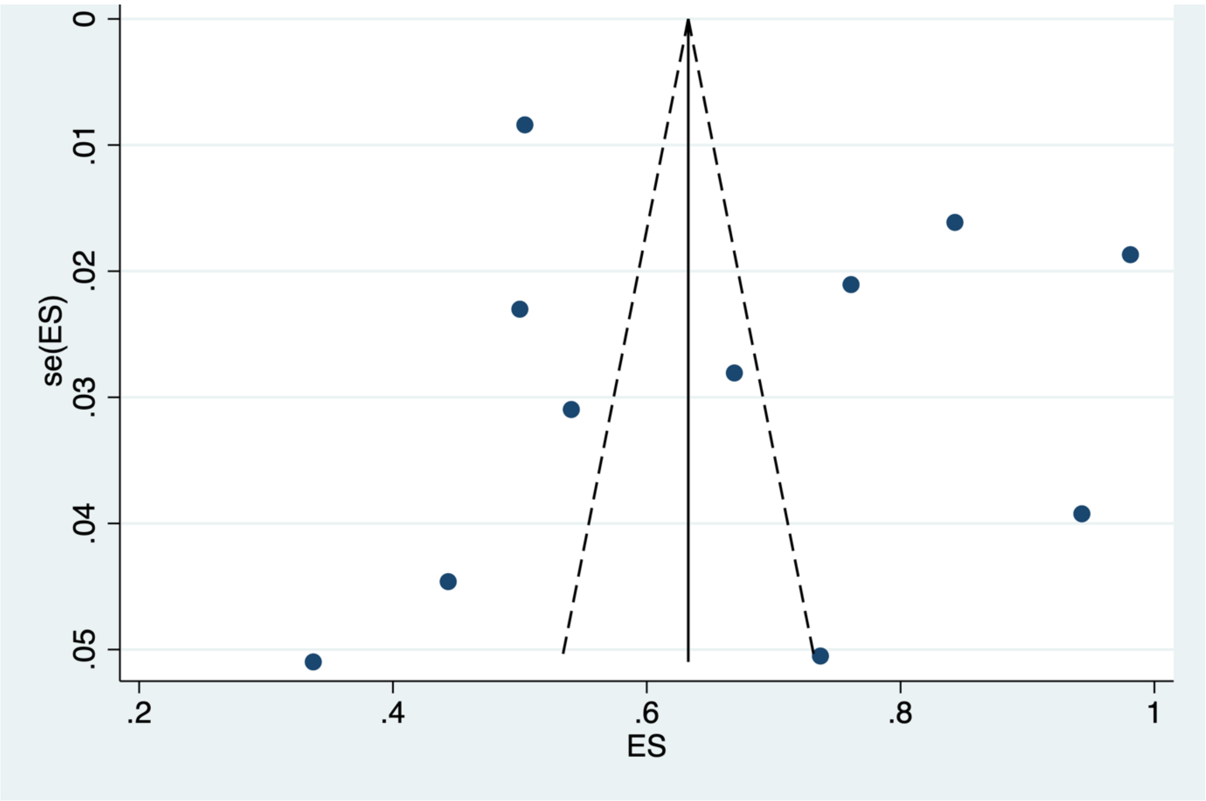


C)

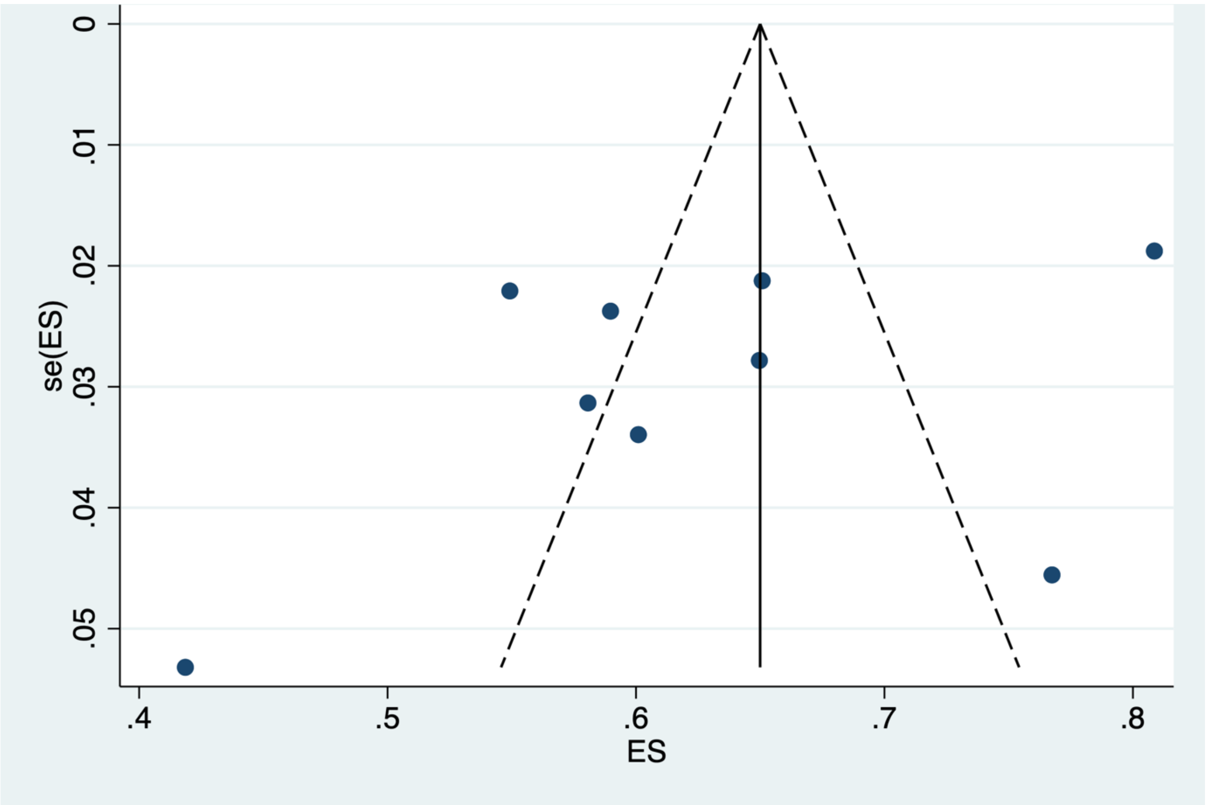

D)

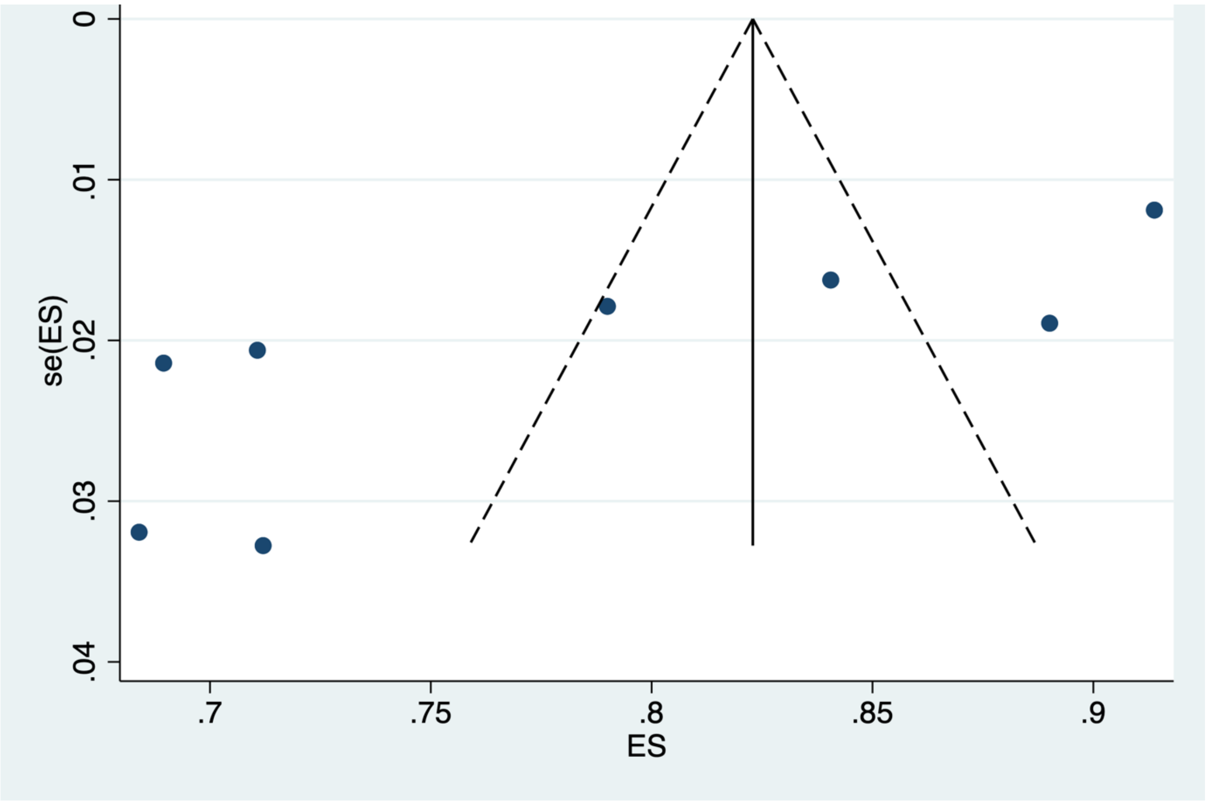


E)

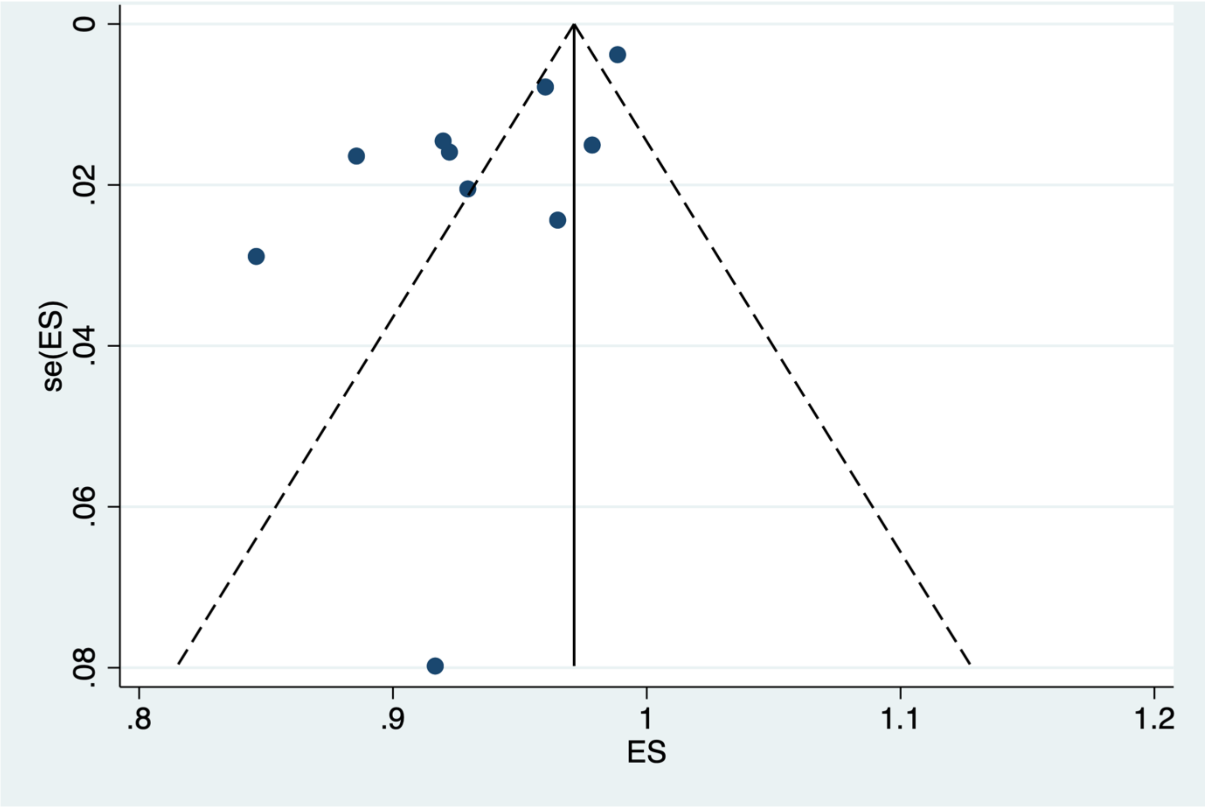

F)

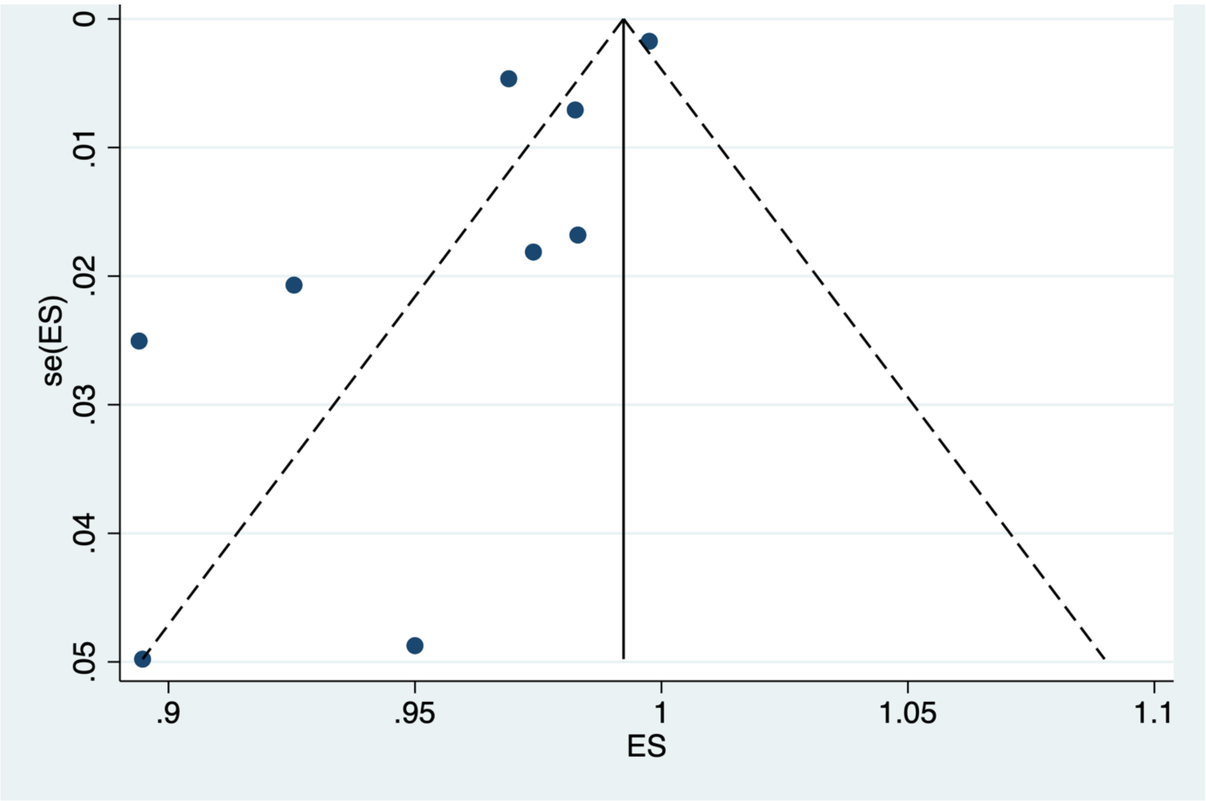
se(ES)
0
.01
.02
.03
.04
.05
.9
.95
1
1.05
1.1
ES

G)

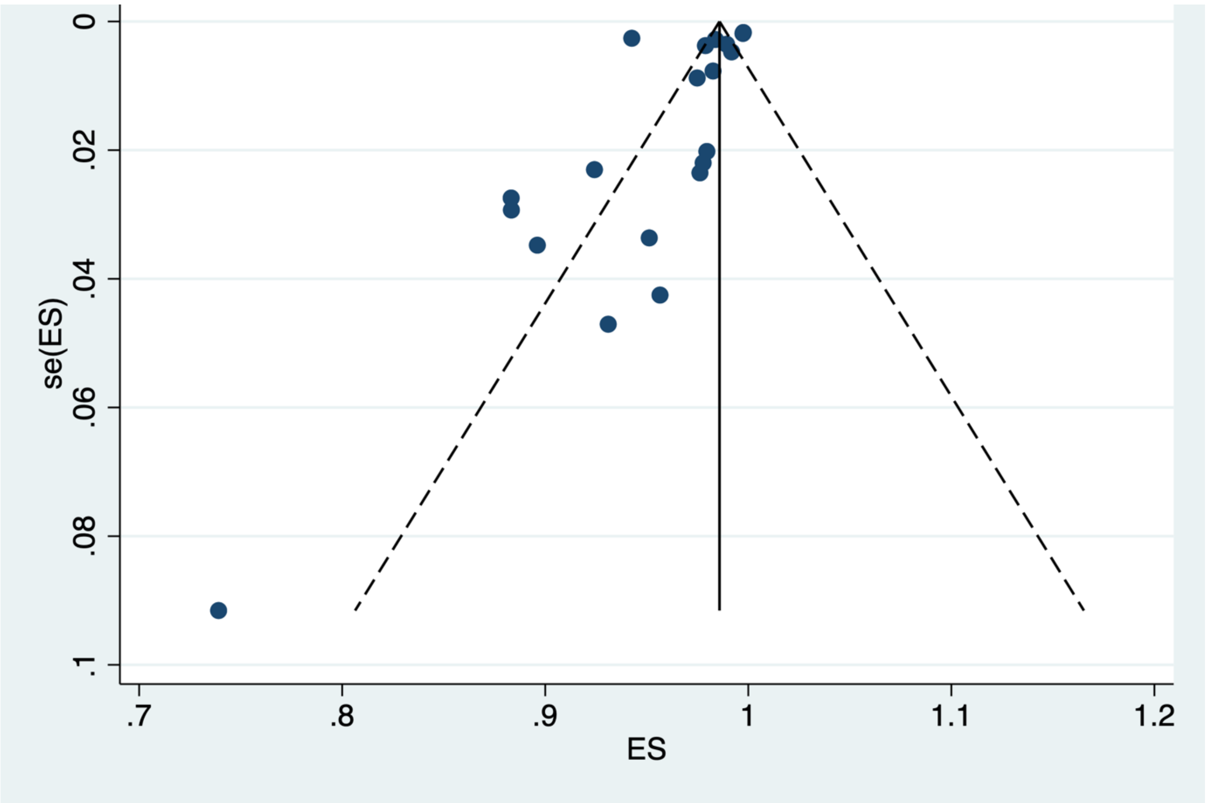
se(ES)
0
.02
.04
.06
.08
.1
.7
.8
.9
1
1.1
1.2
ES

**Supplementary Table 6. Egger's regression test for funnel plot asymmetry.**

This table presents the results of Egger's regression test for funnel plot asymmetry, conducted in age groups with ≥10 studies. The intercept, its confidence interval (CI) and associated p-value are presented. A non-zero intercept suggests asymmetry consistent with publication bias..

| **Age Group** | **Number of Studies** | **Intercept** | **95% CI** | **p-value (Egger)** |
|---|---|---|---|---|
| **0–4** | 14 | 8.90 | -4.08 to 21.88 | 0.161 |
| **5–9** | 11 | 4.80 | -7.90 to 17.49 | 0.415 |
| **20–29** | 10 | -3.69 | -6.12 to -1.27 | 0.008 |
| **>40** | 20 | -2.27 | -5.13 to 0.59 | 0.113 |

**Supplementary Figure 3. Seroprevalence by age (years).** A) African Region[2,7,41,55], B) Ghana, C) Kenya, D) Malawi, and E) Zambia. Horizontal lines represent the age groups covered by a given data point

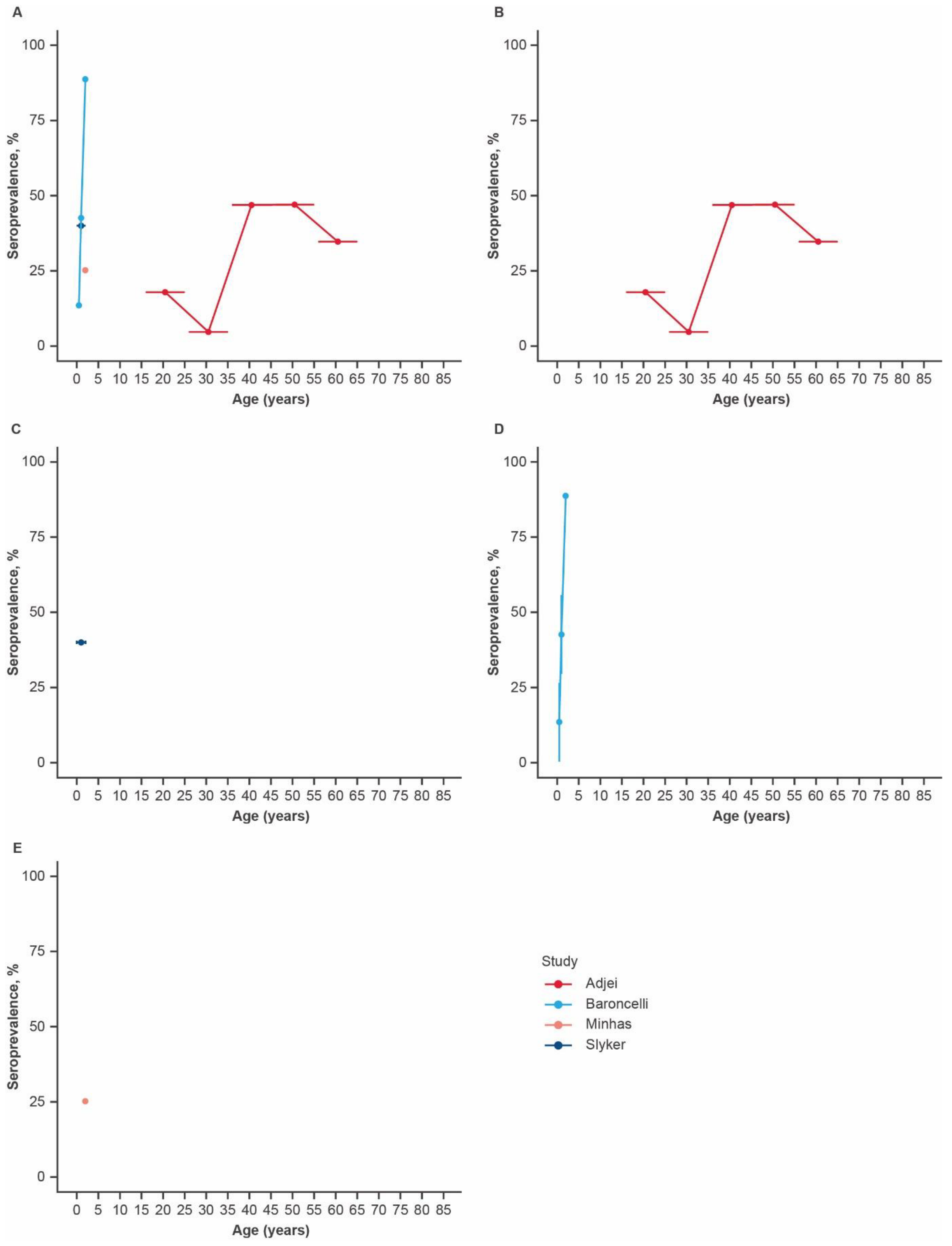

**Supplementary Figure 4. Seroprevalence by age (years).** A) European Region[4,8,9,26,27,30,33,35,36,38,43,45,49,61,63-65,68,71], B) Croatia, C) Finland, D) Germany, E) Greece, F) The Netherlands, G) Sweden, H) Turkey, and I) UK. Horizontal lines represent the age groups covered by a given data point.

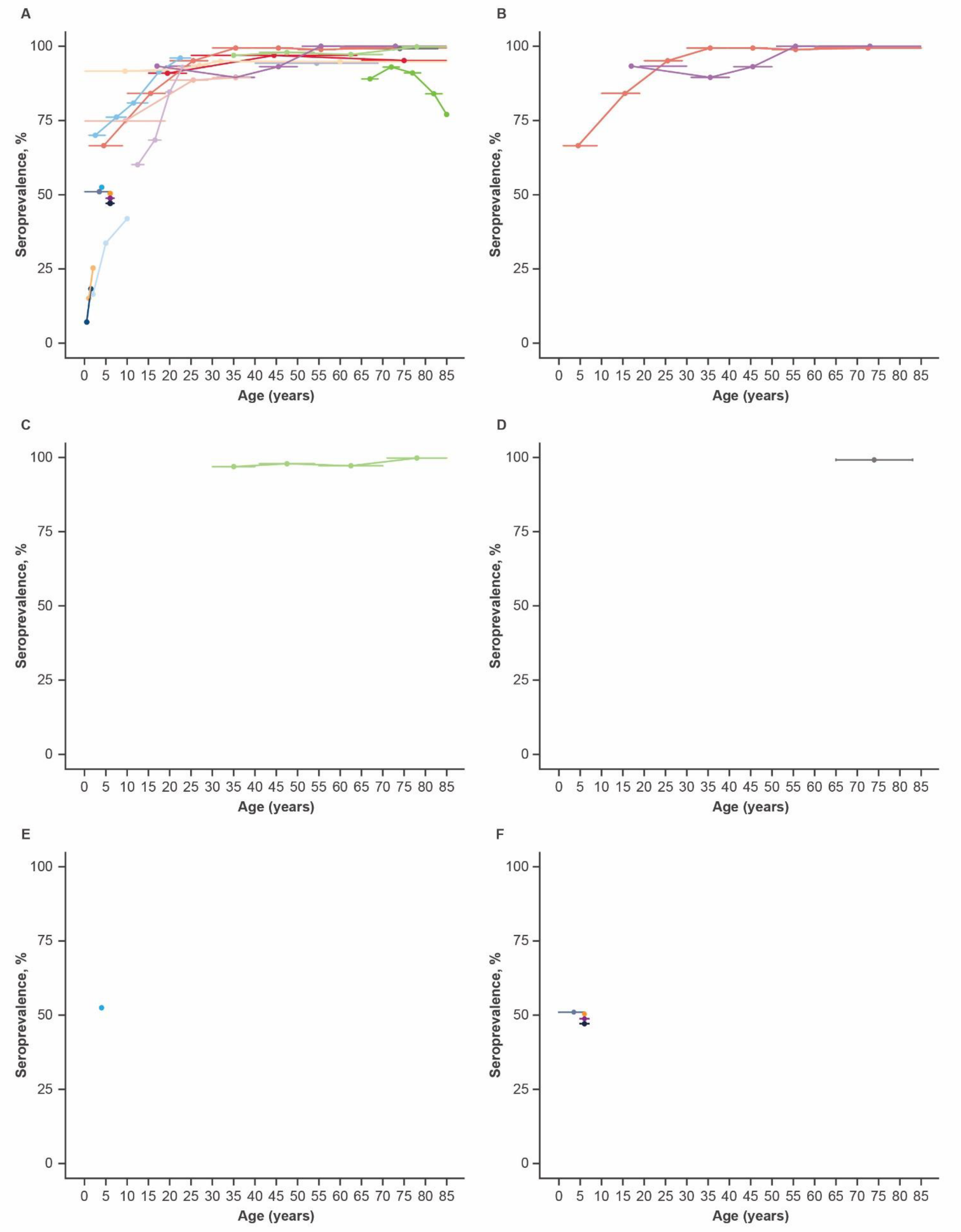

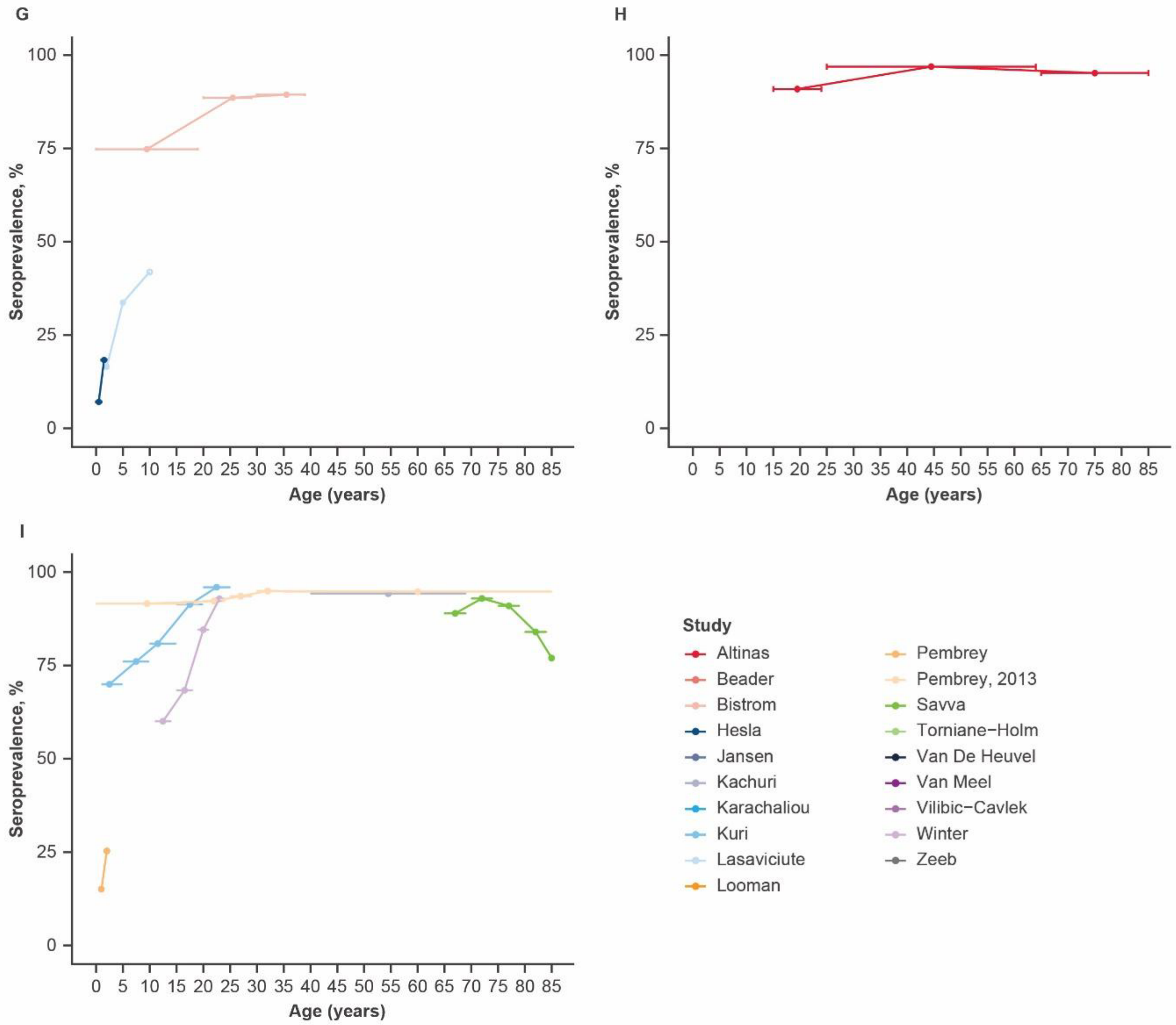

G
H
I
Seroprevalence, %
Age (years)
100
75
50
25
0
0 5 10 15 20 25 30 35 40 45 50 55 60 65 70 75 80 85
Study
Altinas
Beader
Bistrom
Hesla
Jansen
Kachuri
Karachaliou
Kuri
Lasaviciute
Looman
Pembrey
Pembrey, 2013
Savva
Torniane−Holm
Van De Heuvel
Van Meel
Vilibic−Cavlek
Winter
Zeeb

**Supplementary Figure 5. Study-specific seroprevalence (proportion) plotted for included studies[8,9,26,32,35,36,43,45,49,61,64,65,68,71]**. Meta-analysed across the following age groups: A) 0–4 years, B) 5–9 years, C) 10–14 years, D) 20–29 years, E) 30–39 years, and F) ≥40 years. Multiple data points were used from the same study if the study had a more than one seroprevalence measurement within the meta-analysis age-range. CI=confidence interval. ES=effect size.

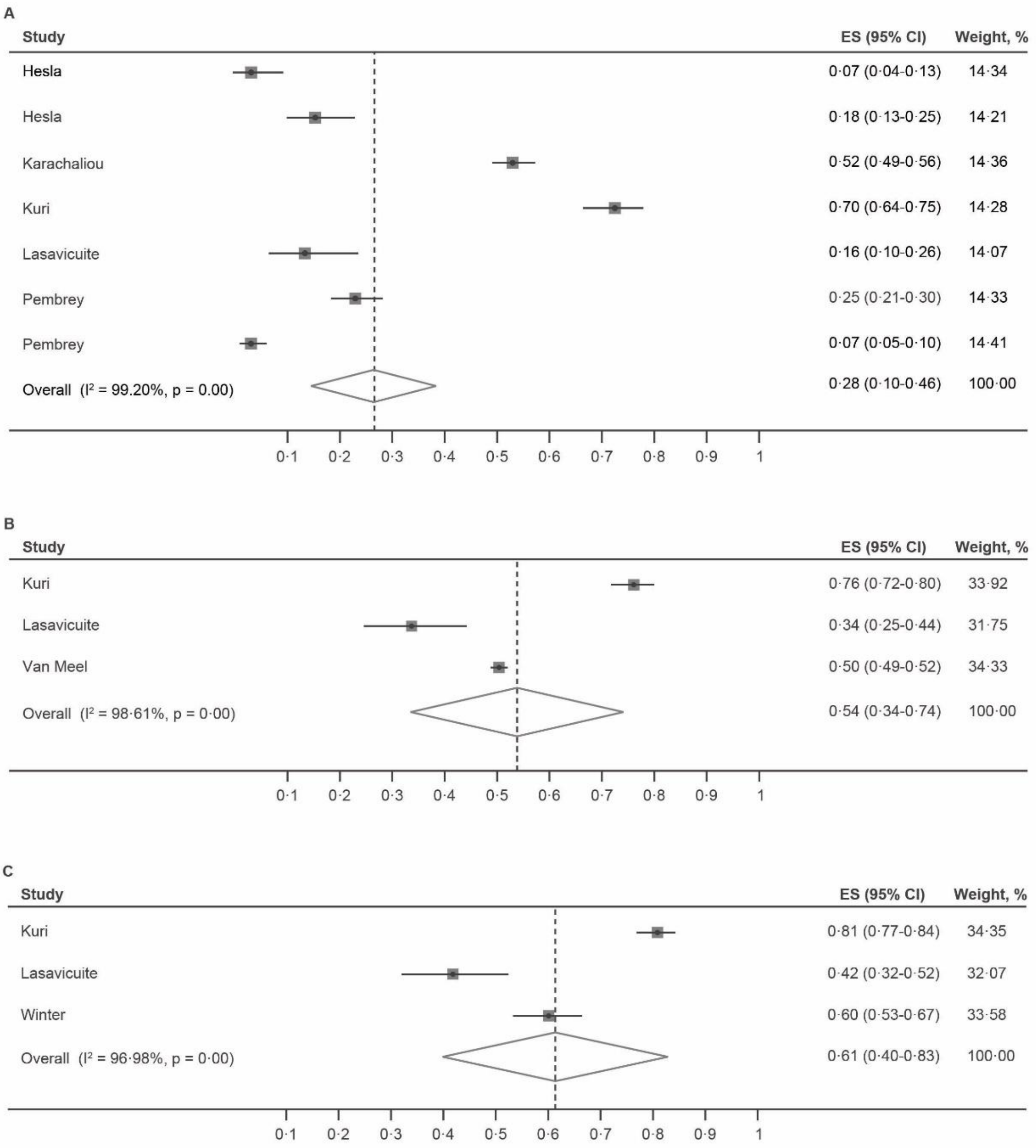

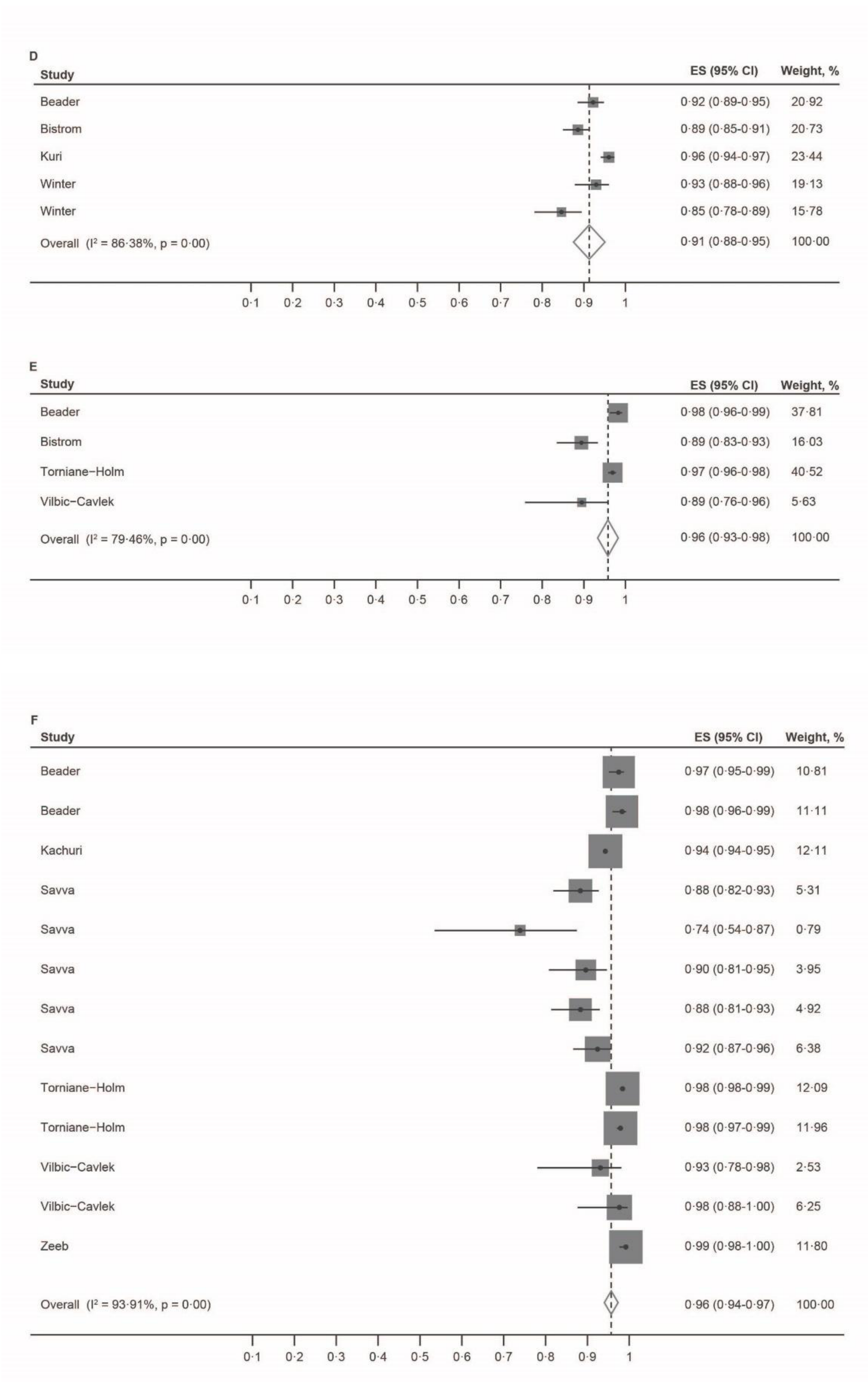
D
Study
ES (95% CI)
Weight, %
Beader 0·92 (0·89-0·95) 20·92
Bistrom 0·89 (0·85-0·91) 20·73
Kuri 0·96 (0·94-0·97) 23·44
Winter 0·93 (0·88-0·96) 19·13
Winter 0·85 (0·78-0·89) 15·78
Overall (I² = 86·38%, p = 0·00) 0·91 (0·88-0·95) 100·00
0·1 0·2 0·3 0·4 0·5 0·6 0·7 0·8 0·9 1
E
Study
ES (95% CI)
Weight, %
Beader 0·98 (0·96-0·99) 37·81
Bistrom 0·89 (0·83-0·93) 16·03
Torniane−Holm 0·97 (0·96-0·98) 40·52
Vilbic−Cavlek 0·89 (0·76-0·96) 5·63
Overall (I² = 79·46%, p = 0·00) 0·96 (0·93-0·98) 100·00
0·1 0·2 0·3 0·4 0·5 0·6 0·7 0·8 0·9 1
F
Study
ES (95% CI)
Weight, %
Beader 0·97 (0·95-0·99) 10·81
Beader 0·98 (0·96-0·99) 11·11
Kachuri 0·94 (0·94-0·95) 12·11
Savva 0·88 (0·82-0·93) 5·31
Savva 0·74 (0·54-0·87) 0·79
Savva 0·90 (0·81-0·95) 3·95
Savva 0·88 (0·81-0·93) 4·92
Savva 0·92 (0·87-0·96) 6·38
Torniane−Holm 0·98 (0·98-0·99) 12·09
Torniane−Holm 0·98 (0·97-0·99) 11·96
Vilbic−Cavlek 0·93 (0·78-0·98) 2·53
Vilbic−Cavlek 0·98 (0·88-1·00) 6·25
Zeeb 0·99 (0·98-1·00) 11·80
Overall (I² = 93·91%, p = 0·00) 0·96 (0·94-0·97) 100·00
0·1 0·2 0·3 0·4 0·5 0·6 0·7 0·8 0·9 1

**Supplementary Figure 6. Study-specific seroprevalence (proportion) plotted for included studies[8,9,26,32,35,36,43,45,49,61,64,65,68,71]**. Meta-analysed. A) 0–4 years, B) 20–29 years, C) ≥40 years. CI=confidence interval. ES=effect size.

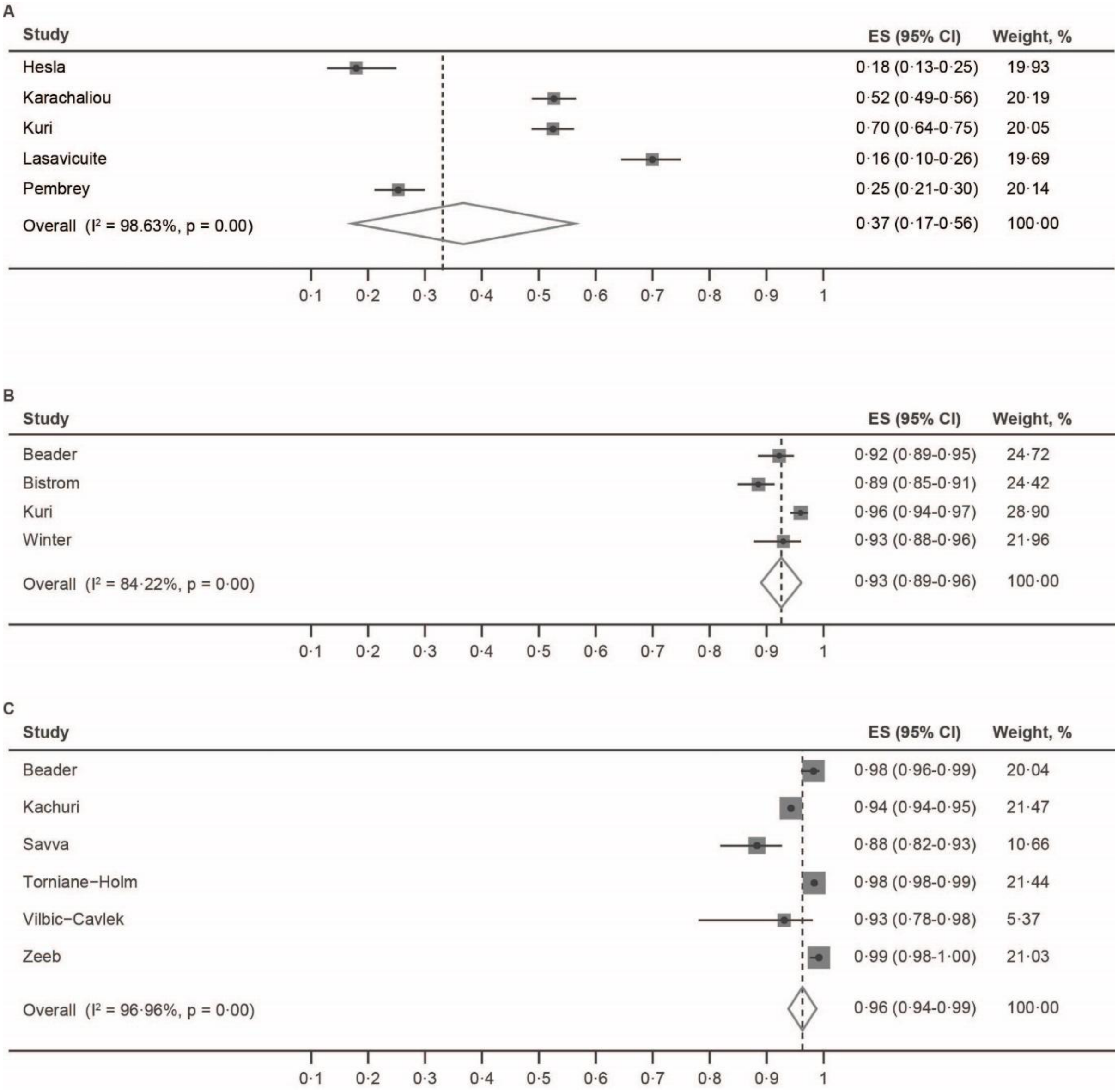

**Supplementary Figure 7. Seroprevalence by age (years).** A) Region of the Americas[3,5,6,14,15,18,19,62,67], B) Brazil, and C) USA. Horizontal lines represent the age groups covered by a given data point.

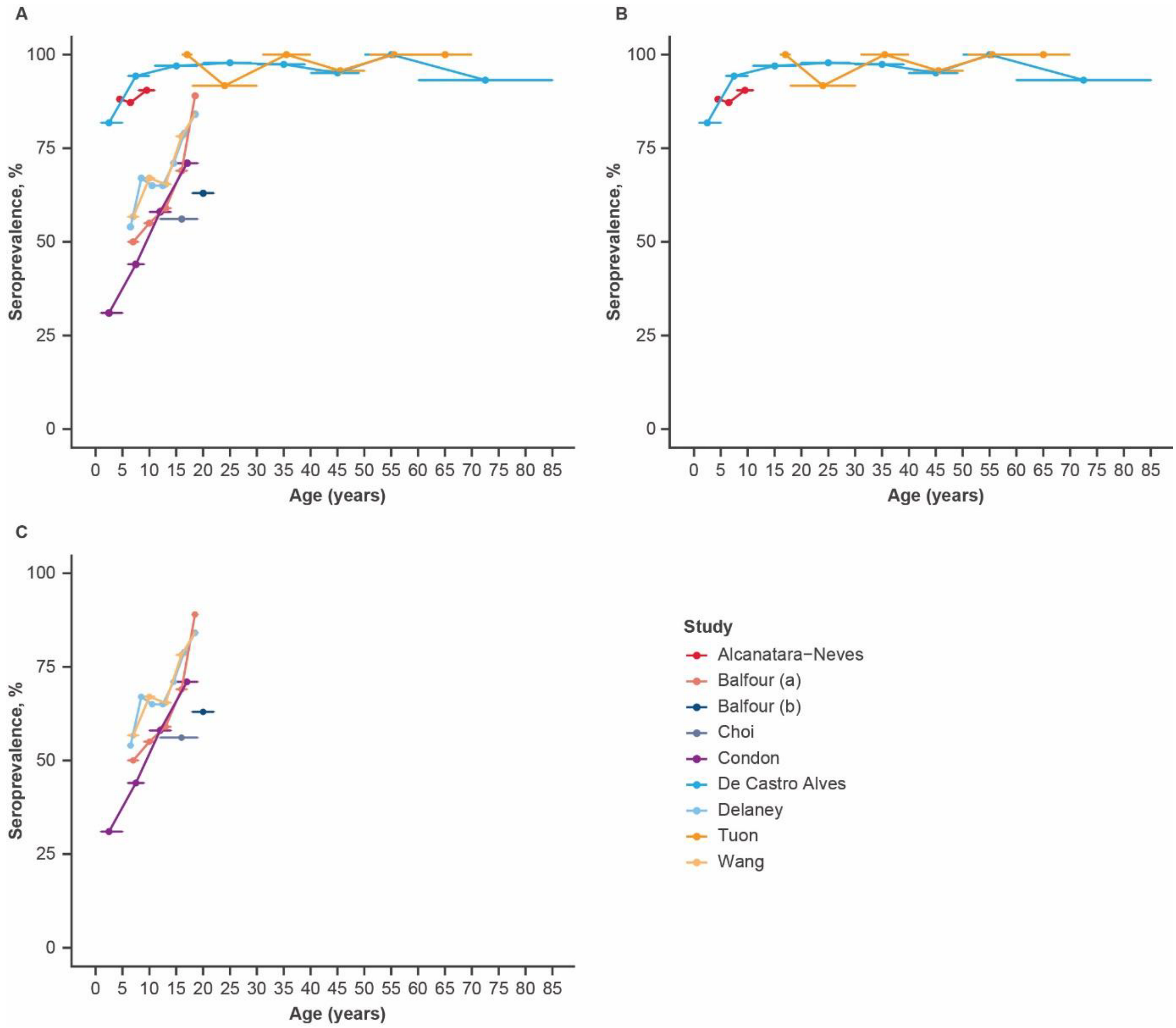

**Supplementary Figure 8. Study-specific seroprevalence (proportion) plotted for included studies[6,15,18,19] .** Meta-analysed across the following age categories: A) 5–9 years, and B) 15–19 years for the Region of the Americas. Multiple data points were used from the same study if the study had a more than one seroprevalence measurement within the meta-analysis age-range. CI=confidence interval. ES=effect size.

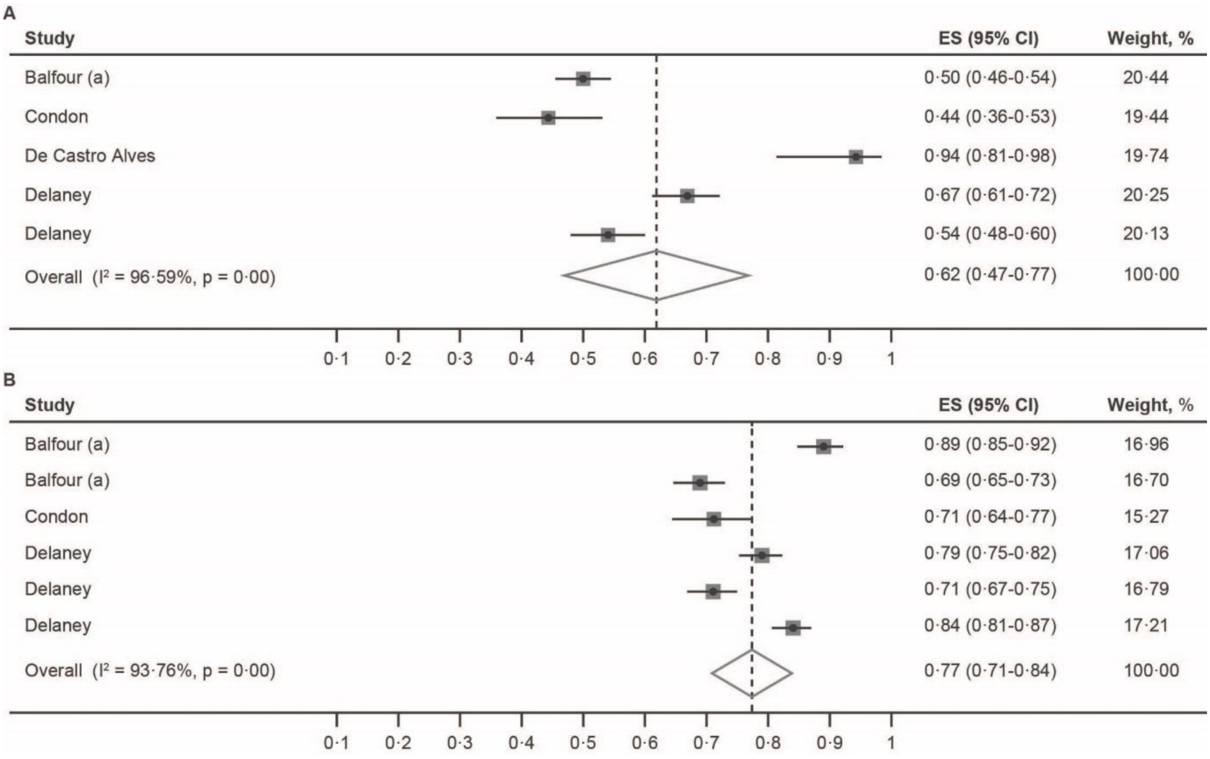

**Supplementary Figure 9. Study-specific seroprevalence plotted for included studies[6,15,18,19] .** Meta-analysed. A) 5–9 years, B) 15–19 years. CI=confidence interval. ES=effect size.

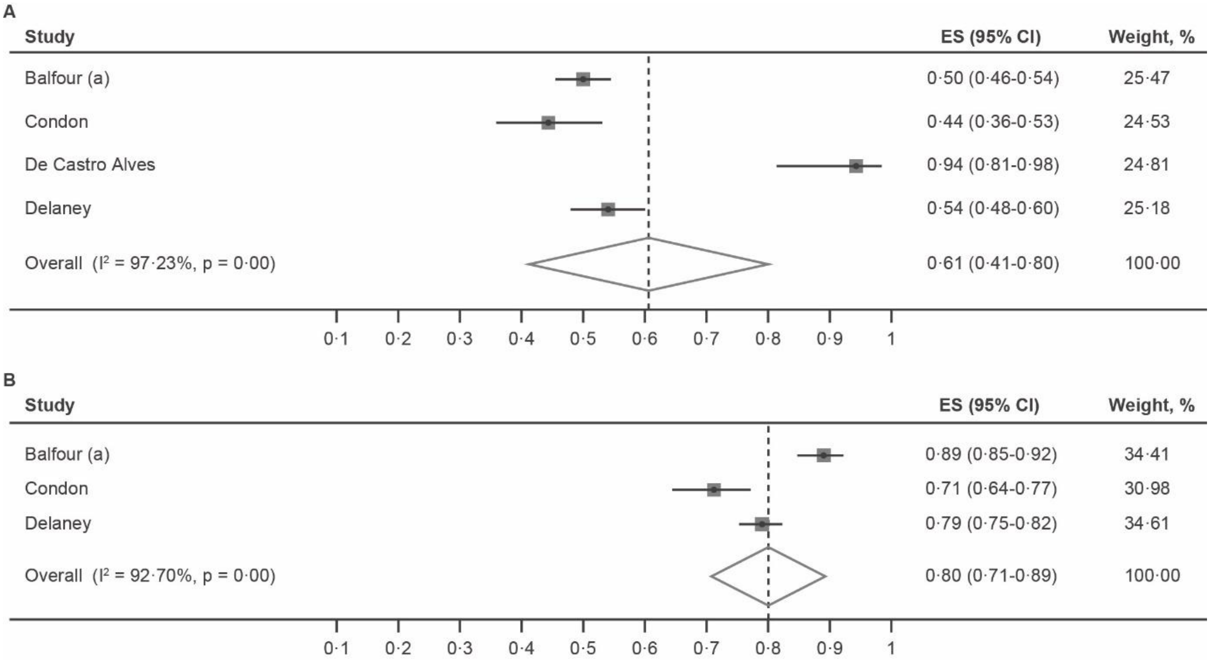

**Supplementary Figure 10. Seroprevalence by age (years).** South-East Asia Region (single study from Thailand). The study used was Suntornlohanakul.[58] Horizontal lines represent the age groups covered by a given data point.

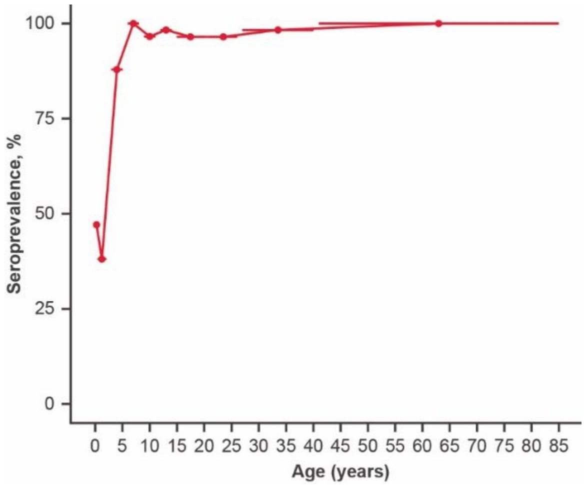

**Supplementary Figure 11. Seroprevalence by age (years)**. Eastern Mediterranean Region (single study from Iran). Study used was Sharifpour.[51] Horizontal lines represent the age groups covered by a given data point.

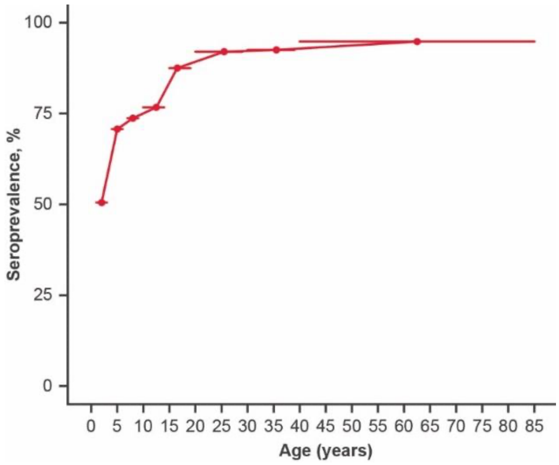

**Supplementary Figure 12. Seroprevalence by age (years).** A) Western Pacific Region[13,16,50,53,69,74] , B) China, and C) Singapore. Horizontal lines represent the age groups covered by a given data point.

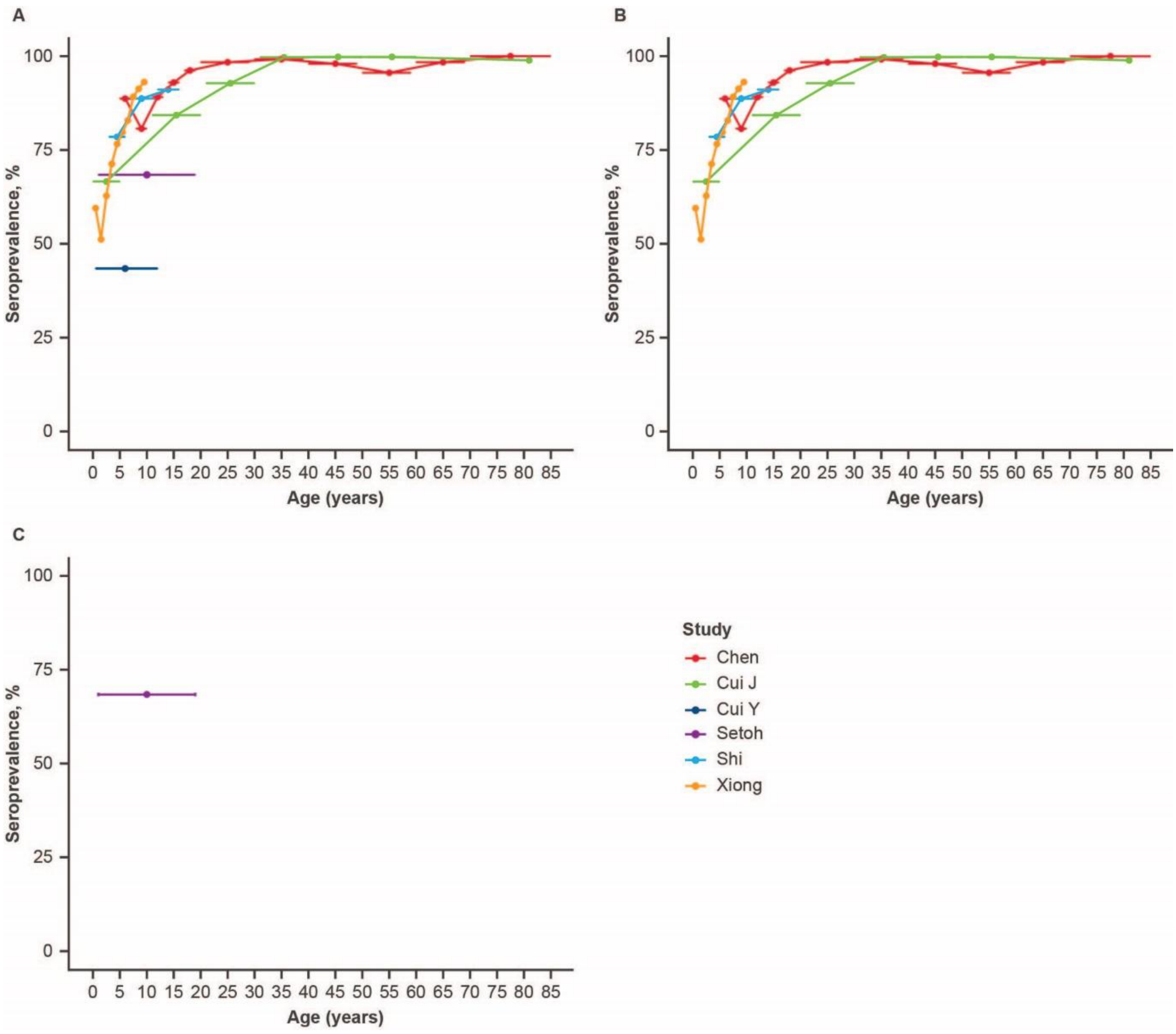

**Supplementary Table 7. Meta-regression results for region (EUR at baseline vs AMR)**

Meta-regression was possible for all age groups (0-4, 5-9, 10-14, 15-19, 20-29, 30-39, >40). The table shows the $R^2$ values for the meta-regression for each age category, which represent the percentage of between study heterogeneity in seroprevalence for these age groups.

| Age Group (years) | $R^2$ (%) |
|---|---|
| **0-4** | 3.1 |
| **5-9** | 25.1 |
| **10-14** | 0.0 |
| **15-19** | 0.0 |
| **20-29** | 25.3 |
| **30-39** | 0.0 |
| **>40** | 28.5 |

**Supplementary Figure 13. Seroprevalence by age (years).** A) high income, B) middle income, and C) low income countries[2-9,13–16,18, 19,24,26,27,30,35,36,38,41,43,45,49,50,51,53,55,58,61-66,68,69,71]. Horizontal lines represent the age groups covered by a given data point.

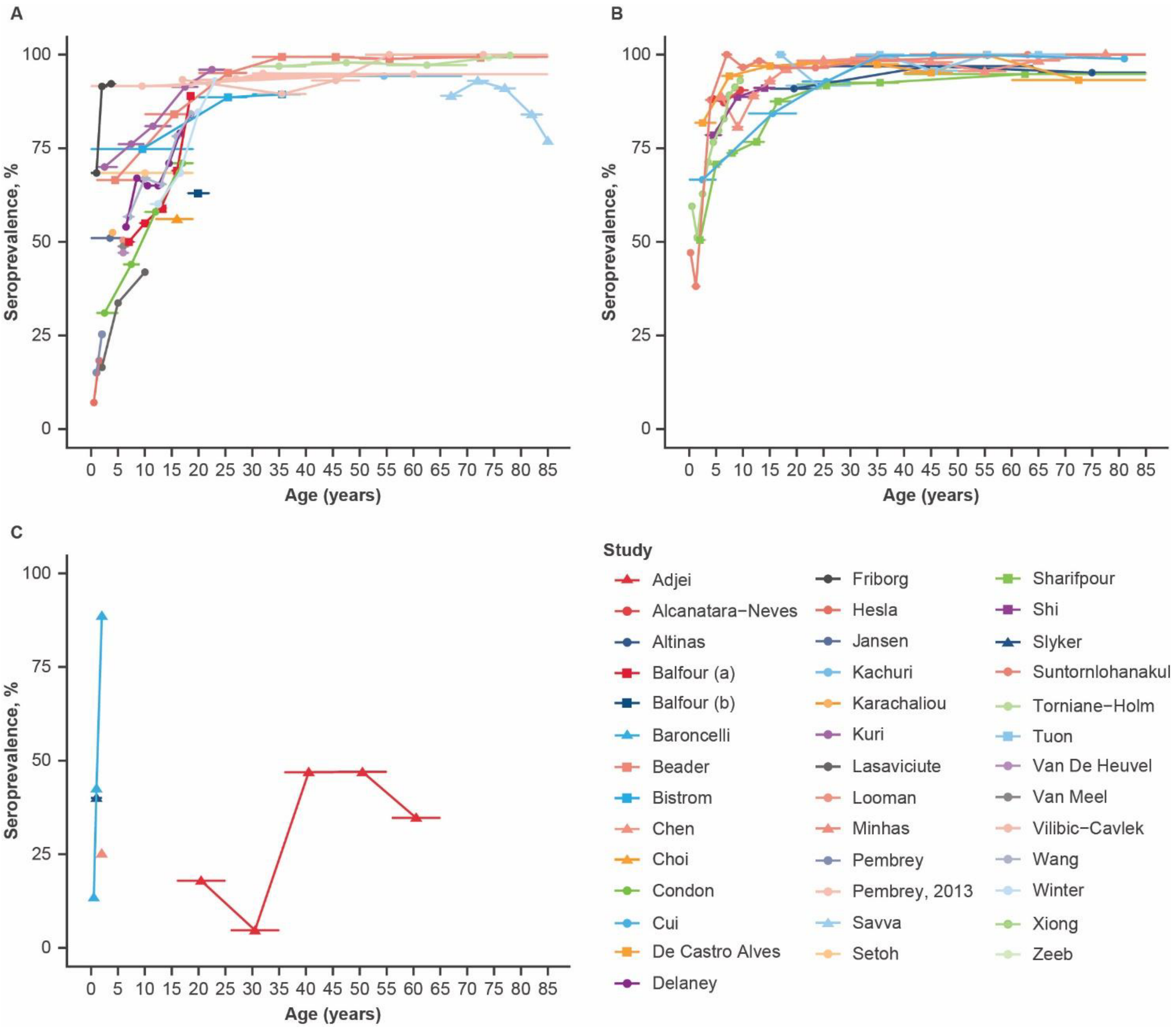

**Supplementary Figure 14. Study-specific seroprevalence (proportion) plotted for included[6,8,9,15,19,26,35,36,43,49,61,64,65,68,71,77,78] studies.** Meta-analysed across the following age groups: A) 0–4 years, B) 5–9 years, C) 10–14 years, D) 15–19 years, E) 20–29 years, F) 30–39 years, and G) ≥40 years. Multiple data points were used from the same study if the study had a more than one seroprevalence measurement within the meta-analysis age-range. CI=confidence interval. ES=effect size.

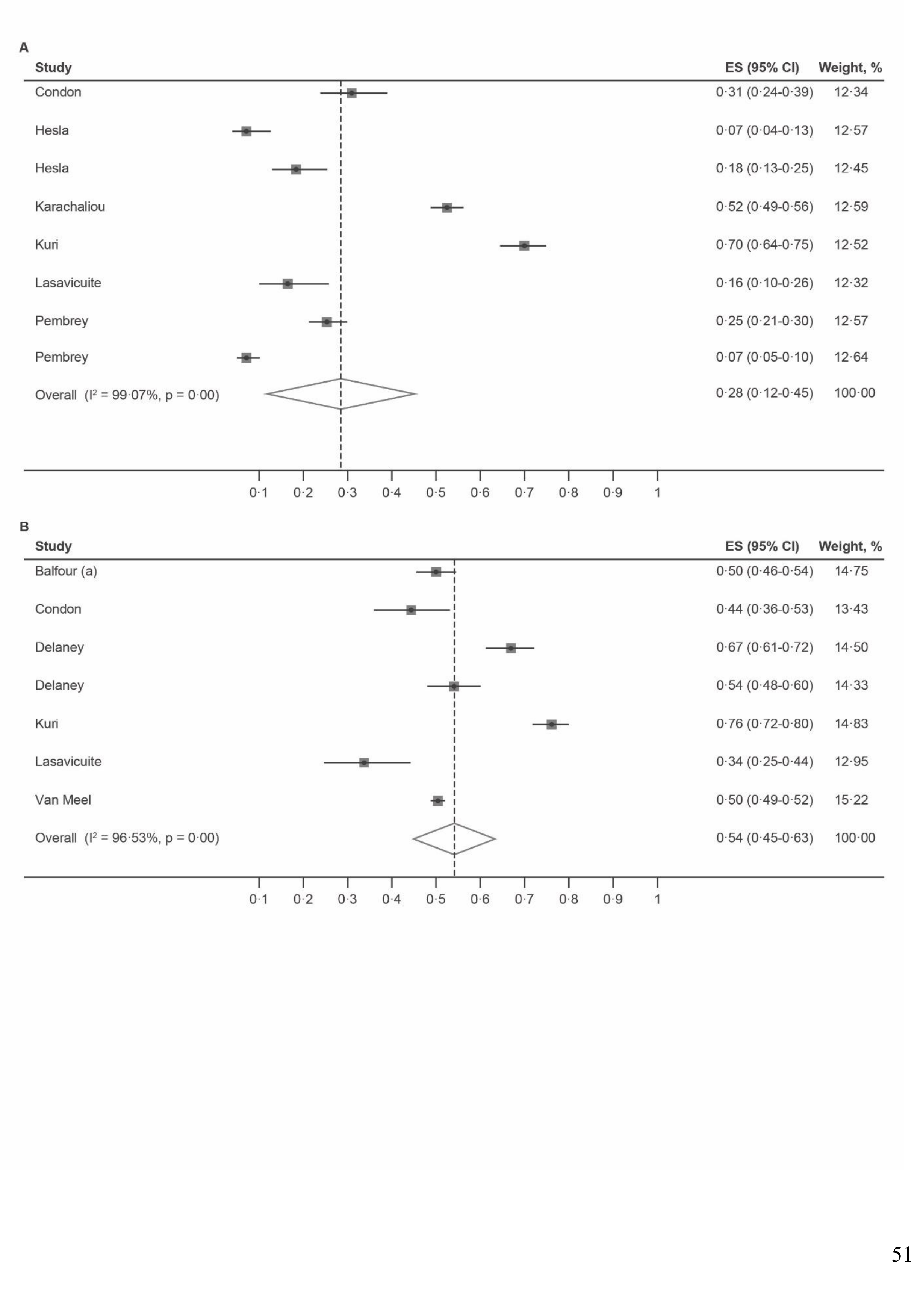

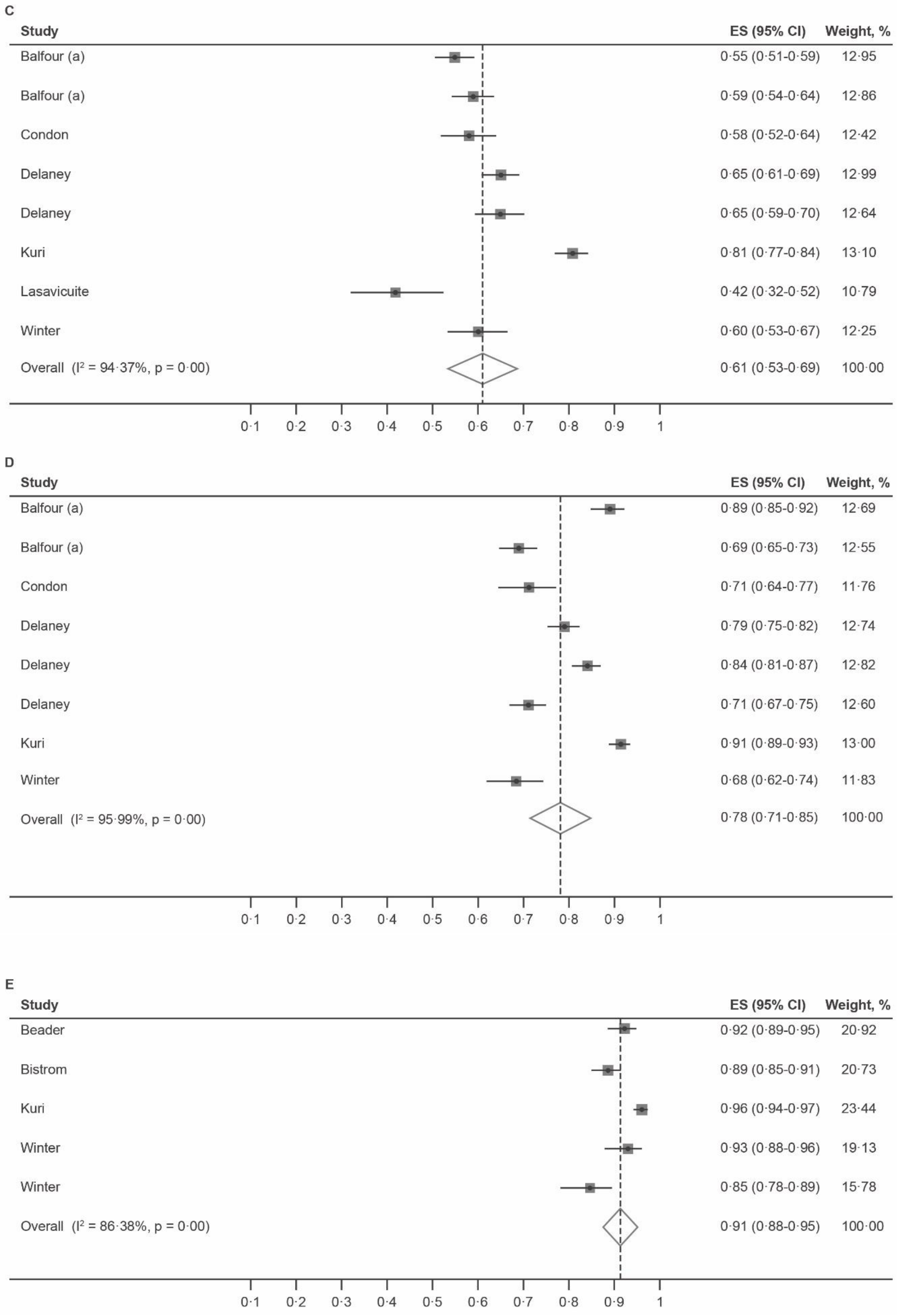
C
Study
ES (95% CI)
Weight, %
Balfour (a)
0·55 (0·51-0·59)
12·95
Balfour (a)
0·59 (0·54-0·64)
12·86
Condon
0·58 (0·52-0·64)
12·42
Delaney
0·65 (0·61-0·69)
12·99
Delaney
0·65 (0·59-0·70)
12·64
Kuri
0·81 (0·77-0·84)
13·10
Lasavicuite
0·42 (0·32-0·52)
10·79
Winter
0·60 (0·53-0·67)
12·25
Overall (I² = 94·37%, p = 0·00)
0·61 (0·53-0·69)
100·00
0·1
0·2
0·3
0·4
0·5
0·6
0·7
0·8
0·9
1
D
Study
ES (95% CI)
Weight, %
Balfour (a)
0·89 (0·85-0·92)
12·69
Balfour (a)
0·69 (0·65-0·73)
12·55
Condon
0·71 (0·64-0·77)
11·76
Delaney
0·79 (0·75-0·82)
12·74
Delaney
0·84 (0·81-0·87)
12·82
Delaney
0·71 (0·67-0·75)
12·60
Kuri
0·91 (0·89-0·93)
13·00
Winter
0·68 (0·62-0·74)
11·83
Overall (I² = 95·99%, p = 0·00)
0·78 (0·71-0·85)
100·00
0·1
0·2
0·3
0·4
0·5
0·6
0·7
0·8
0·9
1
E
Study
ES (95% CI)
Weight, %
Beader
0·92 (0·89-0·95)
20·92
Bistrom
0·89 (0·85-0·91)
20·73
Kuri
0·96 (0·94-0·97)
23·44
Winter
0·93 (0·88-0·96)
19·13
Winter
0·85 (0·78-0·89)
15·78
Overall (I² = 86·38%, p = 0·00)
0·91 (0·88-0·95)
100·00
0·1
0·2
0·3
0·4
0·5
0·6
0·7
0·8
0·9
1

F

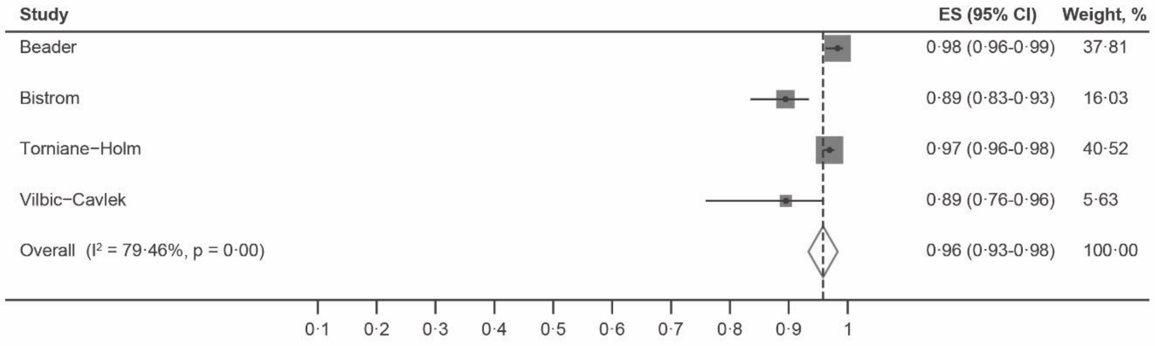
Study
ES (95% CI)
Weight, %
Beader
0·98 (0·96-0·99)
37·81
Bistrom
0·89 (0·83-0·93)
16·03
Torniane−Holm
0·97 (0·96-0·98)
40·52
Vilbic−Cavlek
0·89 (0·76-0·96)
5·63
Overall (I² = 79·46%, p = 0·00)
0·96 (0·93-0·98)
100·00
0·1
0·2
0·3
0·4
0·5
0·6
0·7
0·8
0·9
1

G

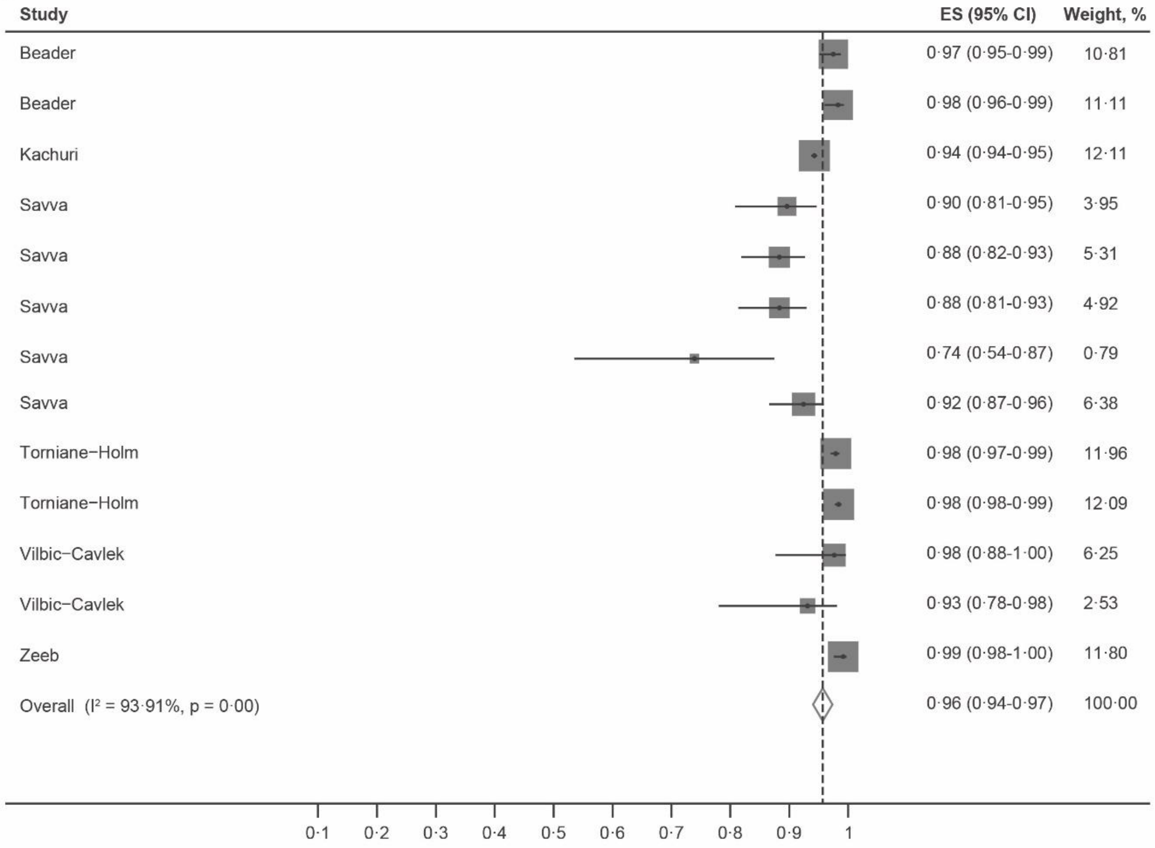
Study
ES (95% CI)
Weight, %
Beader
0·97 (0·95-0·99)
10·81
Beader
0·98 (0·96-0·99)
11·11
Kachuri
0·94 (0·94-0·95)
12·11
Savva
0·90 (0·81-0·95)
3·95
Savva
0·88 (0·82-0·93)
5·31
Savva
0·88 (0·81-0·93)
4·92
Savva
0·74 (0·54-0·87)
0·79
Savva
0·92 (0·87-0·96)
6·38
Torniane−Holm
0·98 (0·97-0·99)
11·96
Torniane−Holm
0·98 (0·98-0·99)
12·09
Vilbic−Cavlek
0·98 (0·88-1·00)
6·25
Vilbic−Cavlek
0·93 (0·78-0·98)
2·53
Zeeb
0·99 (0·98-1·00)
11·80
Overall (I² = 93·91%, p = 0·00)
0·96 (0·94-0·97)
100·00
0·1
0·2
0·3
0·4
0·5
0·6
0·7
0·8
0·9
1

**Supplementary Figure 15. Study-specific seroprevalence (proportion) plotted for included studies**[6,8,9,15,19,26,35,36,43,49,61,64,65,68,71,76–78]. Meta-analysed. A) 0–4 years, B) 5–9 years, C) 10–14 years, D) 15–19 years, E) 20–29 years, and F) ≥40 years. CI=confidence interval. ES=effect size.

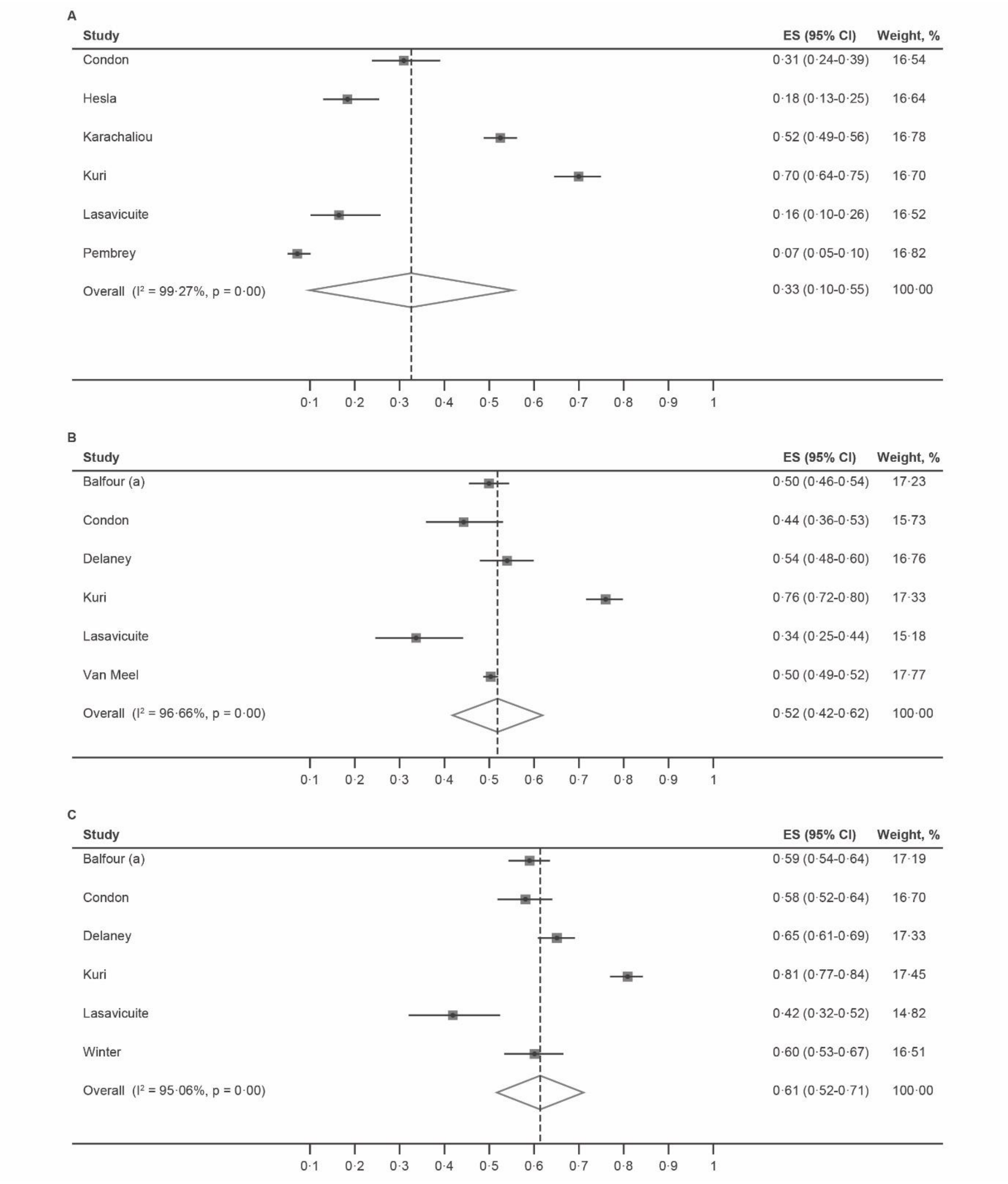

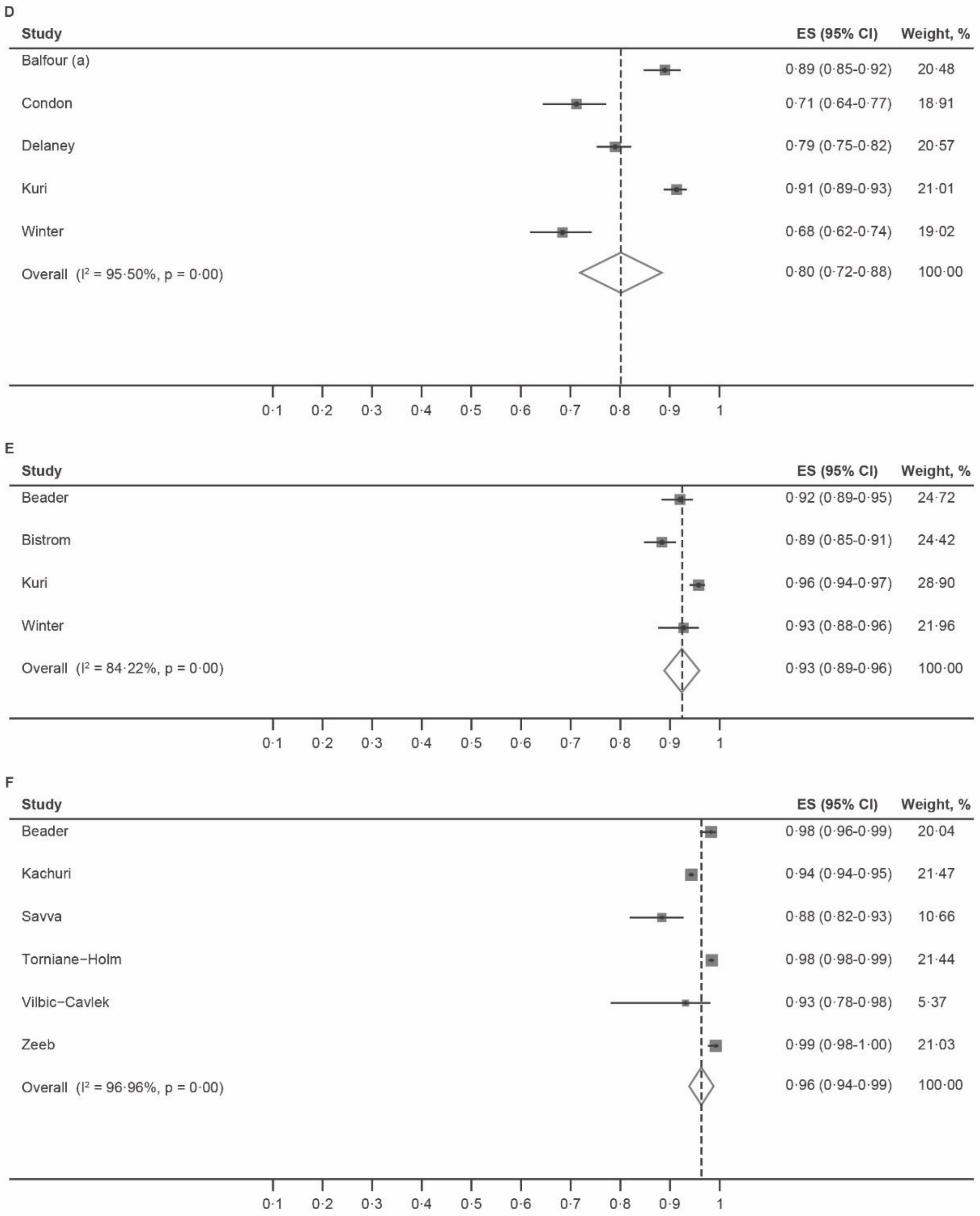
D
Study
ES (95% CI)
Weight, %
Balfour (a) 0·89 (0·85-0·92) 20·48
Condon 0·71 (0·64-0·77) 18·91
Delaney 0·79 (0·75-0·82) 20·57
Kuri 0·91 (0·89-0·93) 21·01
Winter 0·68 (0·62-0·74) 19·02
Overall (I² = 95·50%, p = 0·00) 0·80 (0·72-0·88) 100·00
0·1 0·2 0·3 0·4 0·5 0·6 0·7 0·8 0·9 1
E
Study
ES (95% CI)
Weight, %
Beader 0·92 (0·89-0·95) 24·72
Bistrom 0·89 (0·85-0·91) 24·42
Kuri 0·96 (0·94-0·97) 28·90
Winter 0·93 (0·88-0·96) 21·96
Overall (I² = 84·22%, p = 0·00) 0·93 (0·89-0·96) 100·00
0·1 0·2 0·3 0·4 0·5 0·6 0·7 0·8 0·9 1
F
Study
ES (95% CI)
Weight, %
Beader 0·98 (0·96-0·99) 20·04
Kachuri 0·94 (0·94-0·95) 21·47
Savva 0·88 (0·82-0·93) 10·66
Torniane−Holm 0·98 (0·98-0·99) 21·44
Vilbic−Cavlek 0·93 (0·78-0·98) 5·37
Zeeb 0·99 (0·98-1·00) 21·03
Overall (I² = 96·96%, p = 0·00) 0·96 (0·94-0·99) 100·00
0·1 0·2 0·3 0·4 0·5 0·6 0·7 0·8 0·9 1

**Supplementary Figure 16. Study-specific seroprevalence (proportion) plotted for included studies**[16,18,51,58,62] . Meta-analysed across the following age groups: A) 0–4 years, B) 5–9 years, C) 20–29 years, D) 30–39 years, and E) ≥40 years. CI=confidence interval. Multiple data points were used from the same study if the study had a more than one seroprevalence measurement within the meta-analysis age-range. ES=effect size.

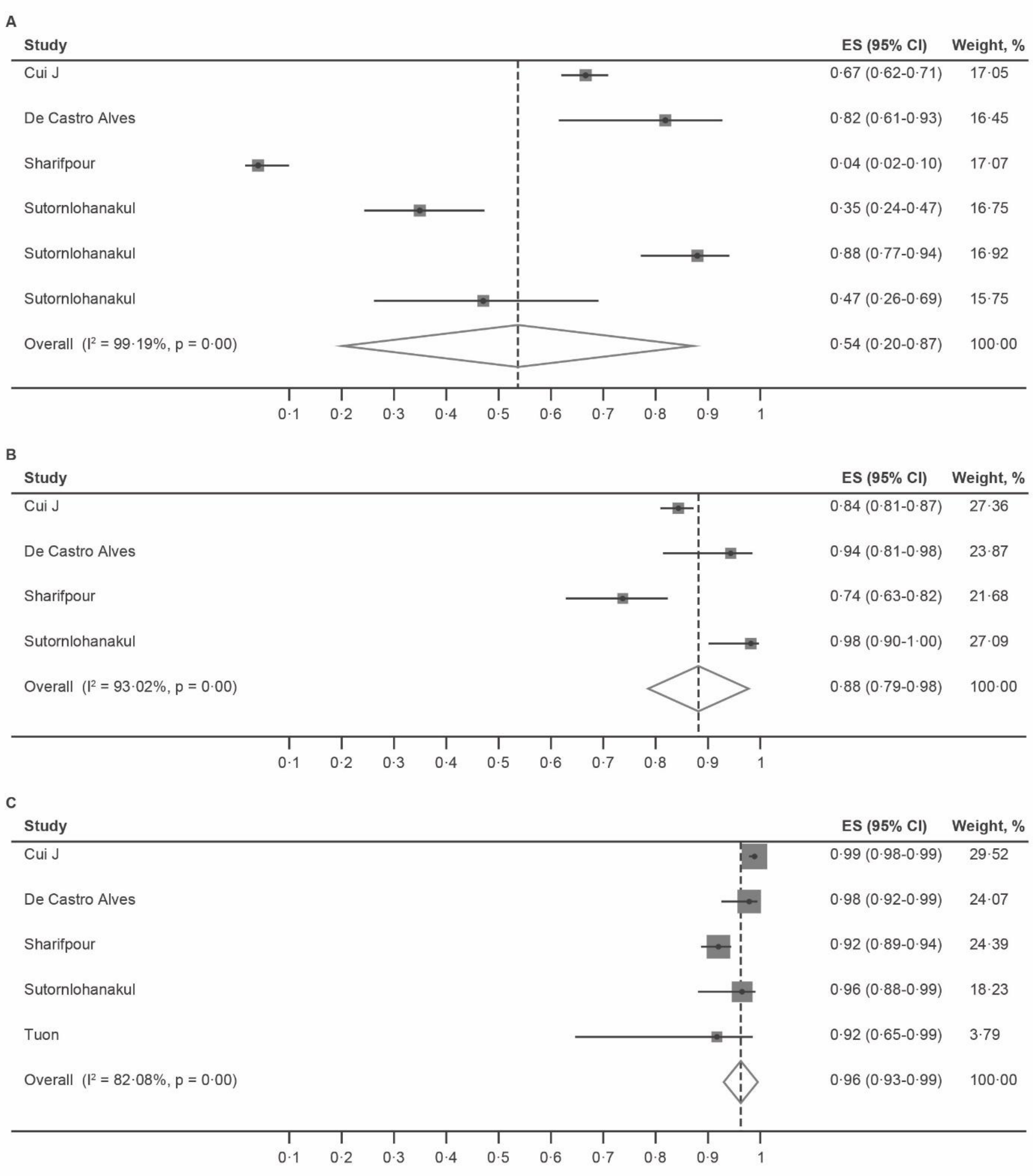

D

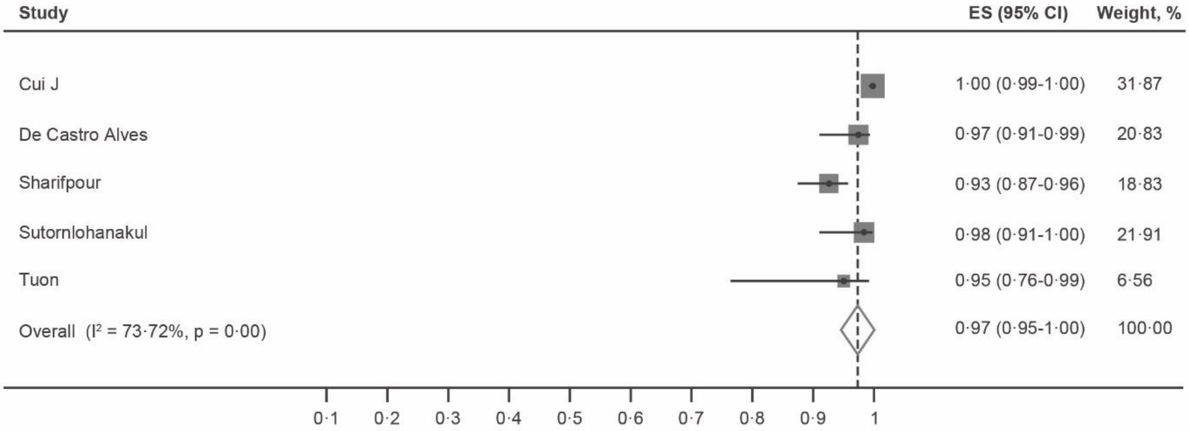
Study
ES (95% CI)
Weight, %
Cui J 1·00 (0·99-1·00) 31·87
De Castro Alves 0·97 (0·91-0·99) 20·83
Sharifpour 0·93 (0·87-0·96) 18·83
Sutornlohanakul 0·98 (0·91-1·00) 21·91
Tuon 0·95 (0·76-0·99) 6·56
Overall (I² = 73·72%, p = 0·00) 0·97 (0·95-1·00) 100·00
0·1 0·2 0·3 0·4 0·5 0·6 0·7 0·8 0·9 1

E

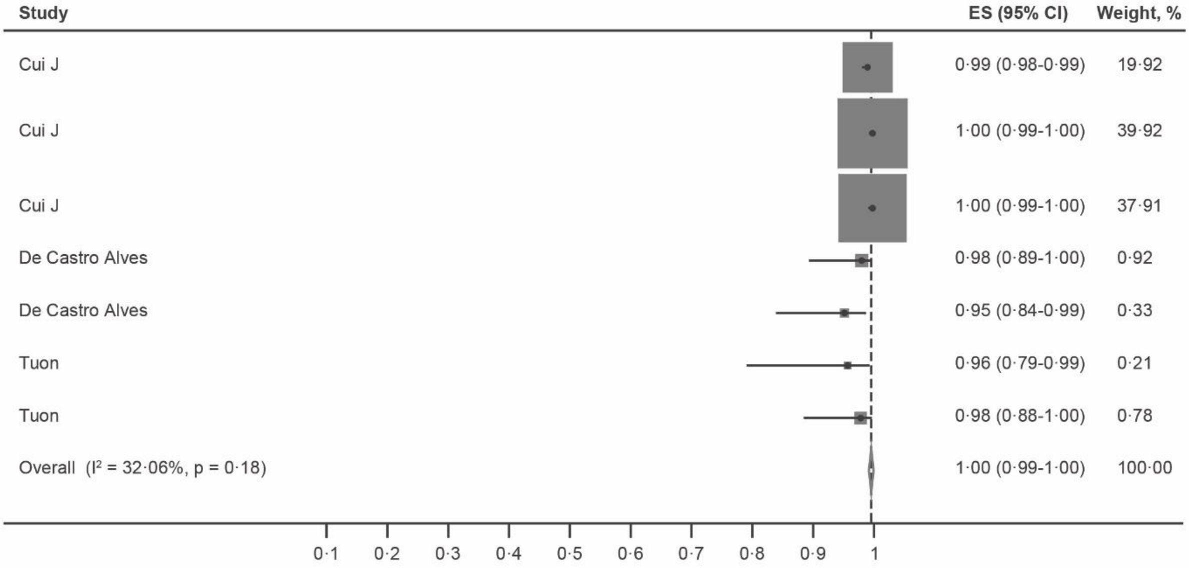
Study
ES (95% CI)
Weight, %
Cui J 0·99 (0·98-0·99) 19·92
Cui J 1·00 (0·99-1·00) 39·92
Cui J 1·00 (0·99-1·00) 37·91
De Castro Alves 0·98 (0·89-1·00) 0·92
De Castro Alves 0·95 (0·84-0·99) 0·33
Tuon 0·96 (0·79-0·99) 0·21
Tuon 0·98 (0·88-1·00) 0·78
Overall (I² = 32·06%, p = 0·18) 1·00 (0·99-1·00) 100·00
0·1 0·2 0·3 0·4 0·5 0·6 0·7 0·8 0·9 1

**Supplementary Figure 17. Study-specific seroprevalence (proportion) plotted for included studies[16,18,51,58,62]**. Meta-analysed. A) 0–4 years, B) 5–9 years, and C) ≥40 years. CI=confidence interval. ES=effect size.

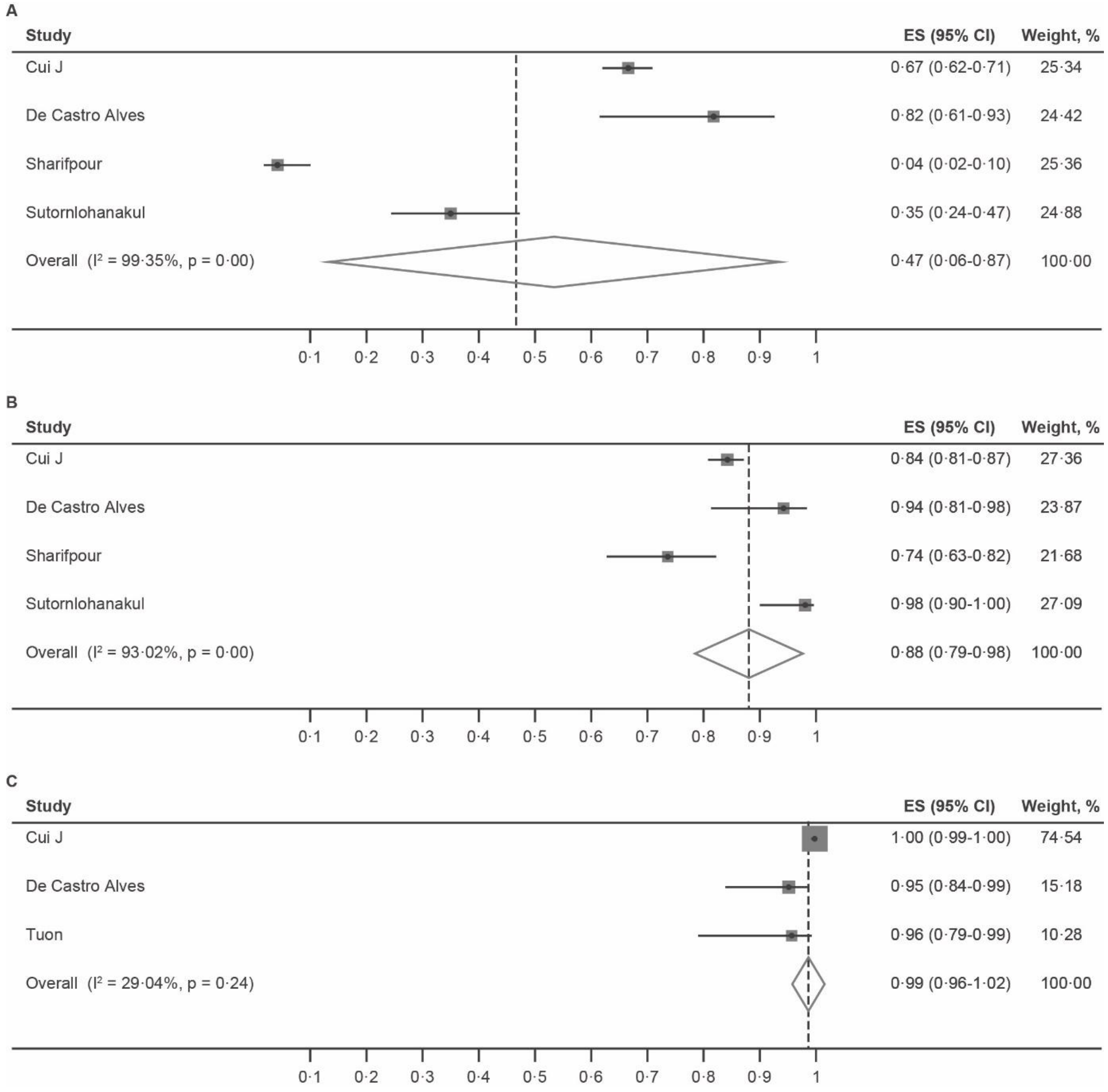

**Supplementary Table 8. Meta-regression results for income-level (High-income at baseline vs middle-income)**

Meta-regression was possible for all but the 15–19-year-old age group (0-4, 5-9, 10-14, 15-19, 20-29, 30-39, >40). The table shows the $R^2$ values for the meta-regression for each age category, which represent the percentage of between study heterogeneity in seroprevalence for these age groups.

| **Age Group (years)** | **$R^2$ (%)** |
|---|---|
| **0-4** | 9.5 |
| **5-9** | 63.4 |
| **10-14** | 7.6 |
| **20-29** | 25.6 |
| **30-39** | 0.0 |
| **>40** | 28.5 |

**Supplementary Table 9. Number of studies per country**

| Country | Number of Studies |
|---|---|
| Ghana | 1 |
| Kenya | 2 |
| Malawi | 1 |
| Zambia | 1 |
| UK | 6 |
| Sweden | 3 |
| Netherlands | 3 |
| Croatia | 2 |
| Germany | 1 |
| Finland | 1 |
| Turkey | 1 |
| USA | 6 |
| Brazil | 3 |
| Thailand | 1 |
| Iran | 1 |
| China/Taiwan | 4 |
| Singapore | 1 |

**Supplementary Figure 18. Study-specific seroprevalence (proportion) plotted for included studies[6,15,19]**
Meta-analysed across the following age categories: A) 5–9 years, B) 10–14 years, and C) 15–19 years. Multiple data points were used from the same study if the study had a more than one seroprevalence measurement within the meta-analysis age-range. CI=confidence interval. ES=effect size.

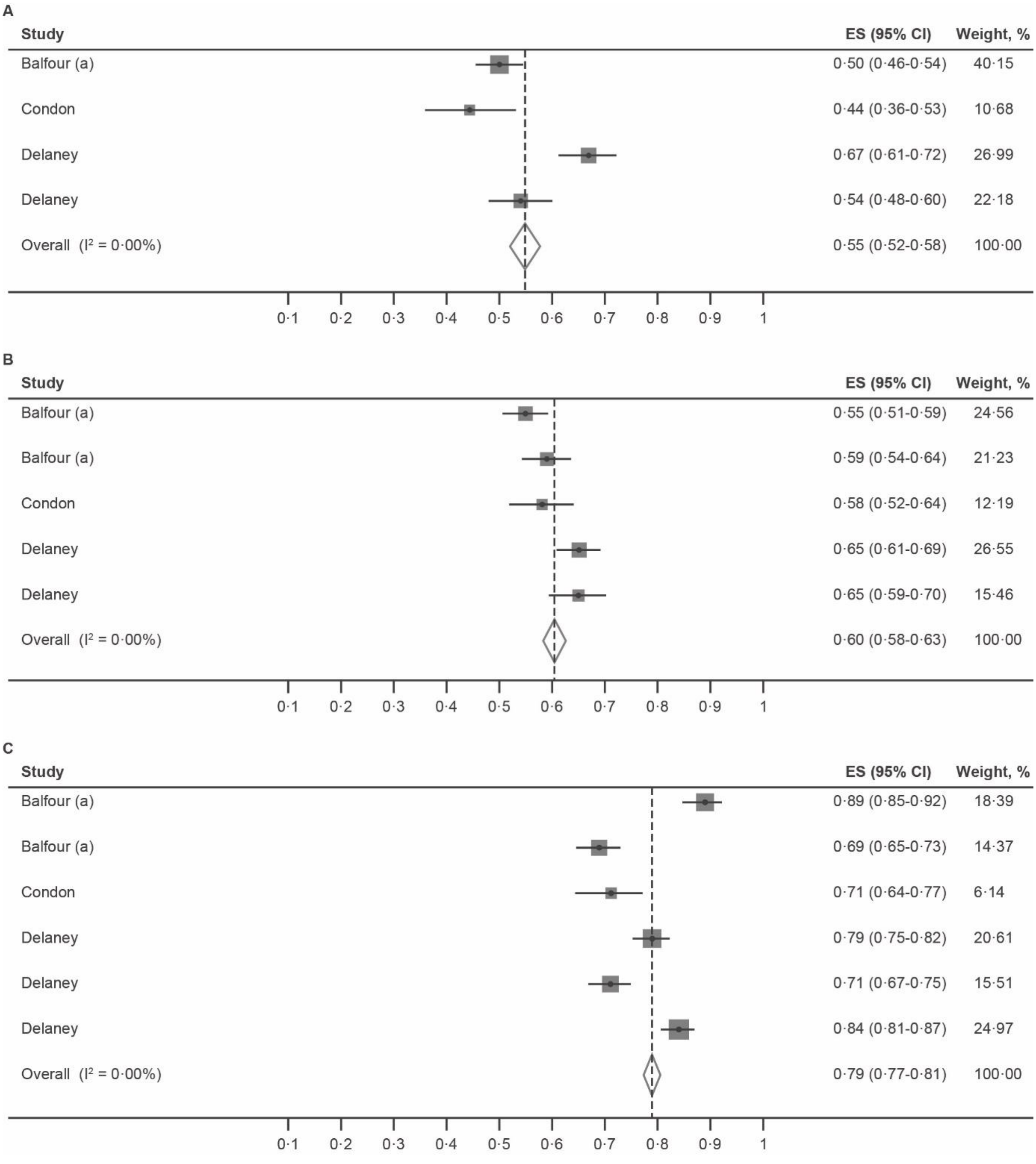

**Supplementary Figure 19. Study-specific seroprevalence (proportion) plotted for included studies[6,15,19].** Meta-analysed A) 5–9 years, B) 15–19 years. CI=confidence interval. ES=effect size.

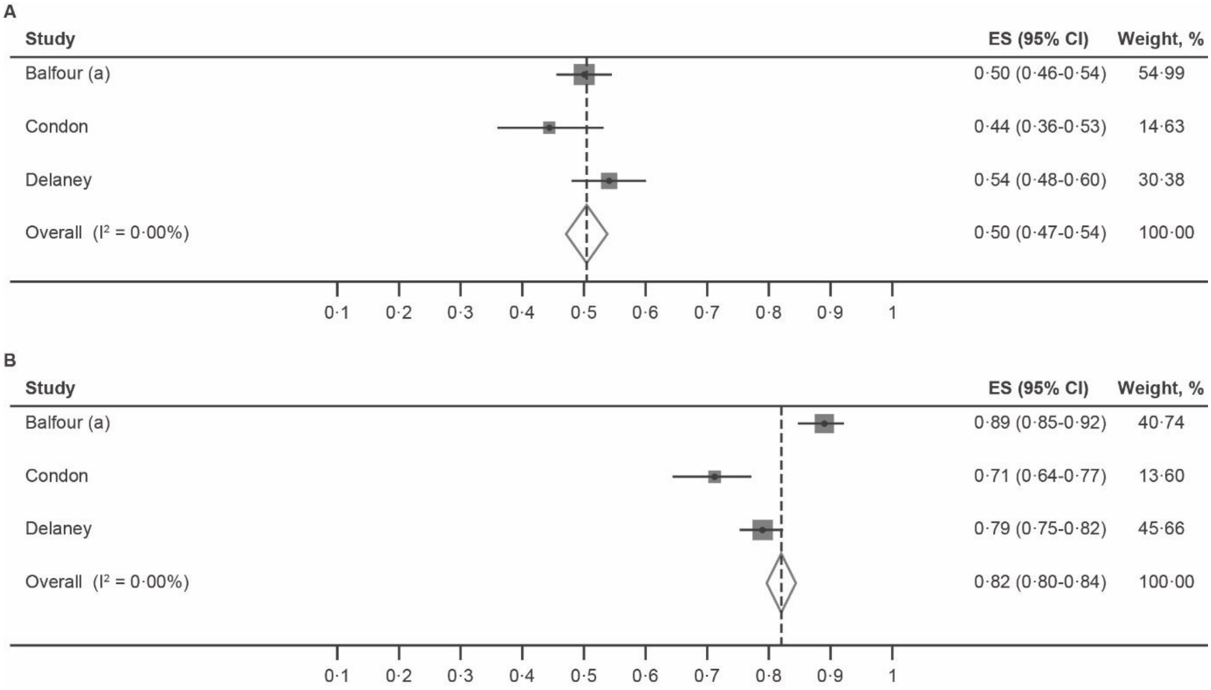

**Supplementary Figure 20. Cumulative study-specific seroprevalence (proportion) plotted by year of recruitment for included studies [6,15,19].** A) 5–9 years, B) 10–14 years, C) 15-19 years. E CI=confidence interval. ES=effect size. For studies with recruitment periods spanning several years, the midpoint was used as the reference year.

**A)**

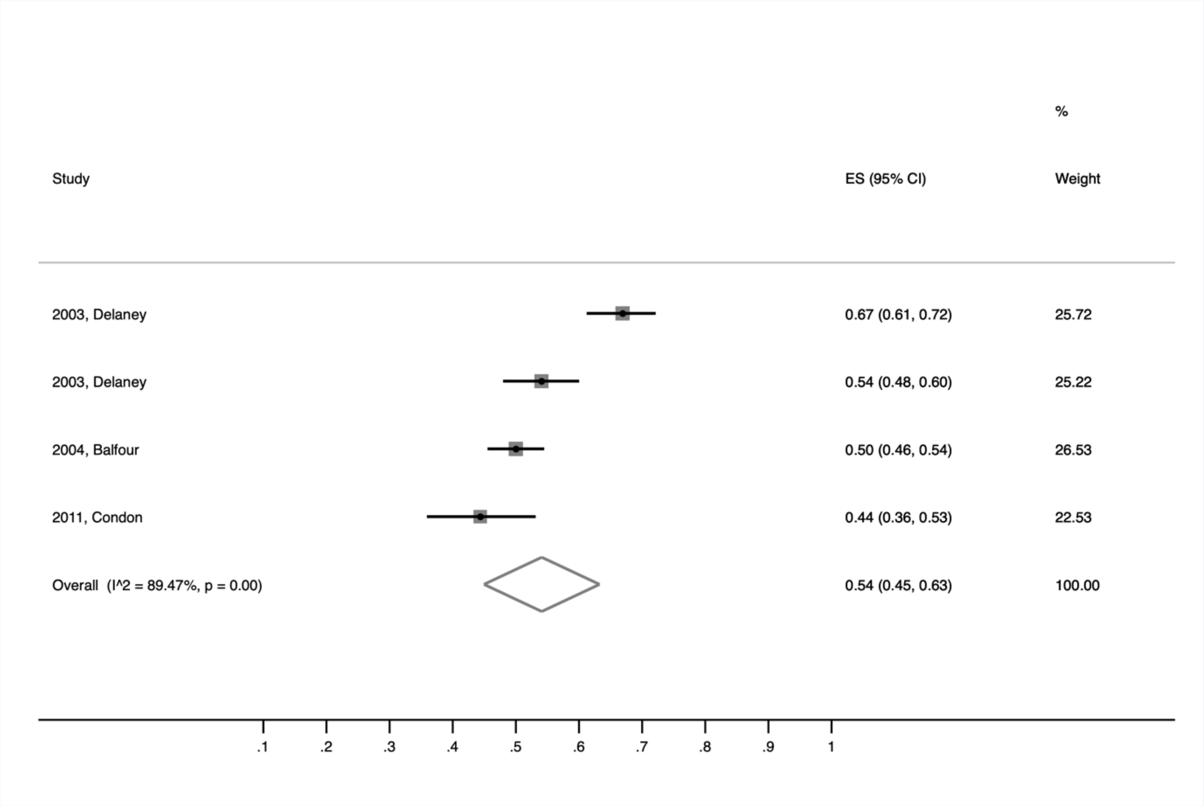

**B)**

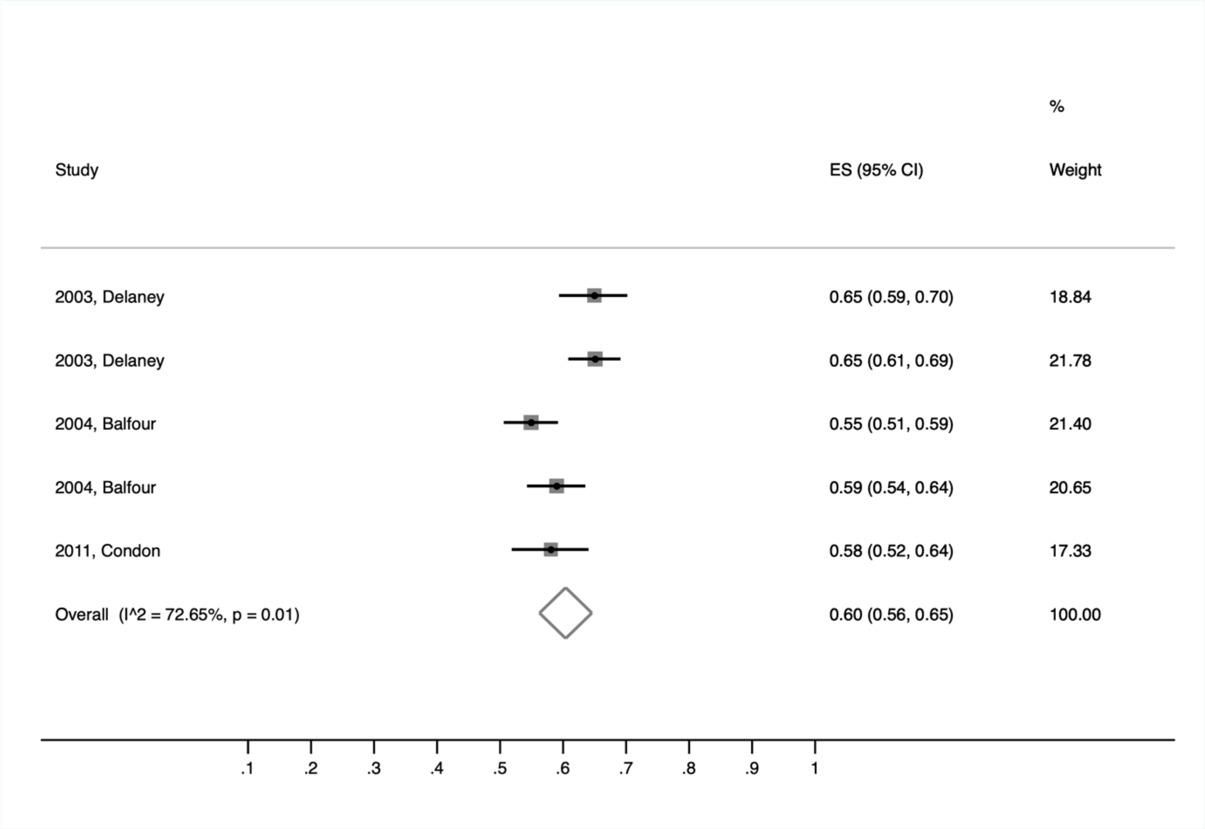
Study
ES (95% CI)
%
Weight
2003, Delaney
0.65 (0.59, 0.70)
18.84
2003, Delaney
0.65 (0.61, 0.69)
21.78
2004, Balfour
0.55 (0.51, 0.59)
21.40
2004, Balfour
0.59 (0.54, 0.64)
20.65
2011, Condon
0.58 (0.52, 0.64)
17.33
Overall (I^2 = 72.65%, p = 0.01)
0.60 (0.56, 0.65)
100.00
.1
.2
.3
.4
.5
.6
.7
.8
.9
1

**C)**

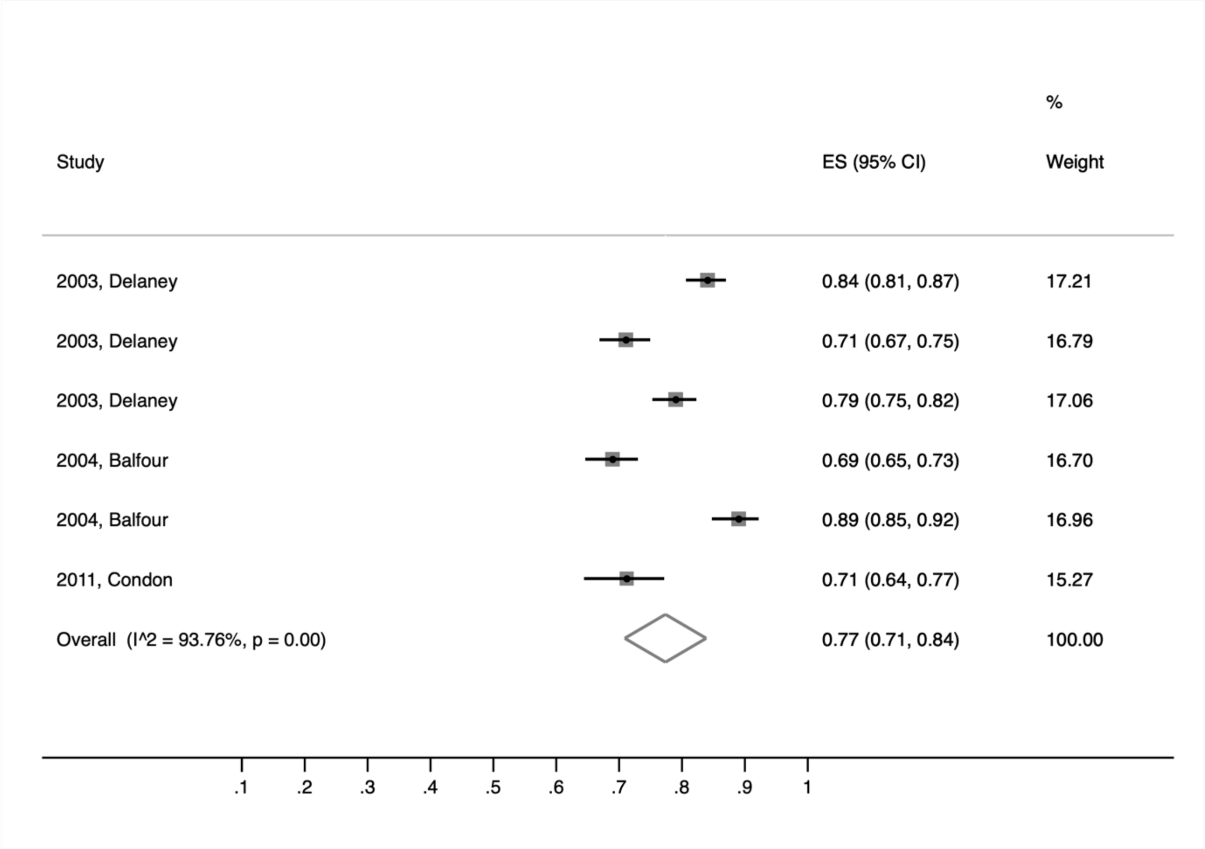
Study
ES (95% CI)
%
Weight
2003, Delaney
0.84 (0.81, 0.87)
17.21
2003, Delaney
0.71 (0.67, 0.75)
16.79
2003, Delaney
0.79 (0.75, 0.82)
17.06
2004, Balfour
0.69 (0.65, 0.73)
16.70
2004, Balfour
0.89 (0.85, 0.92)
16.96
2011, Condon
0.71 (0.64, 0.77)
15.27
Overall (I^2 = 93.76%, p = 0.00)
0.77 (0.71, 0.84)
100.00
.1
.2
.3
.4
.5
.6
.7
.8
.9
1
